# Evolution of the spatial distribution of alcohol consumption following alcohol control policies: a 25-year cross-sectional study in a Swiss urban population

**DOI:** 10.1101/2022.01.16.22269160

**Authors:** David De Ridder, José Sandoval, Rebecca Himsl, Pedro Marques Vidal, Silvia Stringhini, Stéphane Joost, Idris Guessous

## Abstract

**Background:** Alcohol consumption is a major risk factor for multiple chronic diseases and overall mortality. Consequently, multiple alcohol control policies have been implemented in Switzerland. Recent evidence shows that population-wide policies may have impacted some population subgroups and places differently. While the determinants of alcohol consumption are complex, understanding where interventions are needed is necessary for implementing targeted policies. Spatial statistics and modeling allow incorporating this spatial context into the analysis of alcohol consumption and its determinants.

**Methods:** We used the 1993-2018 Bus Santé annual population-based surveys including 18,515 participants aged 35-74 years residing in the canton of Geneva, Switzerland. We assessed the existence of spatial clustering of alcohol consumption the local Moran’s *I* spatial statistics. To evaluate how the spatial distribution of alcohol consumption may have changed over time, we subdivided the study sample into three subsamples according to periods of alcohol-control policies implementation: before (Period 1 (P1): 1993.01.01-1999.06.30), during (Period 2 (P2): 1999.07.01-2009.10.31), and after (Period 3 (P3): 2009.11.01-2018.12.31). We also investigated socio-demographic and built environment determinants of alcohol consumption across periods and provided a comparison of the performance and results of global ordinary least squares (OLS), local geographically weighted regression (GWR), and multiscale GWR (MGWR) models.

**Results:** We found spatial clustering of alcohol consumption in the three analyzed periods. Alcohol consumption in local clusters markedly decreased across periods despite increasing alcohol outlet accessibility. The spatial distribution of higher alcohol consumption clusters (i.e., hot spots) and of lower alcohol consumption clusters (i.e., cold spots) changed slightly across periods. Alcohol consumption in hot spots remained above recommendations in P3 (26.0 g/day, SD ± 17.2 g/day), particularly for women. Cold spots increased in size (4.6% of the participants in P1, 9.3% in P3), particularly in the period following the implementation of policies (P2 to P3) while hot spots remained stable (3.2% of the participants). Alcohol consumption was significantly higher in hot spots than cold spots in all three periods (p < 0.001).

The MGWR model outperformed the OLS and GWR models in most cases and revealed that alcohol consumption was influenced by a mix of global and local relationships. Several of these relationships evolved across periods. Interestingly, in models adjusted for individual-level determinants, we found significant positive relationships between bar density and alcohol consumption in P1 and between density of off-premises alcohol outlets and alcohol consumption in P2. However, these two associations were not significant in P3 potentially due to alcohol control policies.

**Conclusion:** Our results reveal that certain areas and subgroups in need of public health intervention and show that a mix of global and local processes best model alcohol consumption. In addition, this study highlights the potential of geospatial approaches to provide a refined understanding of the determinants of alcohol consumption, and the spatial scales at which they vary.

## Introduction

Alcohol consumption remains a major risk factor of avoidable disease burden [1,2]. It has been associated with an increased risk of developing numerous non-communicable diseases, infectious diseases, and injuries, which together are estimated to represent about 6% of worldwide mortality [1]. Policies, programs, and initiatives aiming at reducing alcohol consumption had overall success in the past decades [3]. In Switzerland, alcohol consumption per capita decreased from around 15 liters of pure alcohol in the 1990s to 10 liters in 2016 [4].

Controlling for the availability of alcohol is a key approach for reducing excessive alcohol consumption [5]. A recent review found that greater outlet density is associated with increased alcohol consumption [6,7]. As in other countries, multiple alcohol control laws were implemented over the last two decades in Switzerland, particularly in the canton of Geneva [4]. In 1999, a cantonal law limiting alcohol and tobacco advertisement was introduced. In 2004, the taxation on alcopop beverages was substantially increased (+300%). In 2005, the legal blood alcohol content driving limited was lowered, off-premises alcohol sale between 9 p.m. and 7 a.m. was interdicted, and an alcohol sale ban in video stores and gas stations were implemented. Moreover, in 2009, a smoking ban in indoor public spaces was introduced. Smoking bans have been suggested to be associated with reduced alcohol consumption [8].

However, population-wide strategies are a one-size-fits-all approach and may not be sufficient for prevention and control. Additionally, these approaches, depending on their design, may not address alcohol-related health inequalities due to unequal responsiveness. For example, in Geneva, despite structural policies implemented over the past decade that probably contributed to a decrease in alcohol consumption, they were not sufficient to reduce the social inequality gap in alcohol consumption, with high consumption persisting in individuals of lower socioeconomic status (SES) [9].

Recent studies have highlighted the existence of spatial clustering of alcohol consumption, raising questions about the factors that may explain such clustering [10–12]. How different policies influence the geographical distribution of alcohol consumption remains unclear.

Health behaviors, including alcohol consumption, are influenced by a complex array of individual-level and social, economic, and built environment factors, themselves shaped by policies [13]. Identifying populations and places where alcohol consumption remains above recommended levels and uncovering how individual and neighborhood-level determinants of alcohol consumption may have evolved following the implementation of alcohol control policies could help define targeted local public health interventions and urban planning policies, complementing global alcohol regulations [11]. However, most existing studies assessing the relationships between individual and neighborhood-level characteristics and alcohol consumption did not account for spatial dependence between individuals [14–17]. The presence of spatial dependence may bias the obtained results in meaningful ways. Notably, it violates the assumption of independence of the studied observations required by traditional linear and multilevel regression models [18].

Using a large, repeated cross-sectional population-based surveys spanning 25 years (1993-2018), we aimed to examine the existence of spatial clusters of alcohol consumption in three periods defined by the implementation of alcohol control policies: i) before (1993.01.01-1999.06.30), ii) during (1999.07.01-2009.10.31) and iii) after implementation (2009.11.01-2018.12.31). A secondary objective was to investigate the individual and neighborhood-level socioeconomic and build-environment determinants of alcohol consumption and whether these determinants may have changed across periods.

## Methods

### Study design and sample

Data for this study was collected through the Bus Santé study, a continuing population-based study in the canton of Geneva (population of approximately 500,000 inhabitants in 2021), monitoring health and cardiovascular risk factors. As previously described [19], independent samples of residents were subjected to health examination surveys since 1993. The Bus Santé study’s recruitment methods have been described in details elsewhere [19,20]. The geocoded residential address of each participant was used for spatial analyses. Since using small units of analysis allows increased resolution of spatial analysis, we used individual points to avoid the bias resulting from the modifiable areal unit problem (MAUP), which affects results when point-based measures are aggregated [21]. The Bus Santé study was approved by the Cantonal Research Ethics Commission of Geneva, Switzerland (PB 2016-00363).

### Periods of alcohol control policies

We defined three time periods to enable analyses and comparisons before, during, and after implementing alcohol control policies. The period from 1993.01.01 to 1999.06.30 correspond to the first period (P1) before implementing alcohol control policies. The period 1999.07.01-2009.10.31 corresponds the second period (P2), during which a set of policies to control alcohol consumption were implemented in Switzerland and the canton of Geneva particularly. Finally, the period 2009.11.01-2018.12.31 corresponds to the period after implementing alcohol control policies (P3). Separating each policy into specific periods according to their implementation date would have been of interest to study their potential impact. However, this would have created several short periods with few participants thus reducing power for spatial and statistical analyses.

### Outcome variable

Total daily alcohol intake was determined using a self-administered, semiquantitative food frequency questionnaire (FFQ) originally developed and validated against 24-hour dietary recalls [22]. The FFQ considers consumption frequency, ranging from “never during the last four weeks” to “two or more times per day”, and the type of alcohol beverage (wine, beer, aperitifs, champagne, and spirits like liqueur, brandy, or whisky) and average serving size compared with a 10g alcohol standard for each beverage (similar, bigger or smaller) [9]. The same FFQ was used throughout the entire study period (1993-2018), and data derived from this FFQ has integrated large international consortia [23,24].

### Individual and neighborhood-level covariates

Individual-level covariates included gender, age, educational attainment (tertiary education: yes or no), civil status (married-cohabiting: yes or no), nationality (Swiss-born, foreign-born), occupational level (low—manual and lower occupations, medium—non-manual occupations, high—professional and intermediate professions, and not working), and smoking status (current smoker: yes or no). Individual income data being missing for participants before 2007, we used the annual neighborhood median household income for the years from 2005 to 2016. We assigned this area-level income to participants based on their year of participation and their corresponding statistical subsector in Geneva (GIRECs) – a neighborhood definition created by the canton of Geneva (Office Cantonal de la Statistique, www.ge.ch/statistique). Participants from 1993 to 2005 were assigned neighborhood income values from 2005, and participants from 2016 to 2018 were attributed values from 2016.

### Physical accessibility to alcohol outlets

We obtained the dataset of all registered companies in the canton of Geneva compiled by the the “Répertoire des entreprises de Genève” (REG) [25] and made available by the “Système d’information du territoire à Genève (SITG)” [26]. Yearly listings of all the companies registered in the canton were available from 2003 to 2018 (excepted 2006 and 2007). They contained designations that allowed us to disaggregate by outlet type, including off-premise outlets (convenience stores, grocery shops, gas stations, and supermarkets) and on-premise outlets (restaurants, nightclubs, and bars). We excluded the category “gas stations” from the calculation of off-premises outlet density after 2005 to consider the implementation of an alcohol sale ban in these amenities starting that year. Geographic coordinates of each amenity were provided in the dataset. For each participant, we assessed the density of each alcohol outlet category through street network-based accessibility analyses. To consider the evolution in the built environment over the study period (1993-2018), we matched the participation year of each participant to the closest year available in the alcohol outlet dataset (Table S1).

For each alcohol outlet category, alcohol outlet density was calculated as the sum of alcohol outlets in participants’ neighborhoods defined as 1,200m street network buffers. The street network of the canton of Geneva was obtained using the OSMnx Python package [27]. The study participants and alcohol outlets were snapped to the nearest segment of the street network using a maximum snapping distance of 50m. The Pandana Python package was used to create the street network buffers and perform the accessibility analyses [28]. A linear decay was used to give more weight to nearby locations compared to farther ones. The distance of 1,200m was selected based on the results of the global autocorrelation analysis and corresponded to a walking time of around 15min, which has been considered relevant to individuals’ residential spatial scale [29].

### Exclusion criteria

We included participants with ages between 35 and 74 years, the age group that has been consistently recruited since the beginning of the Bus Santé study. We excluded abstinent participants (n = 3,568, 15.7%) and participants with missing data for alcohol consumption (n = 53, 0.2%) on civil status (n = 25, 0.1%), educational attainment (n = 222, 1.2%), nationality (n = 91, 0.5%) and neighborhood median household income (n = 294, 1.6%). Data from the remaining 18,515 individuals (P1, n = 5,215; P2, n = 7,013; P3, n = 6,287) were used for the analyses (Table 1).

**Table 1.** Socio-demographic characteristics of the Bus Santé participants, Geneva, Switzerland (1993-2018)

|  | Period 1 (P1) | Period 2 (P2) | Period 3 (P3) |
| --- | --- | --- | --- |
| Sample size (n) | 5,215 | 7,013 | 6,287 |
| Age, mean (SD) | 51.3 (10.5) | 51.7 (10.9) | 52.2 (11.0) |
| Neighborhood median household income (CHF), mean (SD) | 122,989.8 (36,573.3) | 127,293.0 (39,729.3) | 138,323.2 (45,052.5) |
| Sex (%) |  |  |  |
| Woman | 2,423 (46.5) | 3,286 (46.9) | 3,015 (48.0) |
| Man | 2,792 (53.5) | 3,727 (53.1) | 3,272 (52.0) |
| Marital status (%) |  |  |  |
| Married/cohabiting | 4,113 (78.9) | 5,241 (74.7) | 4,309 (68.5) |
| Not married/cohabiting | 1,102 (21.1) | 1,772 (25.3) | 1,978 (31.5) |
| Education |  |  |  |
| Tertiary | 1,595 (30.6) | 2,630 (37.5) | 3,119 (49.6) |
| Lower than tertiary | 3,620 (69.4) | 4,383 (62.5) | 3,168 (50.4) |
| Occupational level |  |  |  |
| Not working | 140 (2.7) | 167 (2.4) | 161 (2.6) |
| Low | 2,119 (40.6) | 3,023 (43.1) | 3,081 (49.0) |
| Medium | 1,902 (36.5) | 2,428 (34.6) | 1,899 (30.2) |
| High | 1,054 (20.2) | 1,395 (19.9) | 1,146 (18.2) |
| Smoking status (%) |  |  |  |
| Smoker | 1,188 (22.8) | 1,741 (24.8) | 1,313 (20.9) |
| Non-smoker | 4,027 (77.2) | 5,272 (75.2) | 4,974 (79.1) |
| Nationality (%) |  |  |  |
| Swiss | 3,781 (72.5) | 5,073 (72.3) | 4,305 (68.5) |
| Other | 1,434 (27.5) | 1,940 (27.7) | 1,982 (31.5) |

Note: SD, standard deviation

### Regression analyses

The assumptions of traditional global regression models such as ordinary least squares (OLS) might not be satisfied in the presence of spatial autocorrelation as these assume that relationships are constant across the study area [18,30]. However, many individuals and environmental characteristics are spatially dependent. Local models such as the geographically weighted regression (GWR) have been introduced to relax this assumption of spatial stationarity by allowing relationships to vary over space and obtains local parameter estimates by calibrating a locally weighted regression at each location of the dataset [31]. GWR generally provides increased model fit and reduced residual spatial autocorrelation compared to traditional global models that assume that relationships are constant over space [32,33].

Local regressions are fitted through a distance-weighting scheme using a spatial weights matrix that defines the relationships between observations and places more weight on closer data points than remote ones [34]. The spatial weights matrix is defined by three characteristics, a kernel function, a bandwidth, and proximity measure. We used a bi-square kernel function with a bandwidth parameter based on the number of nearest neighbors away from the calibration point that minimizes a corrected Akaike information criterion (AICc). The bi-square function presents the advantage of assigning a weight of zero to observations outside the bandwidth, facilitating interpretation compared to the Gaussian kernel function that may retain non-zero weights for all observations. One limitation of the GWR is that it uses the same bandwidth for all relationships, thus assuming that all spatial processes vary at the same scale [34]. A recent extension to the GWR, the multiscale GWR (MGWR), relaxes this assumption using a different bandwidth for each relationship, ensuring that a correct data-borrowing scale is employed, thus capturing spatial heterogeneity more accurately [31].

To investigate the relationships between individual and neighborhood-level socio-demographic and environmental characteristics and daily alcohol intake (g/day), we first calibrated global ordinary least squares (OLS) regression models at each period (i.e., P1, P2, and P3). The built-environment explanatory variables included in the multivariate regression models were selected using univariate OLS models and based on statistical significance (p < 0.05) and a variance inflation factor (VIF) below 7.5. Next, we assessed the presence of spatial autocorrelation in the residuals using the global Moran’s I statistic using a fixed distance bandwidth of 1,200m. Subsequently, GWR and MGWR models were calibrated for the three periods. Model selection between OLS, GWR, and MGWR was done using AICc, examining overall model fit and complexity; lower AICc values indicating a better fit.

### Global spatial clustering

We assessed the presence of overall spatial dependence in the outcome variable with the Global Moran’s I statistic, which is the most commonly used statistic to test for global spatial autocorrelation [35]. Each observation is analyzed within the context of those neighboring observations within a specified critical distance. Relationships between observations are defined using a spatial weights matrix. To determine the distance corresponding to the maximum spatial autocorrelation, the distance at which the spatial process is the most pronounced, we computed the Global Moran’s I at incremental fixed distance bands (200m, 400m, 600m, 800m, 1,000m, and 1,200m) and the distance at which the statistically significant z-score peaked was selected. Observations within the distance band are assigned the same weight, and observations outside the selected distance are assigned a zero weight and thus do not influence calculations for that observation. Statistical significance was assessed through a Monte Carlo procedure consisting of 999 random permutations, which return a pseudo-p-value associated with the Moran’s I statistic.

### Local spatial clustering

Global clustering statistics evaluate spatial clustering in the study area but cannot detect where local clusters may exist. Investigating the presence of local spatial clustering in alcohol consumption through spatial cluster detection provides insights into the impact of alcohol control policies on the spatial distribution of alcohol consumption. Univariate local spatial cluster analysis was conducted by calculating the local Moran’s I statistic [36]. This analysis returns a local Moran’s I value, a z-score, a pseudo-p-value, and a category representing the cluster type for each statistically significant observation. There are four categories of statistically significant values, hot spots (High-High), low-spots (Low-Low), and spatial outliers: high value surrounded by low values (High-Low) and low value surrounded by high values (Low-High). Observations with a p-value smaller than 0.05 are considered statistically significant. To investigate whether individual and neighborhood-level socio-demographic characteristics and alcohol availability may explain local clustering of alcohol consumption, we calculated and mapped the local Moran’s I categories before and after adjustment for covariates using the best performing model among OLS, GWR, and MGWR. After adjustment, a reduction of local clusters signifies that the covariates in the adjustment model explained the spatial clustering.

### Changes in the characteristics of high-high and low-low spatial clusters of alcohol consumption

To investigate a potential change in the characteristics of individuals belonging to hot spots and cold spots of alcohol consumption across the three periods, we compared their socio-demographic and alcohol availability characteristics using Welch’s t-tests and Fisher exact tests for numeric and binary variables, respectively. We presented these results using radar plots to optimize the visualization of many studied explanatory variables.

## Results

### Descriptive statistics

Sample individual and neighborhood-level characteristics are described in Table 1. There was significant variation in the physical access to alcohol outlets due to the study area comprising rural and urban areas. Globally, access to alcohol outlets increased from period one to period three, with a stronger trend for on-premises outlets.

Median daily alcohol consumption stratified by period and individual characteristics are provided in Table 2. Over half of the respondents of each period were male. This slight imbalance can be explained by the exclusion of non-drinkers which are predominantly women. The mean age was around 50 years for all three periods.

**Table 2.**
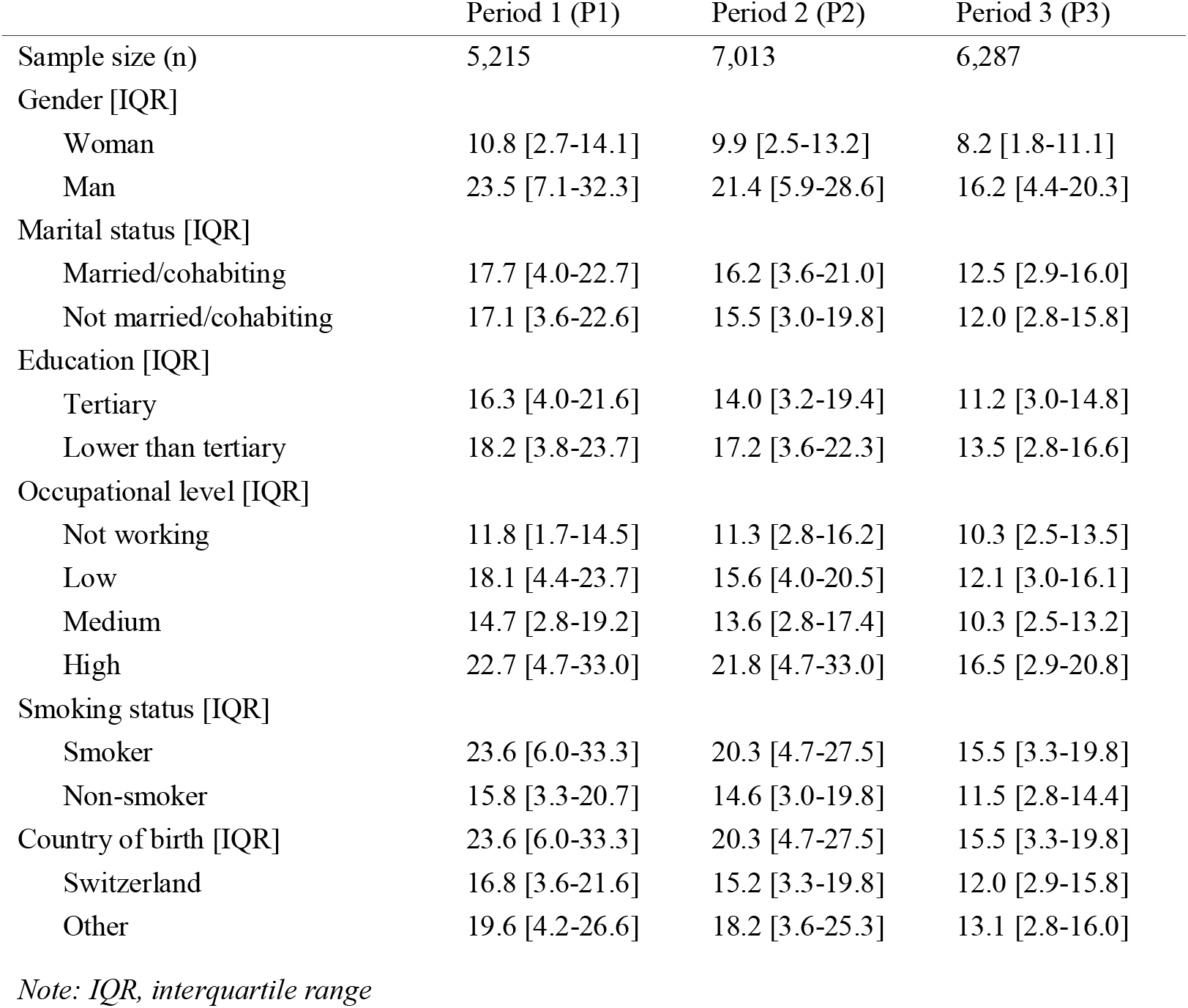
Alcohol intake (g/day) of the Bus Santé participants, Geneva, Switzerland (1993-2018) stratified by individual-level characteristics.

### Regression analyses

The univariate OLS models showed that categories of alcohol outlets were significantly associated with alcohol consumption, at different periods. Bar density had a positive relationship with alcohol consumption in P1, and both off-premises outlet density and on-premises outlet density had statistically significant positive relationship with alcohol consumption in P2. No significant association between measures of alcohol outlets density and alcohol consumption were identified in P3.

We calibrated a separate multivariate OLS model for each of the three retained alcohol outlet categories (i.e., bars, off-premises, and on-premises) for each period. All models controlled for individual and neighborhood-level socio-demographic covariates. The coefficient estimates and standard errors are presented in table S2. In P1, age, smoking status, a high occupational level, neighborhood median income, and bar density were positively associated with alcohol consumption. Being a woman had a strong negative relationship with alcohol consumption and, to a lesser degree, having Swiss nationality. In P2, the same relationships were observed apart from neighborhood median income, which was not significantly associated with alcohol consumption. Additionally, while bar density was not associated with alcohol consumption, the density of on-premises and off-premises alcohol outlets had a relatively strong positive relationship with alcohol consumption. Finally, in P3, we found a negative relationship between tertiary education and alcohol consumption and no statistically significant relationships between alcohol outlets density and alcohol consumption. Of note, gender, age, high occupational level, and smoking status remained associated with alcohol consumption, but to a lesser extent than in P1 and P2. The global spatial autocorrelation analysis in the models’ residuals revealed statistically significant global Moran’s I in several models (Table S3), suggesting that global OLS models may be inappropriate.

GWR models were calibrated using the same sets of explanatory variables used in global models. In P1 and P2, the optimal bandwidths included all nearest neighbors (i.e., global scale), indicating that all the data was included in each local subset. In P3, the optimal bandwidths were relatively more regional, with 4,327 nearest neighbors in model 1, 3,676 in model 2, and 3,763 in model 3. GWR models showed mixed performance. Values of *R*^2^ were slightly higher in P1 and P2 and much higher in P3. In terms of AIC, in P1 and P2, GWR showed a worse model fit than OLS, while in P3, GWR outperformed the global model. Finally, residual spatial autocorrelation remained in several instances. Therefore, a MGWR model may be necessary to allow processes to vary at unique scales.

The MGWR model showed improved performance in most cases (Table S3), with lower AICc and reduced residual spatial autocorrelation. The calibration of a MGWR model produced an optimized bandwidth for each explanatory variable, describing the spatial scale at which each spatial process varies (Table S4) [34]. Relationships are thus not forced to be global or local a priori. Indeed, we observed that certain relationships occurred globally, indicated by a bandwidth comprising all neighbors, while others had small bandwidths indicating that they vary locally (Fig S1-3). This resulted in lower AIC and AICc and higher *R*^2^.

We only presented parameter estimate surfaces for the best performing model of each period, model 1 in P1 (Fig S1) and model 3 in P2 (Fig S2) and P3 (Fig S3). Several spatial patterns can be observed thanks to the MGWR parameter estimates surfaces (Fig S1-3, right) in comparison to those from GWR (Fig S1-3, left). First, few parameters are effectively global with a statistically significant relationship with alcohol consumption: intercept, gender, nationality, neighborhood median income in P1; intercept, gender, high occupational level, and off-premises outlet density in P2; and none in P3. These showed little or no spatial variation and are in accordance with the GWR models where bandwidths were global in P1 and P2 and more local in P3. The negative or positive character of these global relationships was consistent with the results of the global regression models. Second, some parameters were found to be only statistically significant in some areas where there is little spatial variation: bar density in P1; tertiary education and nationality in P2 and P3. Again, the observed relationships with alcohol consumption agree with global models. Third, another category included surfaces with statistically significant parameters that vary locally: age and smoking status in P1 and P2; and intercept, age, gender, and smoking status in P3. Finally, some parameters were not statistically significant across the study area: civil status, educational attainment, occupational level in P1; civil status, medium- and low occupational level, neighborhood-level median income in P2; civil status, low and medium occupational level, neighborhood-level median income, off-premises outlet density in P3.

### Global spatial autocorrelation

There were spatial patterns in the physical accessibility to alcohol outlets in all periods. Global Moran’s I values for most measures of physical accessibility were statistically significant (p = 0.001). The density of participants and the high variability of daily alcohol consumption made it impossible to detect spatial patterns by simple observation. The global Moran’s I calculation at incremental fixed distance bands showed that spatial autocorrelation in alcohol consumption peaked with a 400m fixed-distance band in P1 and a 1,200m band in P3 (Table S2). Alcohol consumption showed no statistically significant spatial autocorrelation in P2. We conducted spatial analyses using both 400m and 1,200m distance bands, but only results for 1,200m are presented. We preferred 1,200m over 400m because many participants, especially in rural areas, would not have neighbors at that distance, thus excluding them for spatial analyses. Both distances showed similar spatial patterns.

### Local clustering of alcohol consumption

We found local spatial clusters of alcohol consumption in each period. Spatial clusters of high and low daily alcohol consumption for the three periods are mapped in Fig 1-3. The spatial distribution of clusters was relatively stable across periods, with large hot and cold spots appearing in the same areas (Fig S4). The percentage of individuals in cold spots decreased from P1 to P2 and doubled from P2 to P3. The percentage of individuals in hot spots remained more stable in size. From P1 to P2, it showed a slight increase that reduced to P1 levels in P3. P2 shows a distinctive pattern with a large hot spot located in the urban part of the canton of Geneva (Fig 2A).

**Figure 1.**
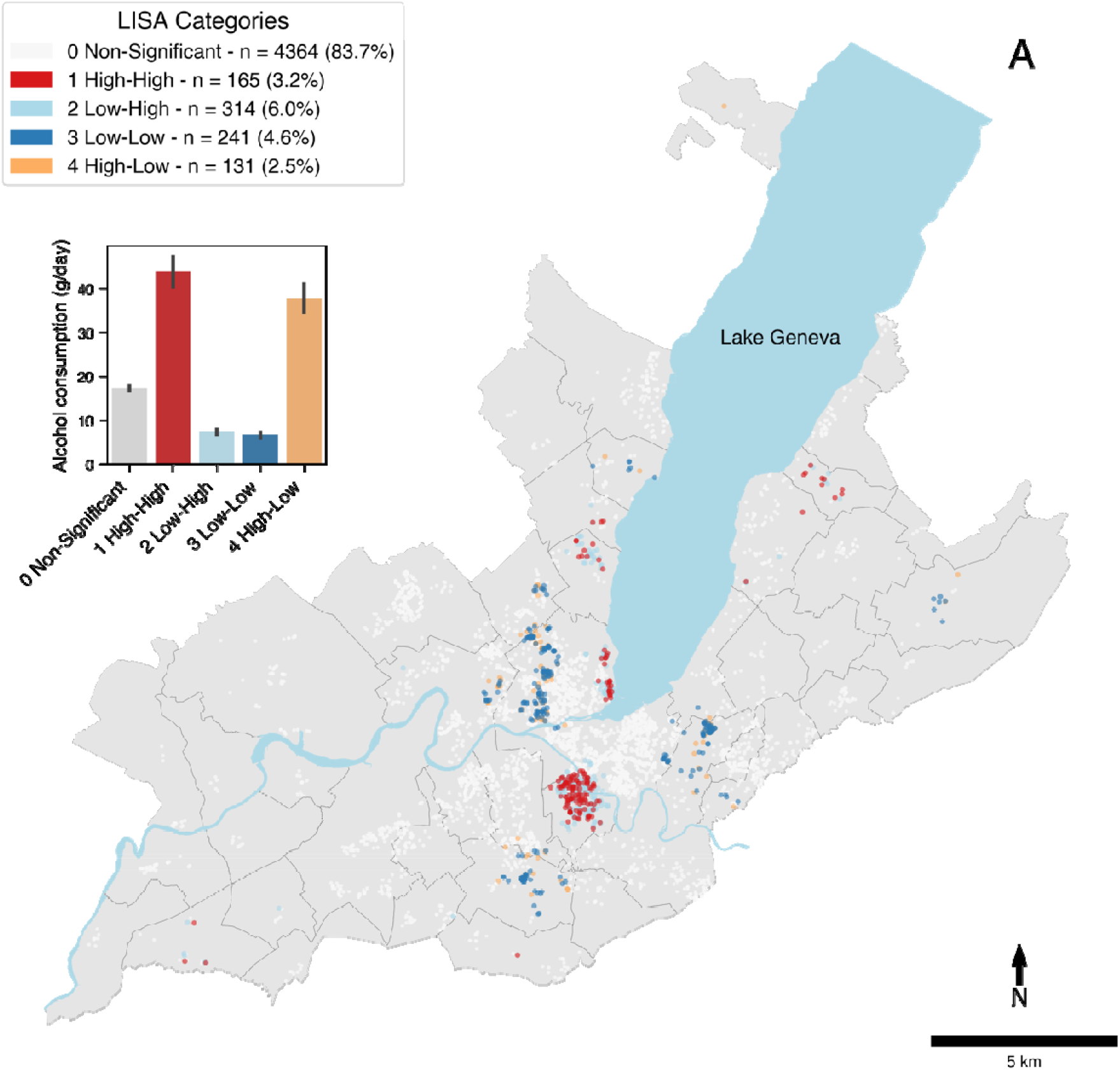
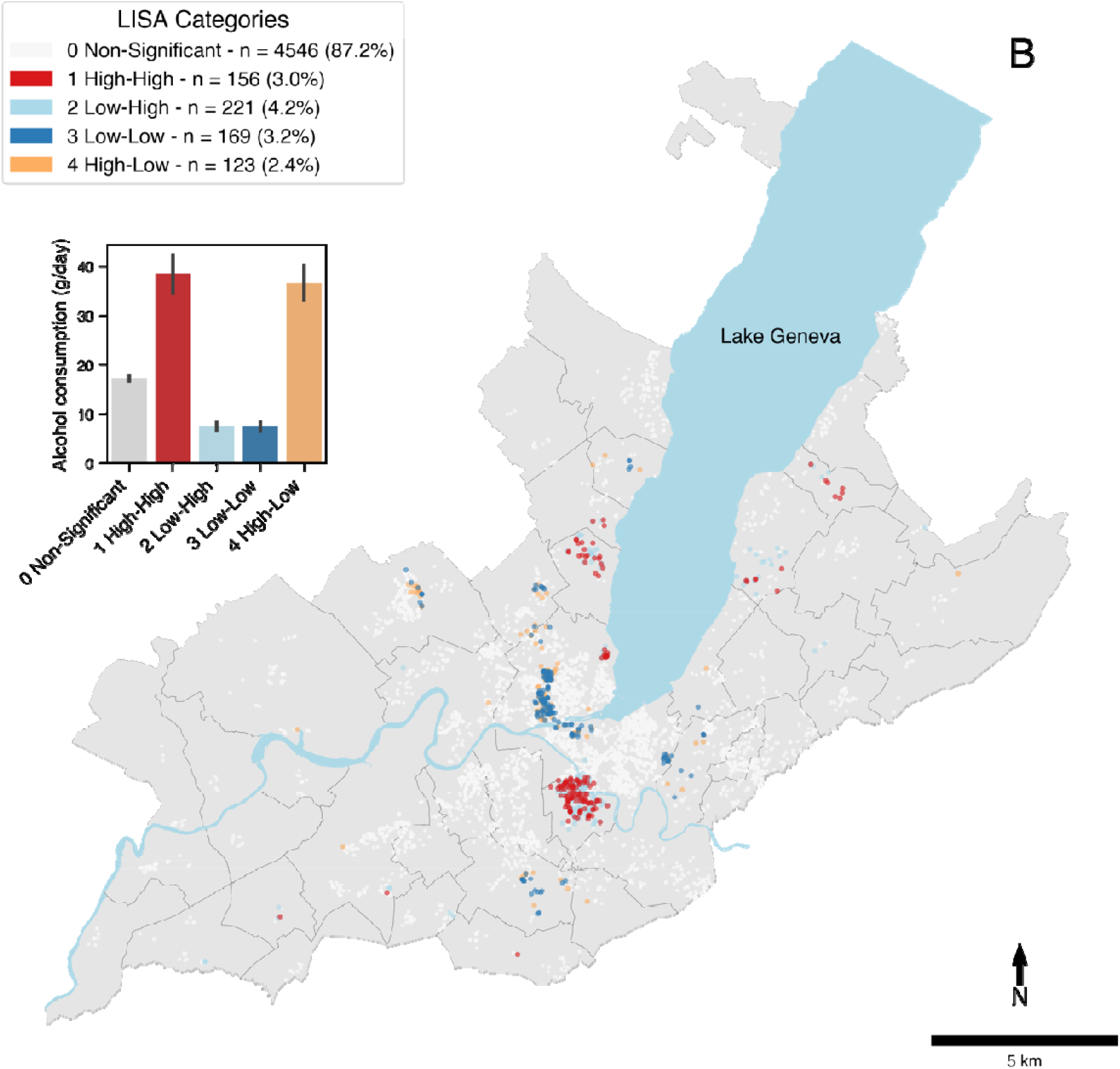
Local spatial clustering of alcohol consumption in period 1 (P1 – 1993.01.01-2009.06.30). Local Moran’s I spatial clusters of alcohol consumption (A) unadjusted and (B) adjusted for socio-demographic characteristics and physical accessibility to bars using a multiscale geographically weighted regression (MGWR model 1). The dark red markers (1 High-High) correspond to the individuals with a high alcohol consumption surrounded by individuals with a high alcohol consumption. The light blue markers (2 Low-High) correspond to individuals with a low alcohol consumption surrounded by individuals with a high alcohol consumption. The dark blue markers (3 Low-Low) correspond to individuals with a low alcohol consumption surrounded by individuals with a low alcohol consumption. The light red markers (4 High-Low) correspond to individuals with a high alcohol consumption surrounded by individuals with a low alcohol consumption. White markers are not significant at = 0.05. Black lines correspond to municipality delimitations. The bar plot shows the average alcohol consumption (g/day) according to LISA cluster categories. Error bars show 95% confidence intervals.

**Figure 2.**
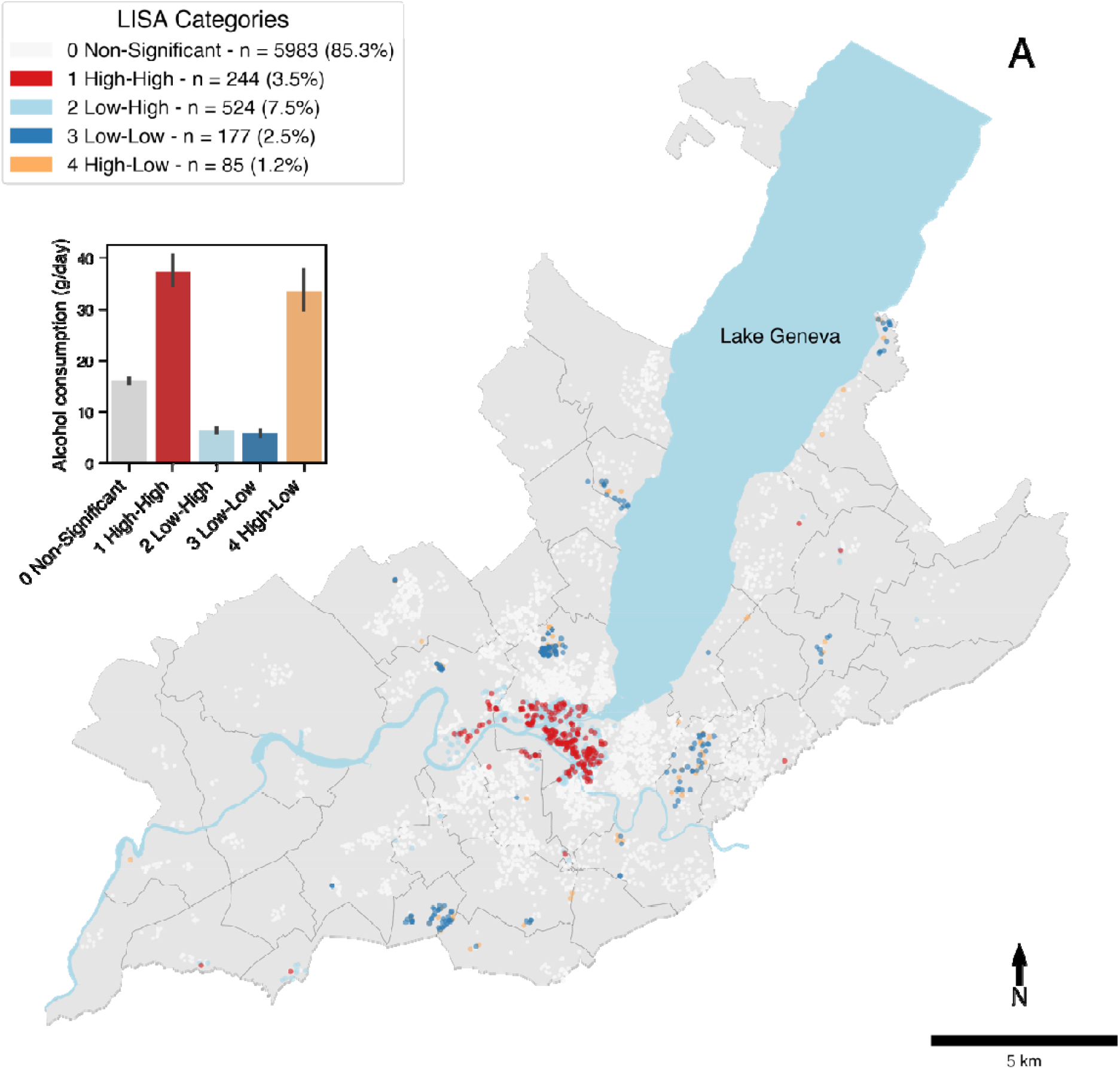
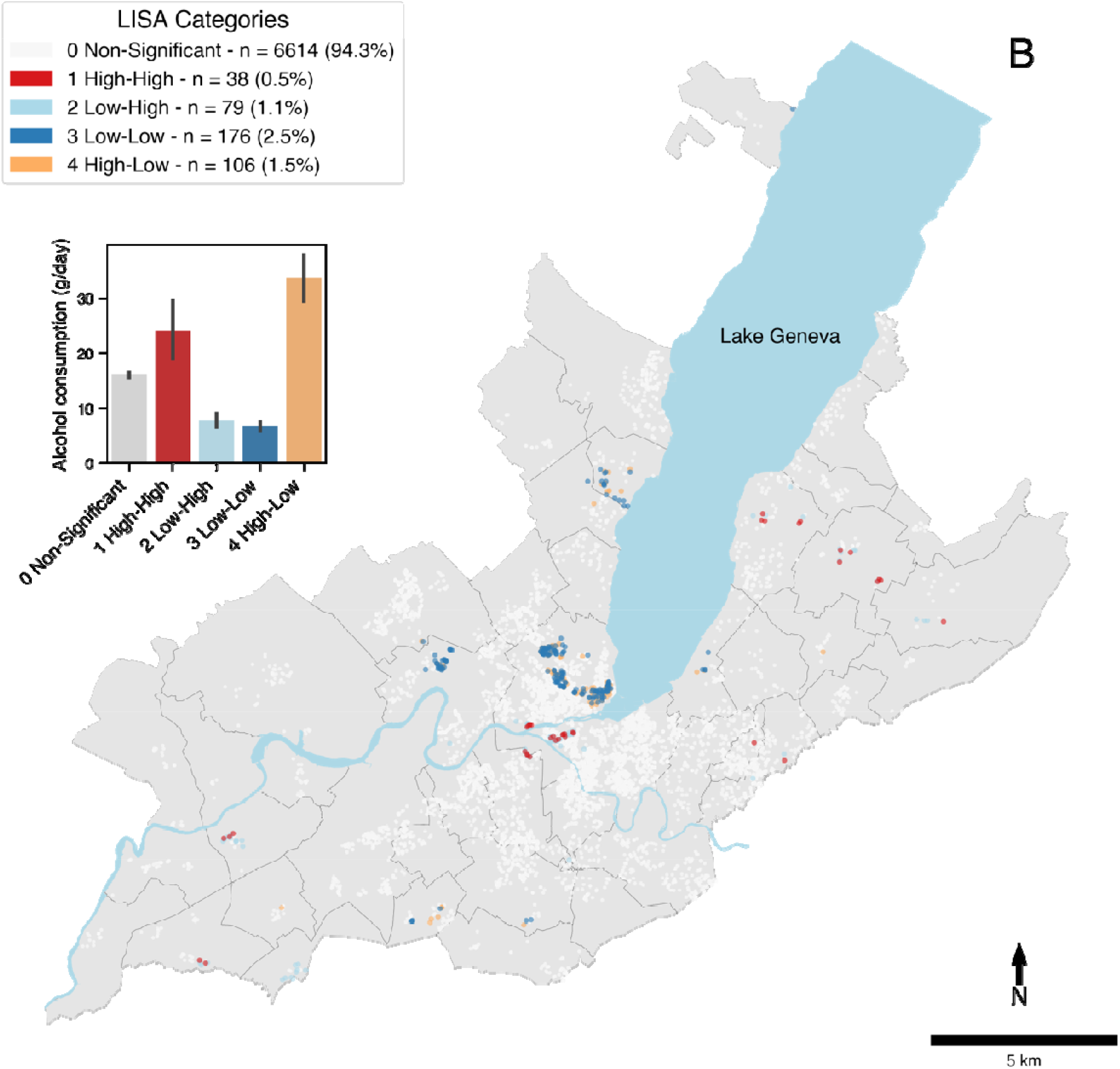
Local spatial clustering of alcohol consumption in period 2 (P2 – 1999.07.01-2009.10.31). Local Moran’s I spatial clusters of alcohol consumption (A) unadjusted and (B) adjusted for socio-demographic characteristics and physical accessibility to off-premises alcohol outlets using a multiscale geographically weighted regression (MGWR model 3). The interpretation of local cluster categories remains the same as described on Fig 1.

**Figure 3.**
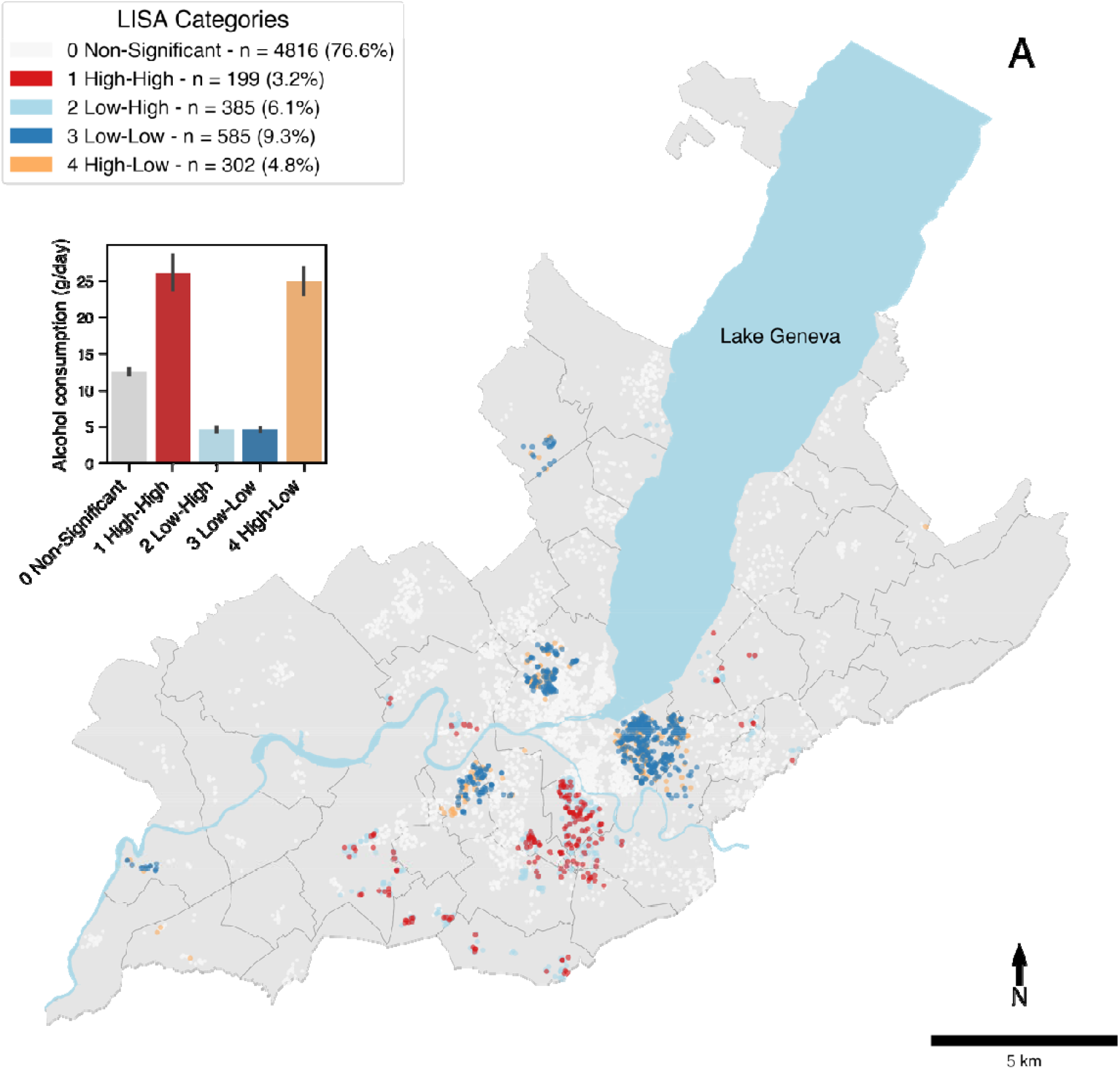
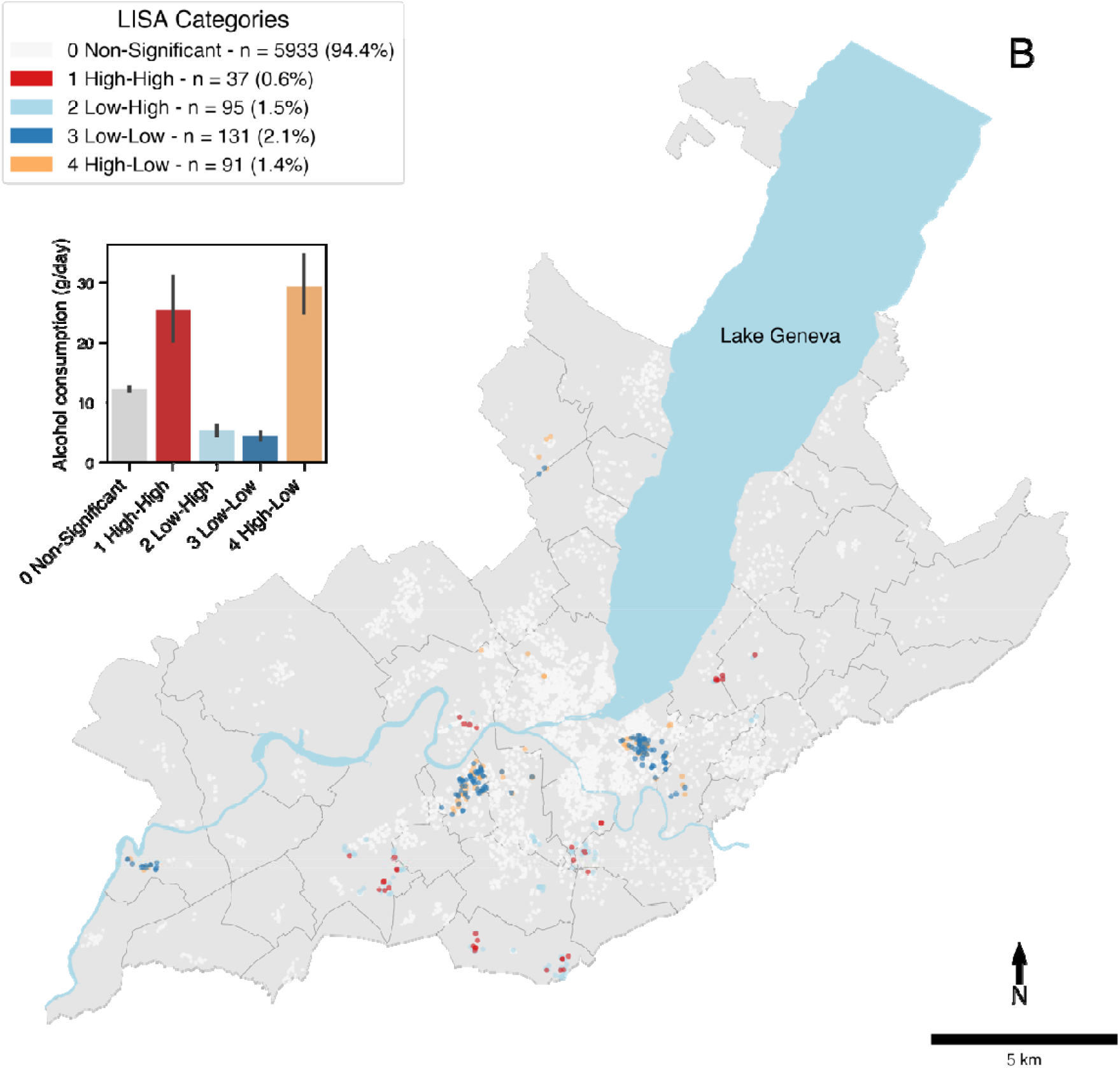
Local spatial clustering of alcohol consumption in period 3 (P3 – 2009.11.01-2018.12.31). Local Moran’s I spatial clusters of alcohol consumption (A) unadjusted and (B) adjusted for socio-demographic characteristics and physical accessibility to off-premises alcohol outlets using a multiscale geographically weighted regression (MGWR model 3). The interpretation of local cluster categories remains the same as described on Fig 1.

The local cluster analyses of adjusted alcohol consumption, using the MGWR residuals, showed that local spatial clusters were greatly attenuated in P2 and P3, suggesting that the explanatory variables in the models were responsible for most of the spatial clustering (Fig 1-3B). In P1, the clusters remained stable after adjustment.

### Characteristics of local spatial clusters of alcohol consumption

Individuals belonging to hot and cold spots had significant differences in socio-demographic characteristics and access to alcohol outlets.

In P1, individuals in the high-high category had an average alcohol consumption of 43.8g/day (SD ± 22.5g/day), while individuals in the low-low category had an alcohol consumption of 6.7g/day (SD ± 5.1g/day). Alcohol consumption was significantly higher in hot spots than in cold spots in all three periods (p < 0.001). Significance was assessed with Welch’s t-tests. The proportion of high occupational level individuals, smokers, men, and physical accessibility to alcohol outlets were higher in the high-high category. Neighborhood median household income and the proportion of individuals with a medium occupational level were higher in the low-low category (Fig 4A).

**Figure 4.**
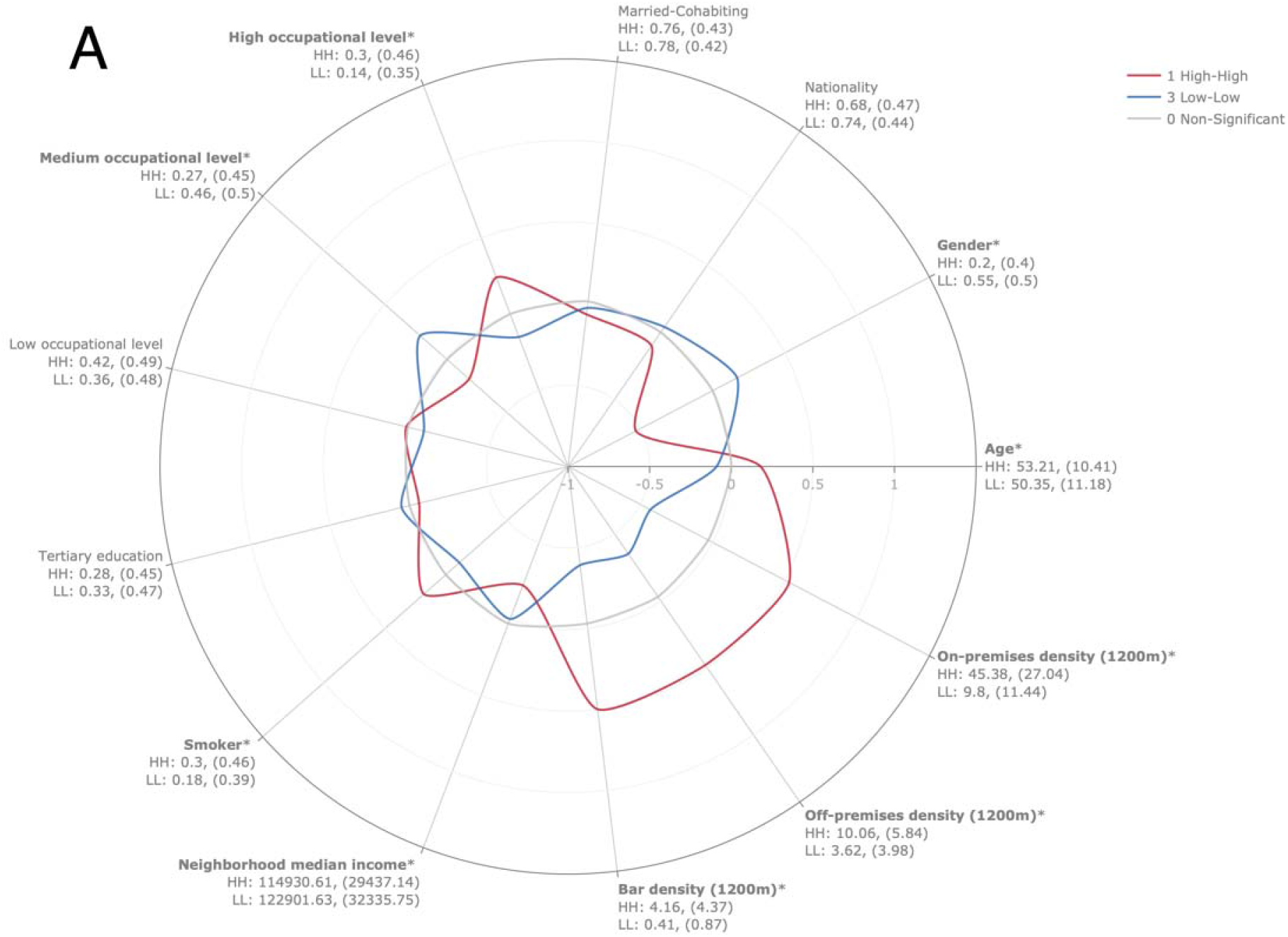
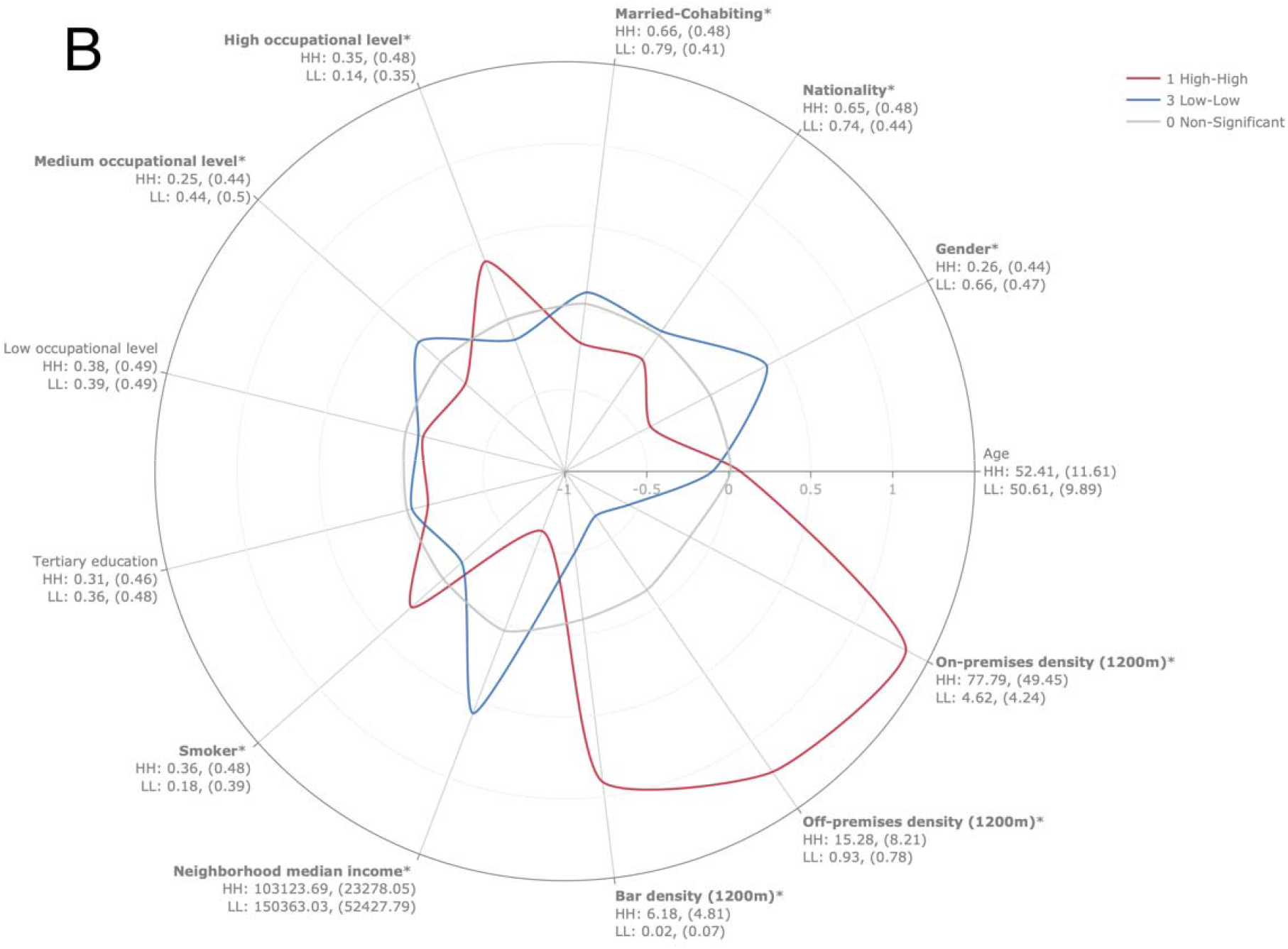
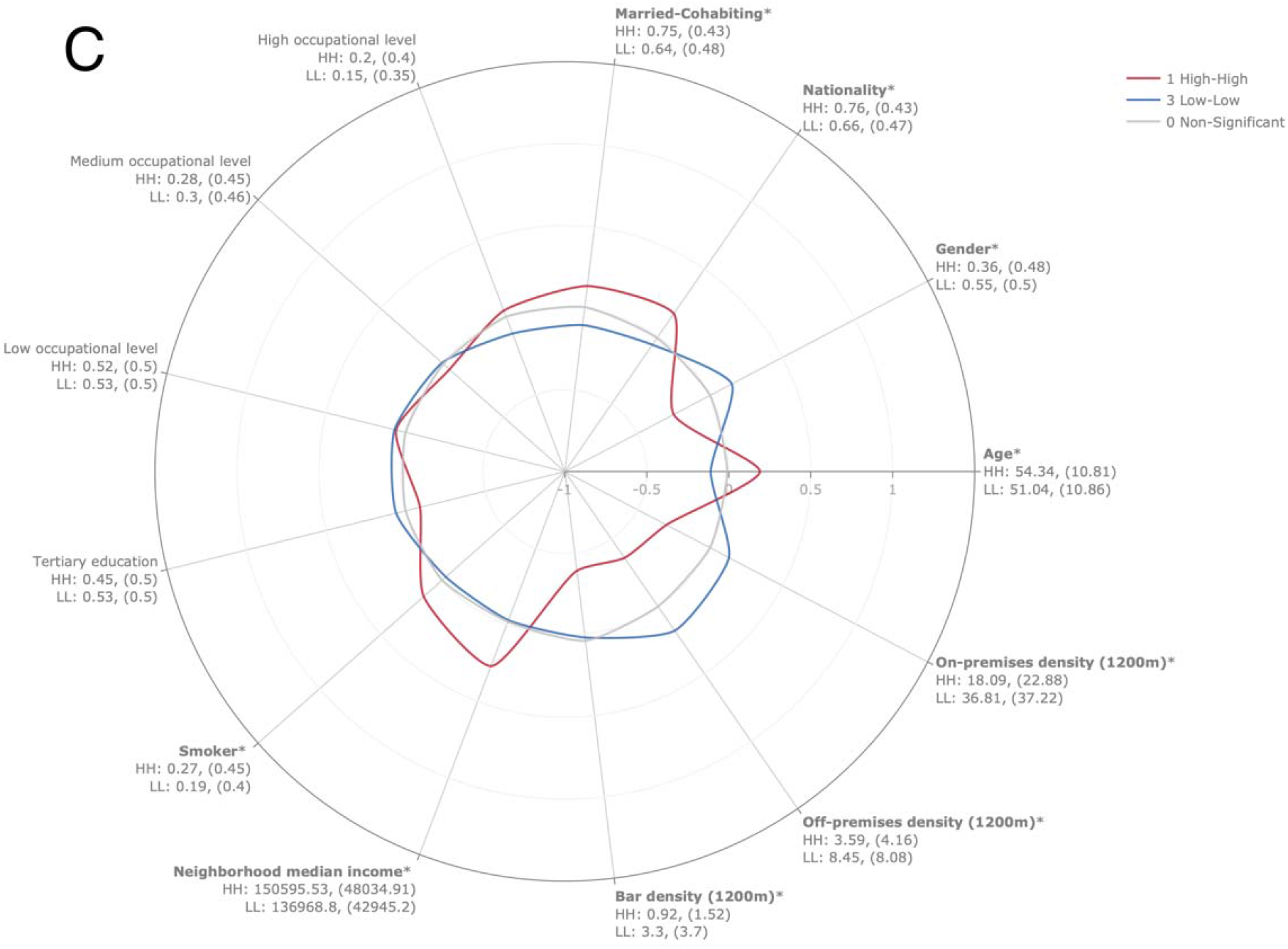
Characterization of local spatial clusters. Radar plot of standardized socio-demographic characteristics and physical alcohol availability in high-high and low-low spatial clusters for (A) period (P1), (B) period 2 (P2) and (C) period 3 (P3). Mean, (SD) is provided under each characteristic. HH, high-high class; LL, low-low class. Statistical significance evaluated using Welch’s t-tests for continuous variables and Fisher’s exact tests for binary variables. *p-value < 0.05.

In P2, the differences in socio-demographic characteristics and alcohol accessibility were exacerbated, notably concerning physical accessibility to alcohol outlets, which was much higher for individuals in the high-high category (Fig 4B). Additionally, high-high clusters had a lower neighborhood median income and a higher proportion of smokers. Of note, nationality and the proportion of married and cohabiting individuals were significantly higher in the low-low category. The average alcohol consumption decreased among individuals in the high-high category compared to P1 (37.3g/day, SD ± 22.3g/day). The consumption remained relatively stable in the low-low category (5.8g/day, SD ± 4.3g/day).

Finally, in P3, which corresponds to the period after the implementation of alcohol control policies, differences were strongly attenuated or even inversed (Fig 4C). The high-high category had lower physical accessibility to alcohol outlets than the low-low category. While inversed, the difference was less contrasted than in P1 and P2. The high-high category had a significantly higher proportion of Swiss nationality, married or cohabiting individuals, and individuals with a high occupational level. Additionally, individuals in the high-high category were significantly older than those in the low-low category. The proportion of individuals having a tertiary education was significantly higher in the low-low category. The average alcohol consumption among individuals in the high-high category decreased starkly (26.0g/day, SD ± 17.2g/day) compared to P2, yet it remained at levels harmful for health. The alcohol consumption in the low-low category decreased slightly to 4.6g/day (SD ± 3.2g/day).

## Discussion

In this study, we detected spatial clustering of alcohol consumption in the canton of Geneva and changes in their spatial distribution before, during, and after the implementation of alcohol control policies. We found significant associations between alcohol consumption, individual and neighborhood-level socio-demographic characteristics and physical availability of alcohol outlets. Additionally, several of these associations evolved across periods. The results also demonstrate the potential of local models to improve the understanding of the determinants of alcohol consumption and the spatial scales at which they influence it.

In the first period (P1—1993.01.01-1999.06.30), individuals in hot spots of alcohol consumption had an average alcohol consumption much higher than the recommendation for low-risk alcohol consumption of 10g/day for women and 20g/day for men [37]. We found that average alcohol consumption in hot spots decreased between P1 and P2 (1999.07.01-2009.10.31) and, even more importantly, between P2 and P3 (2009.11.01-2018.12.31), despite increasing physical availability of alcohol outlets. In P3, alcohol consumption was closer to recommendations yet remaining inadequate, particularly for women. This highlights the potential of individual-level spatial analyses in uncovering precise areas and populations at risk of harmful chronic alcohol consumption and that would benefit from specific public health interventions. Studies conducted at lower spatial resolution may have missed such local variations in alcohol consumption behaviors due to the lost variance inherent to spatial aggregation [21,38].

Although at a much coarser spatial scale, local spatial clustering of alcohol consumption was previously reported in the literature [39]. Another study using individual-level data found no significant spatial clustering in alcohol consumption [11]; however the studied population were adolescents. Interestingly, the observed hot spots of alcohol consumption were much smaller than hot spots previously reported in the canton of Geneva for other risk factors such as body mass index, sugar-sweetened beverages, and salt intake [40–42]. Of note, hot spots decreased, and cold spots increased in size from P2 to P3, the period after implementing alcohol-control policies. Together, this suggests that the alcohol reduction strategies implemented in P2 were effective beyond reducing overall consumption [4,9] through a reduction in the size of consumption hot spots.

The adjustment for socio-demographic and physical alcohol availability led to an almost complete disappearance of spatial clusters of alcohol consumption, suggesting that these factors were responsible for most of the spatial clustering. However, other factors not accounted for in this study could explain the remaining spatial patterns. For example, we did not consider alcohol temporal and price availability, which has been reported to affect consumption [5,14]. Furthermore, non-spatial determinants of alcohol consumption have been reported in the literature, including the critical influence of individuals’ social networks [43].

Our regression analyses, notably using the modern MGWR modeling technique, highlighted the existence of both global and local relationships between socio-demographic and physical availability of alcohol outlets and alcohol consumption. These relationships changed across periods, which could be due to the implementation of alcohol control policies. For example, there was a positive association between physical accessibility to bars and alcohol consumption in P1 and on-premises and off-premises alcohol outlets in P2, while no significant association was observed in P3. This trend was also observed in the hot and cold spots differences in alcohol availability between hot and cold spots, which were drastically reduced between P2 and P3. This suggests that the relationship between the built environment and alcohol consumption, reported in previous studies [5,44], may have changed over time, potentially due to the effectiveness of alcohol control policies implemented in P2 and explicitly targeting the availability of alcohol (i.e., restricted alcohol sale hours and sale interdiction in gas stations and video stores) [6,17].

Finally, results also showed the complementarity of MGWR and local clustering. Indeed, geographic variation in the intercept of MGWR models corresponded to high and low alcohol consumption areas adjusted for the explanatory variables included in the model. This information supported the results of the local clustering analyses. Indeed, spatial patterns in the intercept coincided with the observed spatial clusters after adjustment for covariates. For example, the large hot spot identified in P3 corresponded to the area where the MGWR model showed a strong negative and significant relationship between higher education and alcohol consumption. This suggests that in that area where alcohol consumption is generally high, higher education is particularly protective against at-risk alcohol consumption.

### Strengths and limitations

First, our study uses data from a large population-based study over a long period and with a homogenous and dense geographical distribution across the study area. The spatial scales of analysis were objectively determined for spatial autocorrelation analysis and regression analyses. Our spatial clustering analyses relied on widely used spatial statistics. The global and local Moran’s I have been shown to be valid and robust indicators of spatial autocorrelation [30]. Additionally, our implementation of GWR and MGWR followed best practices for robust modeling of multiscale multivariate processes [31,34]. To the best of our knowledge, no existing studies have examined the spatial context of the determinants of alcohol consumption using MGWR. Such a geospatial approach was notably lacking in Switzerland, as most existing studies focused on US and UK populations [6,11]. Our study also aimed to address several methodological gaps reported in the literature [5]. We examined the relationship between alcohol physical availability and alcohol consumption using objective geographic measures of the built environment. Current and historical business listings were collected from a public source to avoid potential biases and missing data associated with business listings obtained from private companies. We used a street network-based approach recognized to capture better human mobility patterns than buffer or container approaches used in many previous studies [6]. We also examined both aggregated (i.e., on- and off-premises alcohol outlets) and disaggregated outlet categorizations to measure outlet density. This approach has been previously recommended to disentangle whether observed relationships apply to all outlet types or only certain ones [5]. Of note, our regression analyses showed no significant relationship between most disaggregated outlet categories and alcohol consumption, apart from bar density in P1. Additionally, our study focuses on the problem of average alcohol consumption, which has received little attention compared to acute alcohol consumption and related harms [45,46].

Several limitations of this study merit comment. First, the cross-sectional nature of the Bus Santé study precludes from evaluating the causality of observed relationships. Secondly, alcohol consumption was derived from self-reported FFQ data; we thus cannot exclude a social desirability bias. However, the same FFQ was used throughout the study period. The magnitude of the bias may have remained constant over time, allowing trends to be unaffected. Third, while recruitment methods of the Bus Santé study aimed at collecting information on a representative sample of the general population, participation bias may exist. Still, multiple strategies described in detail elsewhere have been introduced in the Bus Santé study recruitment to limit such bias [42]. Finally, we were only able to consider individuals’ exposure to alcohol outlets around the place of residence. Recent evidence suggests additional exposure outside of the residential neighborhood. Future research should ideally draw on travel surveys and GPS data to better capture individuals’ exposures. Nevertheless, exposure assessment is even complicated by increasing online alcohol advertising and sale with home delivery [5].

## Conclusions and policy implications

Local clustering of alcohol consumption was detected in three time periods (P1, 1993-1999; P2, 1999-2009; P3, 2009-2018) defined by the implementation of an array of alcohol control policies from 1999 to 2009. While decreasing substantially across periods, alcohol consumption in hot spots remained at inappropriate levels, particularly for women. Furthermore, the determinants of alcohol consumption changed over periods. Our findings show that specific socio-demographic and built environment characteristics are associated with increased alcohol consumption regardless of location (i.e., global scale). These may be the focus of population-wide programs and strategies. In contrast, other factors such as smoking status, nationality, being a woman, and higher education had more local relationships with alcohol consumption in the most recent period (2009-2018). By shedding light on the mix of spatial scales at which socio-demographic characteristics and alcohol availability influence alcohol consumption, our results may facilitate the development of improved, targeted public health interventions.

## Data Availability

All data produced in the present study are available upon reasonable request to the authors.

## Acknowledgments

The authors are grateful to the participants of the Bus Santé study.

## Bibliography

1. WHO. Global Status Report on Alcohol and Health 2018.; 2018.

2. Marmet S, Rehm J, Gmel G, Frick H, Gmel G. Alcohol-attributable mortality in Switzerland in 2011 - Age-specific causes of death and impact of heavy versus non-heavy drinking. Swiss Med Wkly. 2014;144(2122):13947. doi:10.4414/smw.2014.13947

3. OECD. Tackling Harmful Alcohol Use. OECD; 2015. doi:10.1787/9789264181069-en

4. Dumont S, Marques-Vidal P, Favrod-Coune T, et al. Alcohol policy changes and 22-year trends in individual alcohol consumption in a Swiss adult population: A 1993-2014 cross-sectional population-based study. BMJ Open. 2017;7(3). doi:10.1136/bmjopen-2016-014828

5. Holmes J, Guo Y, Maheswaran R, Nicholls J, Meier PS, Brennan A. The impact of spatial and temporal availability of alcohol on its consumption and related harms: A critical review in the context of UK licensing policies. Drug Alcohol Rev. 2014;33(5):515–525. doi:10.1111/dar.12191

6. Popova S, Giesbrecht N, Bekmuradov D, Patra J. Hours and days of sale and density of alcohol outlets: Impacts on alcohol consumption and damage: A systematic review. Alcohol Alcohol. 2009;44(5):500–516. doi:10.1093/alcalc/agp054

7. Campbell CA, Hahn RA, Elder R, et al. The Effectiveness of Limiting Alcohol Outlet Density As a Means of Reducing Excessive Alcohol Consumption and Alcohol-Related Harms. Am J Prev Med. 2009;37(6):556–569. doi:10.1016/j.amepre.2009.09.028

8. Picone GA, Sloan F, Trogdon JG. The effect of the tobacco settlement and smoking bans on alcohol consumption. Health Econ. 2004;13(10):1063–1080. doi:10.1002/hec.930

9. Sandoval JL, Leão T, Theler JM, et al. Alcohol control policies and socioeconomic inequalities in hazardous alcohol consumption: A 22-year cross-sectional study in a Swiss urban population. BMJ Open. 2019;9(5):e028971. doi:10.1136/bmjopen-2019-028971

10. Faeh D, Viswanathan B, Chiolero A, Warren W, Bovet P. Clustering of smoking, alcohol drinking and cannabis use in adolescents in a rapidly developing country. BMC Public Health. 2006;6(1):169. doi:10.1186/1471-2458-6-169

11. Duncan DT, Rienti M, Kulldorff M, et al. Local spatial clustering in youths’ use of tobacco, alcohol, and marijuana in Boston. Am J Drug Alcohol Abuse. 2016;42(4):412–421. doi:10.3109/00952990.2016.1151522

12. Assanangkornchai S, Li J, McNeil E, Saingam D. Clusters of alcohol and drug use and other health-risk behaviors among Thai secondary school students: A latent class analysis. BMC Public Health. 2018;18(1):1272. doi:10.1186/s12889-018-6205-z

13. Gruenewald PJ, Remer LG, Lascala EA. Testing a social ecological model of alcohol use: The California 50-city study. Addiction. 2014;109(5):736–745. doi:10.1111/add.12438

14. Bryden A, Roberts B, McKee M, Petticrew M. A systematic review of the influence on alcohol use of community level availability and marketing of alcohol. Heal Place. 2012;18(2):349–357. doi:10.1016/j.healthplace.2011.11.003

15. Bryden A, Roberts B, Petticrew M, McKee M. A systematic review of the influence of community level social factors on alcohol use. Heal Place. 2013;21:70–85. doi:10.1016/j.healthplace.2013.01.012

16. Collins SE. Associations between socioeconomic factors and alcohol outcomes. Alcohol Res Curr Rev. 2016;38(1):83–94.

17. Sherk A, Stockwell T, Chikritzhs T, et al. Alcohol consumption and the physical availability of take-away alcohol: Systematic reviews and meta-analyses of the days and hours of sale and outlet density. J Stud Alcohol Drugs. 2018;79(1):58–67. doi:10.15288/jsad.2018.79.58

18. Chaix B, Merlo J, Chauvin P. Comparison of a spatial approach with the multilevel approach for investigating place effects on health: The example of healthcare utilisation in France. J Epidemiol Community Health. 2005;59(6):517–526. doi:10.1136/jech.2004.025478

19. Guessous I, Bochud M, Theler JM, Gaspoz JM, Pechère-Bertschi A. 1999-2009 trends in prevalence, unawareness, treatment and control of hypertension in Geneva, Switzerland. Barengo NC, ed. PLoS One. 2012;7(6):e39877. doi:10.1371/journal.pone.0039877

20. Morabia A, Bernstein M, Héritier S, Ylli A. Community-based surveillance of cardiovascular risk factors in Geneva: Methods, resulting distributions, and comparisons with other populations. Prev Med (Baltim). 1997;26(3):311–319. doi:10.1006/pmed.1997.0146

21. Openshaw S. The Modifiable Areal Unit Problem. Vol 38. Geo Books; 1983.

22. Morabia A, Bernstein M, Kumanyika S, et al. Développement et validation d’un questionnaire alimentaire semi-quantitatif à partir d’une enquête de population. Sozial-und Prä ventivmedizin SPM. 1994;39(6):345–369. doi:10.1007/BF01299666

23. Mozaffarian D, Fahimi S, Singh GM, et al. Global Sodium Consumption and Death from Cardiovascular Causes. N Engl J Med. 2014;371(7):624–634. doi:10.1056/nejmoa1304127

24. Micha R, Khatibzadeh S, Shi P, et al. Global, regional, and national consumption levels of dietary fats and oils in 1990 and 2010: A systematic analysis including 266 country-specific nutrition surveys. BMJ. 2014;348. doi:10.1136/bmj.g2272

25. REG. Accessed January 7, 2021. https://www.ge.ch/utiliser-repertoire-entreprises-jreg/consulter-reg

26. SITG. Catalogue | SITG. Accessed January 7, 2021. https://ge.ch/sitg/sitg_catalog/sitg_donnees

27. Boeing G. OSMnx: New methods for acquiring, constructing, analyzing, and visualizing complex street networks. Comput Environ Urban Syst. 2017;65:126–139. doi:10.1016/j.compenvurbsys.2017.05.004

28. Foti F, Waddell P, Luxen D. A Generalized Computational Framework for Accessibility□: From the Pedestrian to the Metropolitan Scale.; 2012.

29. Buehler R, Pucher J, Merom D, Bauman A. Active travel in Germany and the U.S.: Contributions of daily walking and cycling to physical activity. Am J Prev Med. 2011;41(3):241–250. doi:10.1016/j.amepre.2011.04.012

30. Auchincloss AH, Gebreab SY, Mair C, Diez Roux AV. A review of spatial methods in epidemiology, 2000-2010. Annu Rev Public Health. 2012;33(1):107–122. doi:10.1146/annurev-publhealth-031811-124655

31. Oshan TM, Li Z, Kang W, Wolf LJ, Stewart Fotheringham A. MGWR: A python implementation of multiscale geographically weighted regression for investigating process spatial heterogeneity and scale. ISPRS Int J Geo-Information. 2019;8(6). doi:10.3390/ijgi8060269

32. Chen DR, Truong K. Using multilevel modeling and geographically weighted regression to identify spatial variations in the relationship between place-level disadvantages and obesity in Taiwan. Appl Geogr. 2012;32(2):737–745. doi:10.1016/j.apgeog.2011.07.018

33. Chi SH, Grigsby-Toussaint DS, Bradford N, Choi J. Can Geographically Weighted Regression improve our contextual understanding of obesity in the US? Findings from the USDA Food Atlas. Appl Geogr. 2013;44:134–142. doi:10.1016/j.apgeog.2013.07.017

34. Oshan TM, Smith JP, Fotheringham AS. Targeting the spatial context of obesity determinants via multiscale geographically weighted regression. Int J Health Geogr. 2020;19(1):11. doi:10.1186/s12942-020-00204-6

35. Diggle P. Applied Spatial Statistics for Public Health Data. Vol 100. John Wiley & Sons, Inc.; 2005. doi:10.1198/jasa.2005.s15

36. Anselin L. Local Indicators of Spatial Association—LISA. Geogr Anal. 1995;27(2):93–115. doi:10.1111/j.1538-4632.1995.tb00338.x

37. Consommation d’alcool à risque. Accessed May 4, 2021. https://www.bag.admin.ch/bag/fr/home/gesund-leben/sucht-und-gesundheit/alkohol/problemkonsum.html

38. Tatem AJ. Innovation to impact in spatial epidemiology. BMC Med. 2018;16(1):209. doi:10.1186/s12916-018-1205-5

39. Fu SH, Jha P, Gupta PC, Kumar R, Dikshit R, Sinha D. Geospatial analysis on the distributions of tobacco smoking and alcohol drinking in India. PLoS One. 2014;9(7). doi:10.1371/journal.pone.0102416

40. Guessous I, Joost S, Jeannot E, Theler JM, Mahler P, Gaspoz JM. A comparison of the spatial dependence of body mass index among adults and children in a Swiss general population. Nutr Diabetes. 2014;4(3):e111–e111. doi:10.1038/nutd.2014.8

41. Joost S, De Ridder D, Marques-Vidal P, et al. Overlapping spatial clusters of sugar-sweetened beverage intake and body mass index in Geneva state, Switzerland. Nutr Diabetes. 2019;9(1):1–10. doi:10.1038/s41387-019-0102-0

42. De Ridder D, Belle FN, Marques□vidal P, et al. Geospatial analysis of sodium and potassium intake: A swiss population□based study. Nutrients. 2021;13(6):1798. doi:10.3390/nu13061798

43. Huang GC, Soto D, Fujimoto K, Valente TW. The interplay of friendship networks and social networking sites: Longitudinal analysis of selection and influence effects on adolescent smoking and alcohol use. Am J Public Health. 2014;104(8). doi:10.2105/AJPH.2014.302038

44. Lo CC, Weber J, Cheng TC. A spatial analysis of student binge drinking, alcohol-outlet density, and social disadvantages. Am J Addict. 2013;22(4):391–401. doi:10.1111/j.1521-0391.2013.12022.x

45. Zhu L, Gorman DM, Horel S. Alcohol outlet density and violence: A geospatial analysis. Alcohol Alcohol. 2004;39(4):369–375. doi:10.1093/alcalc/agh062

46. Ying YH, Wu CC, Chang K. The effectiveness of drinking and driving policies for different alcohol-related fatalities: A quantile regression analysis. Int J Environ Res Public Health. 2013;10(10):4628–4644. doi:10.3390/ijerph10104628

