## Supplementary material for "Evolution of the spatial distribution of alcohol consumption following alcohol control policies: a 25-year cross-sectional study in a Swiss urban population"

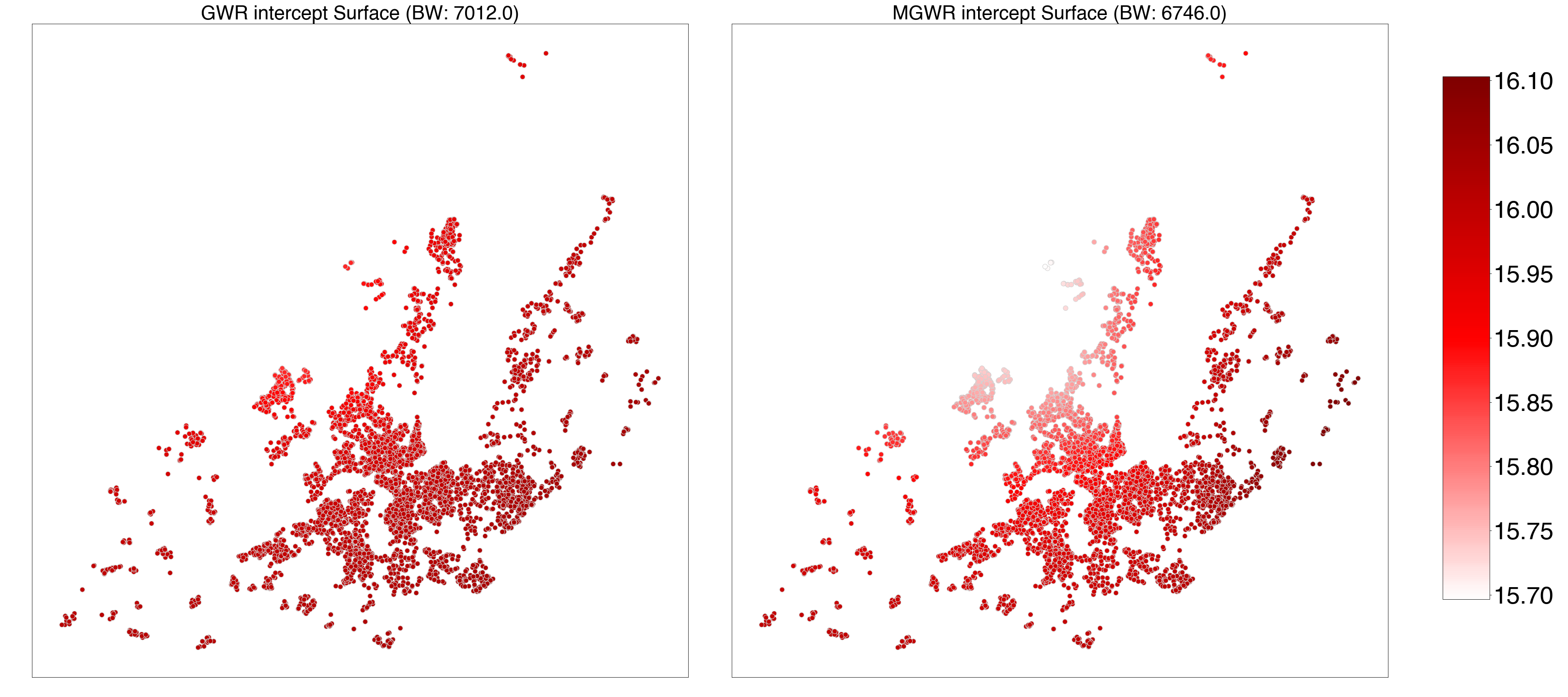

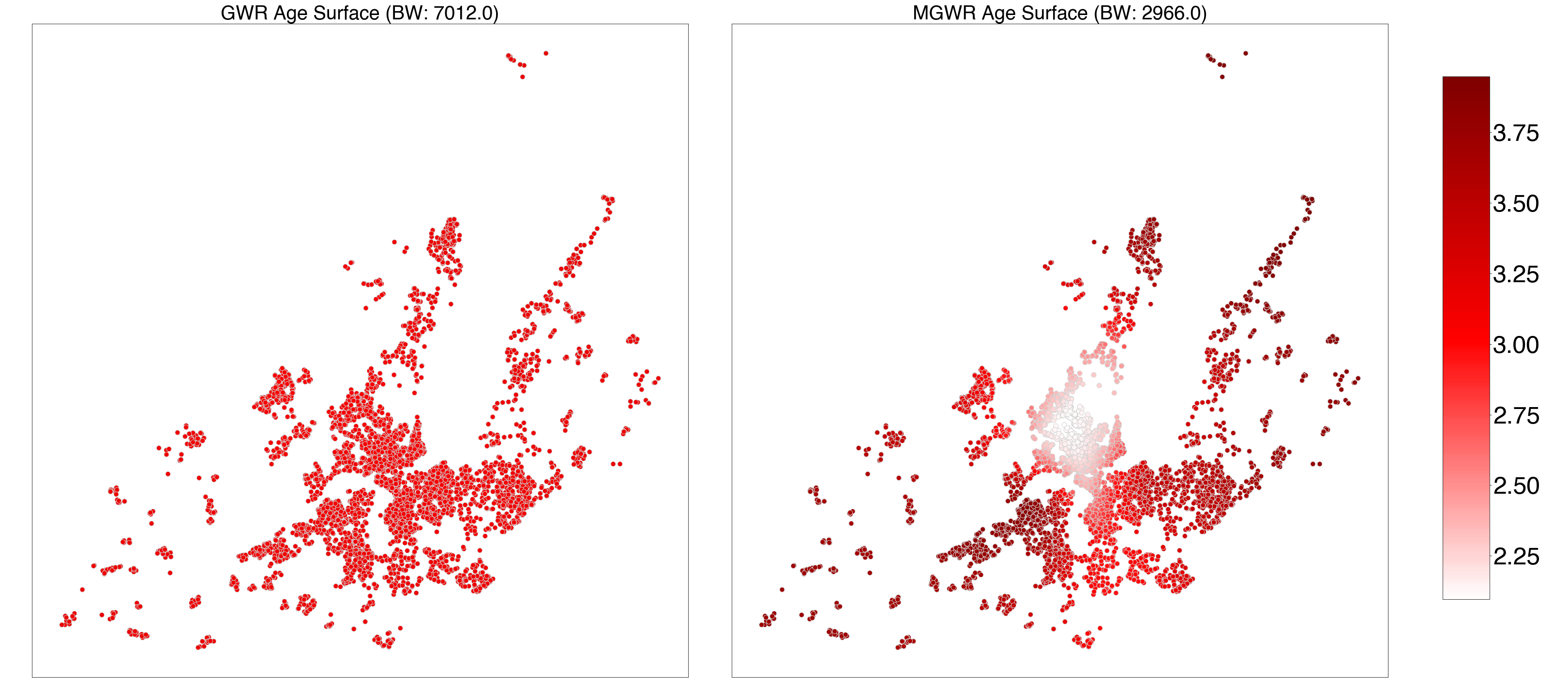

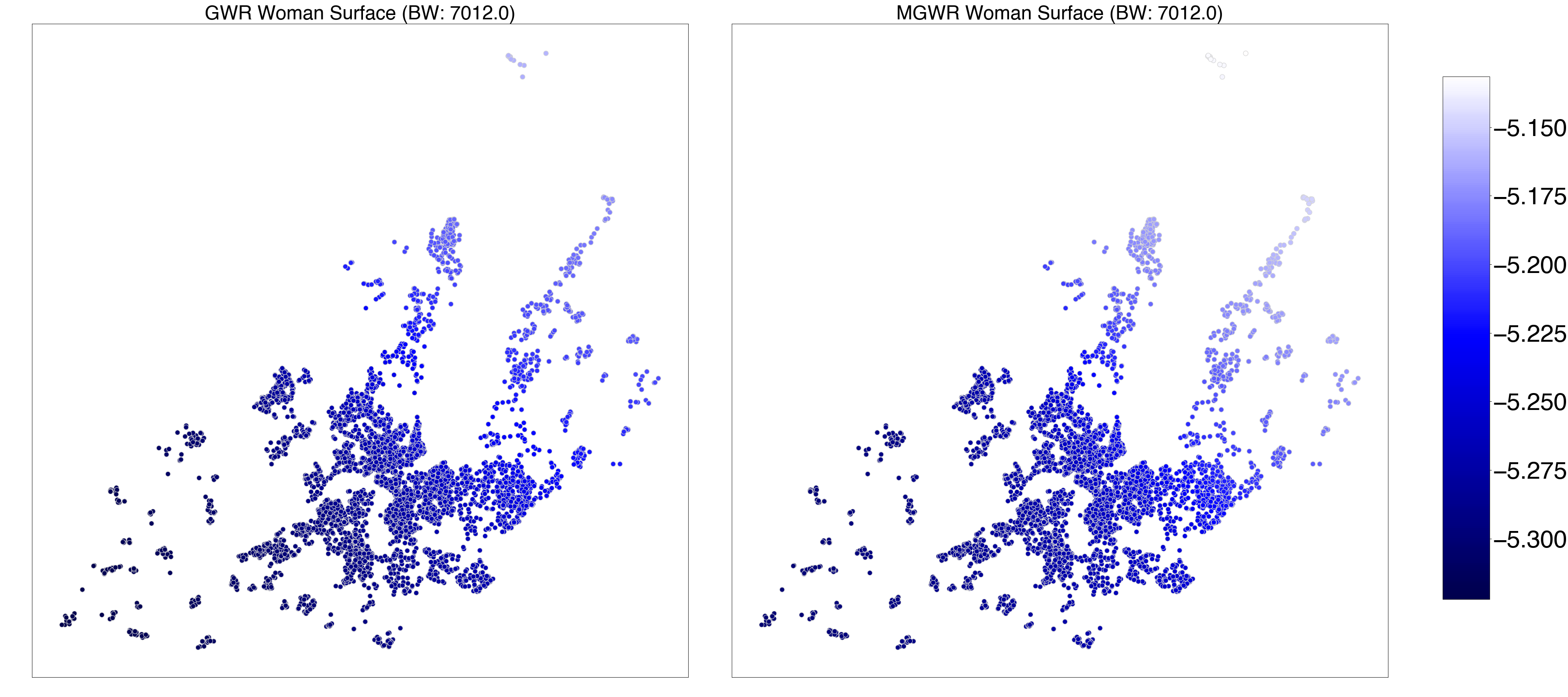

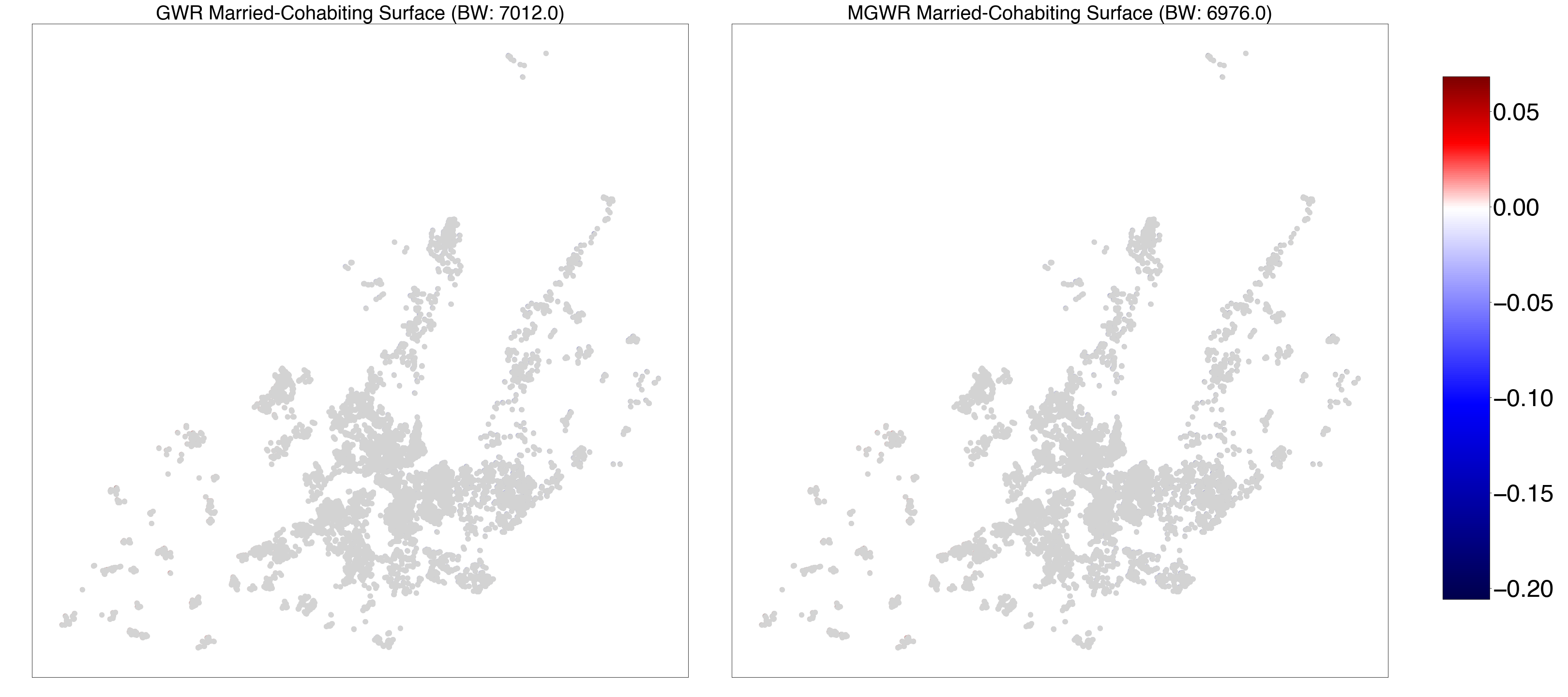

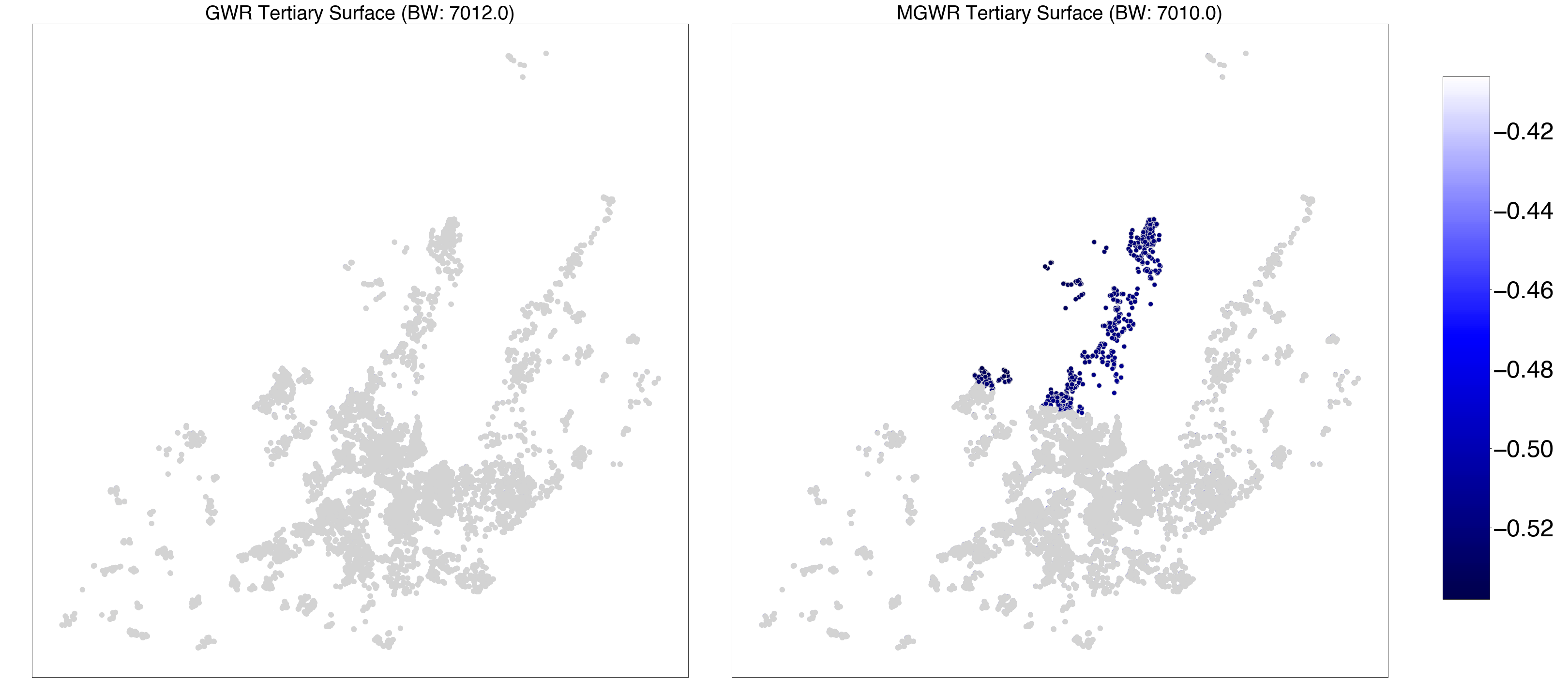

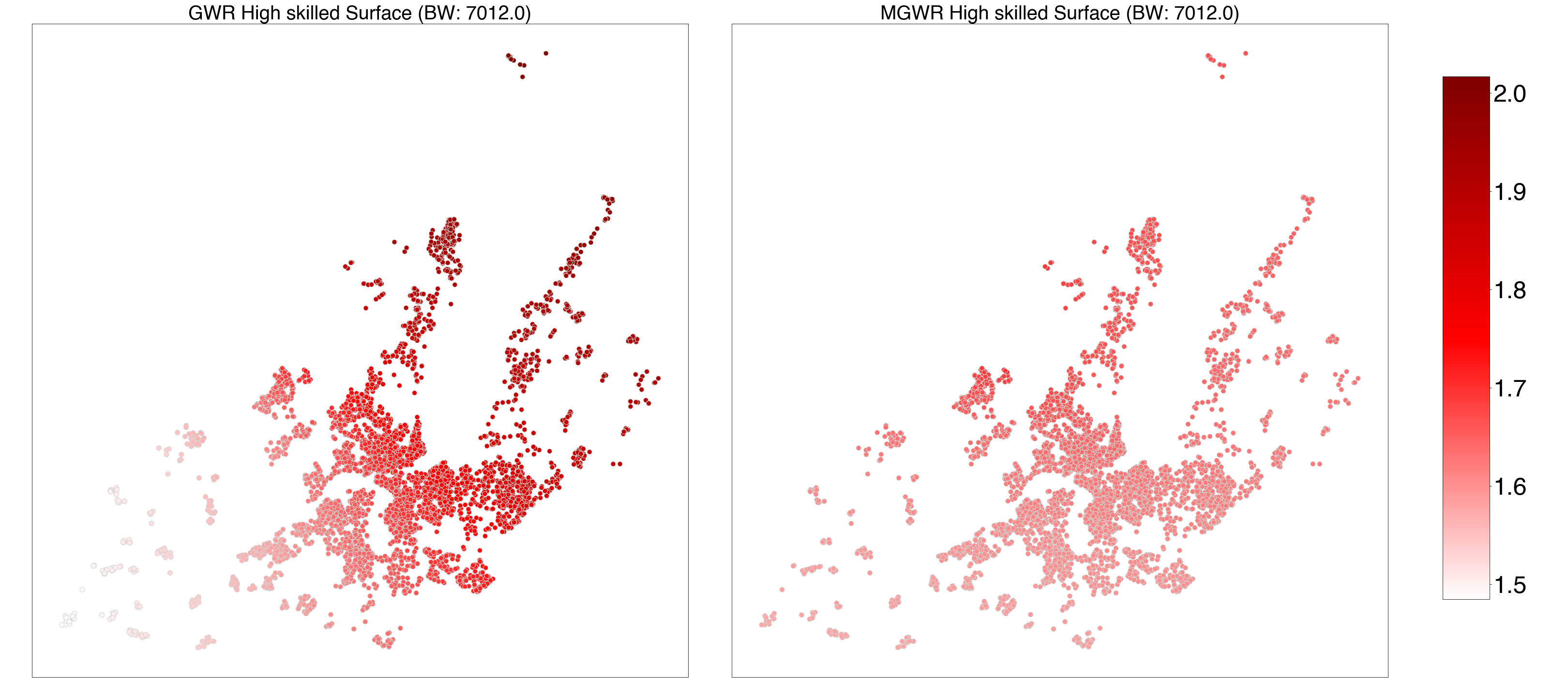

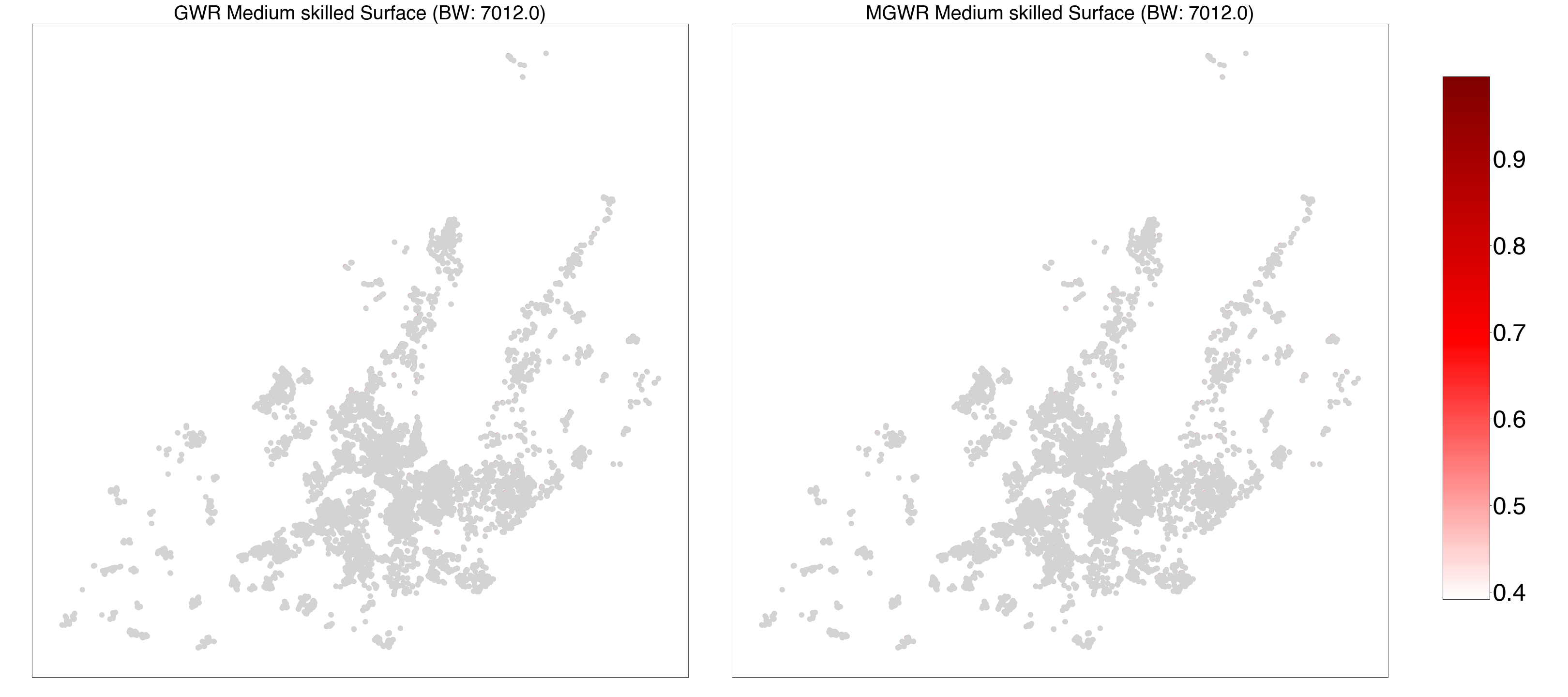

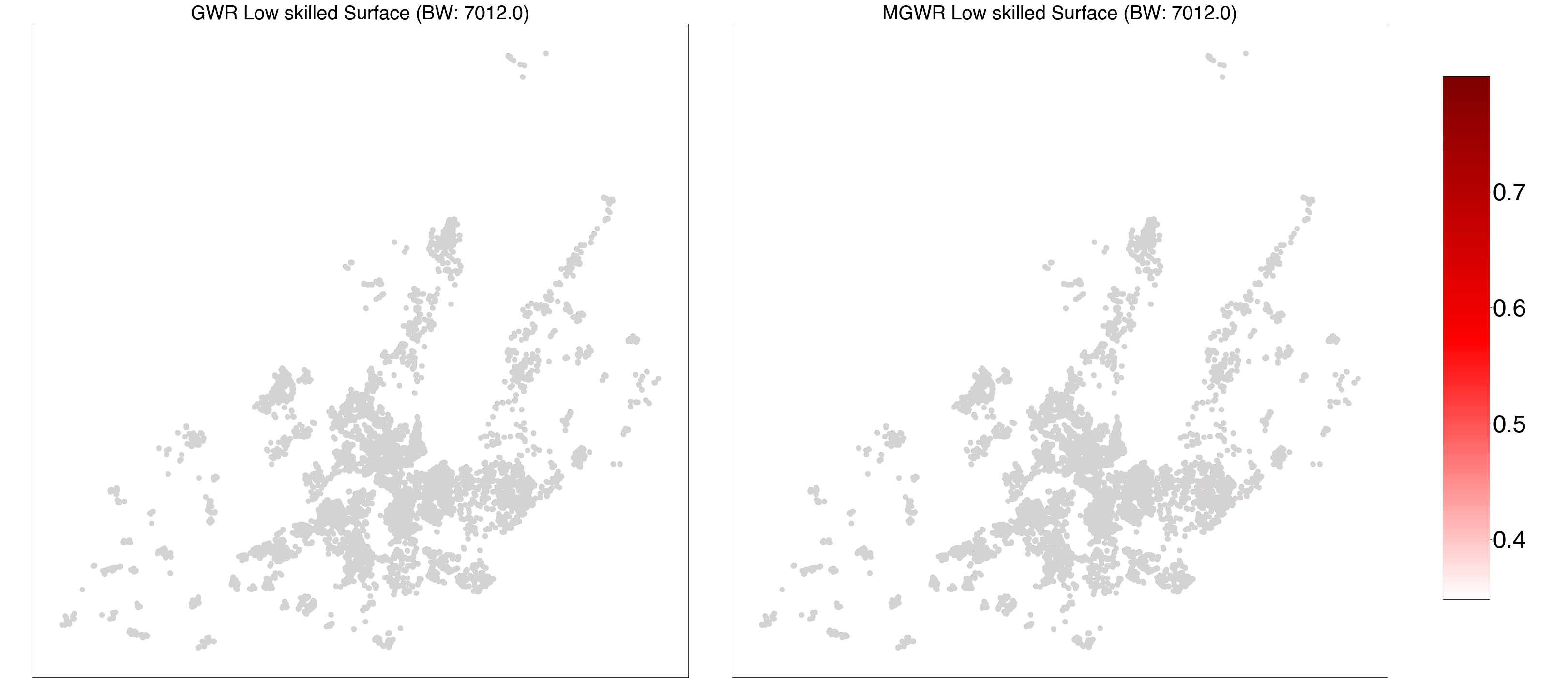

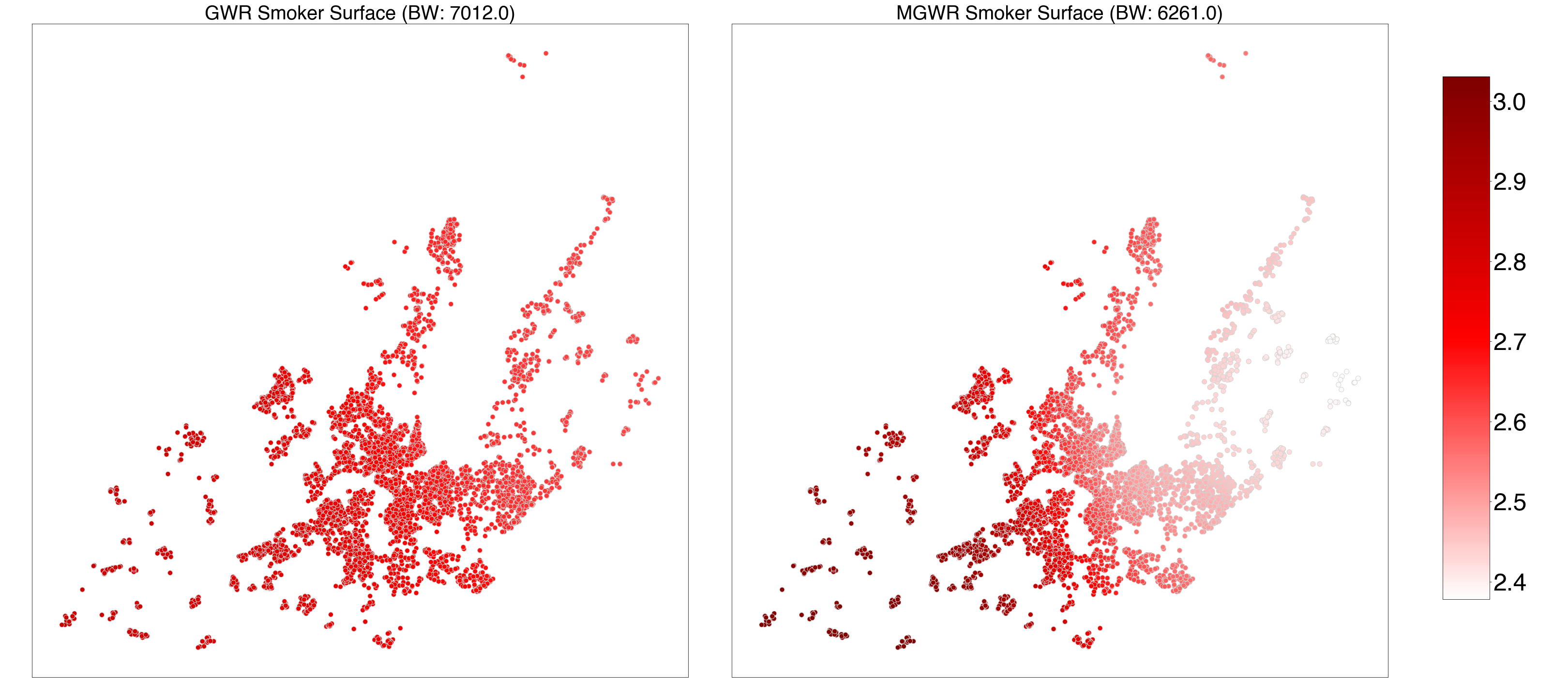

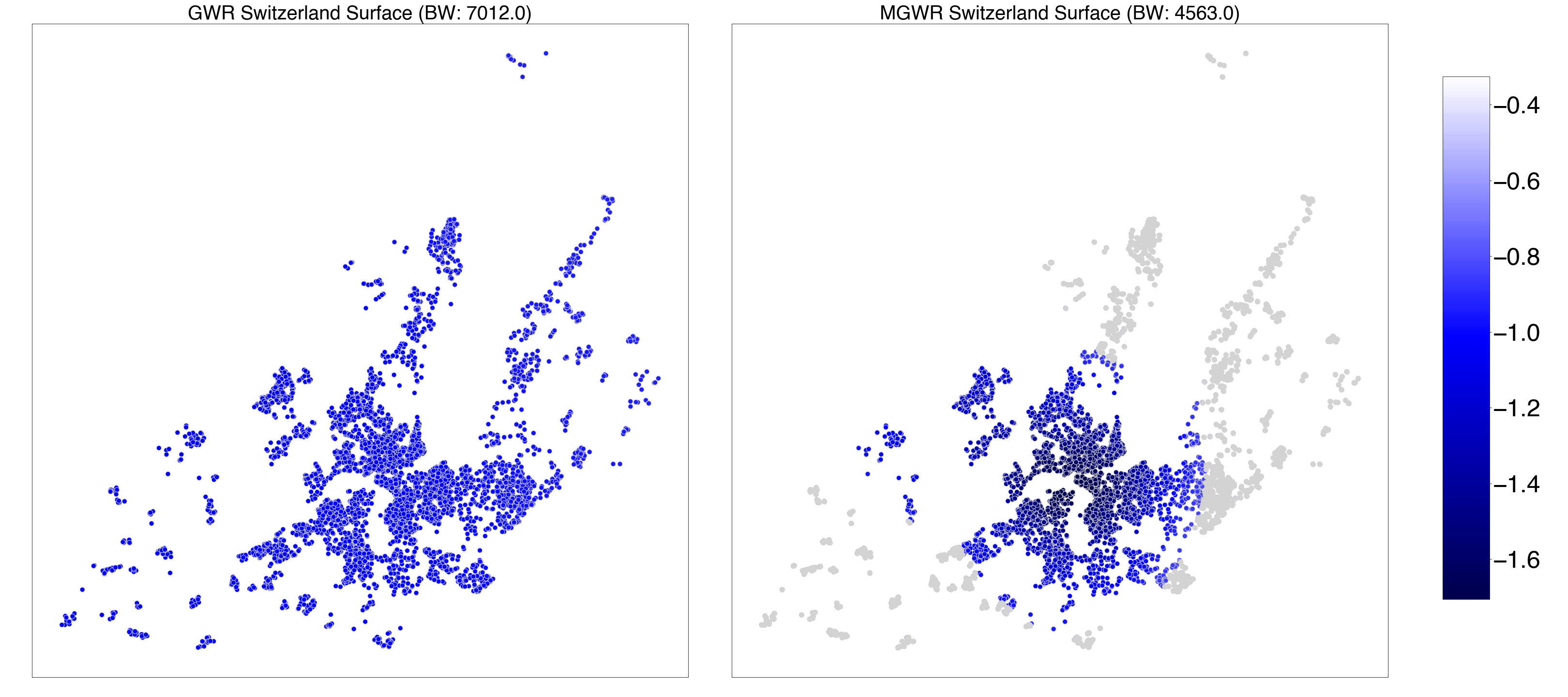

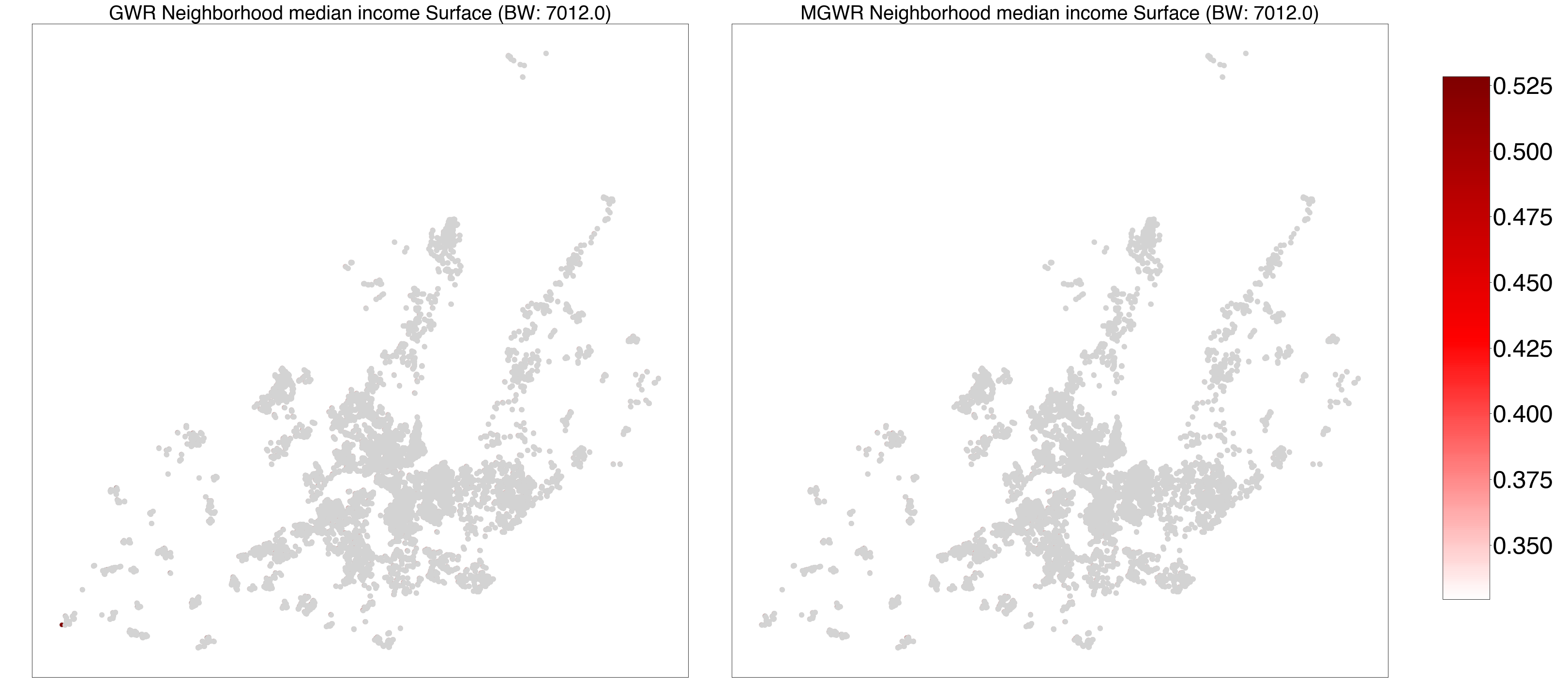

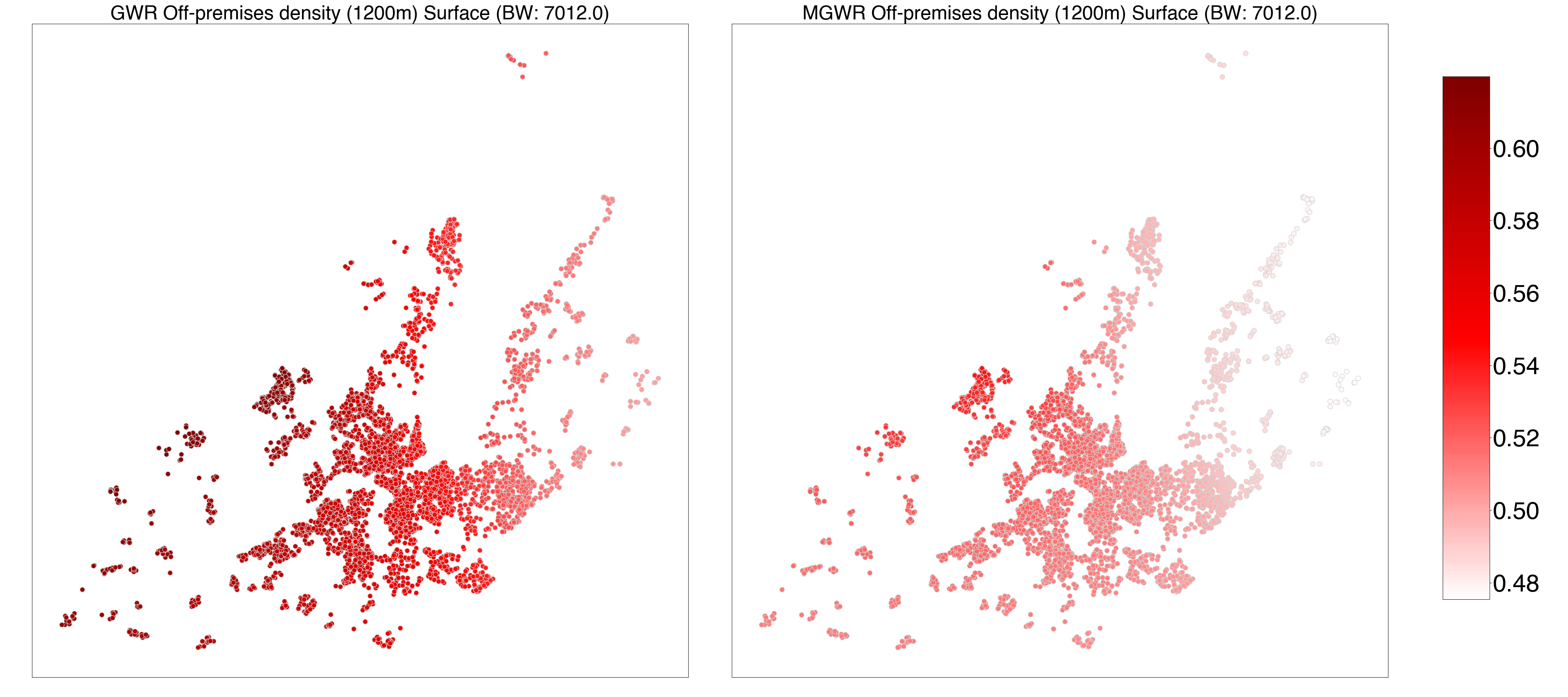

***Figure S1*** *-* ***GWR and MGWR parameter estimate surfaces for period 1 (P1)—1993.01.01-1999.06.30.*** *Composite maps for geographically weighted regression (GWR) (left) and multiscale GWR (MGWR) (right) parameter estimate surfaces for intercept, age, gender, married-cohabiting, tertiary education, high-, medium-, low-occupational level, smoking status, nationality, neighborhood median household income and off-premises outlet density.*

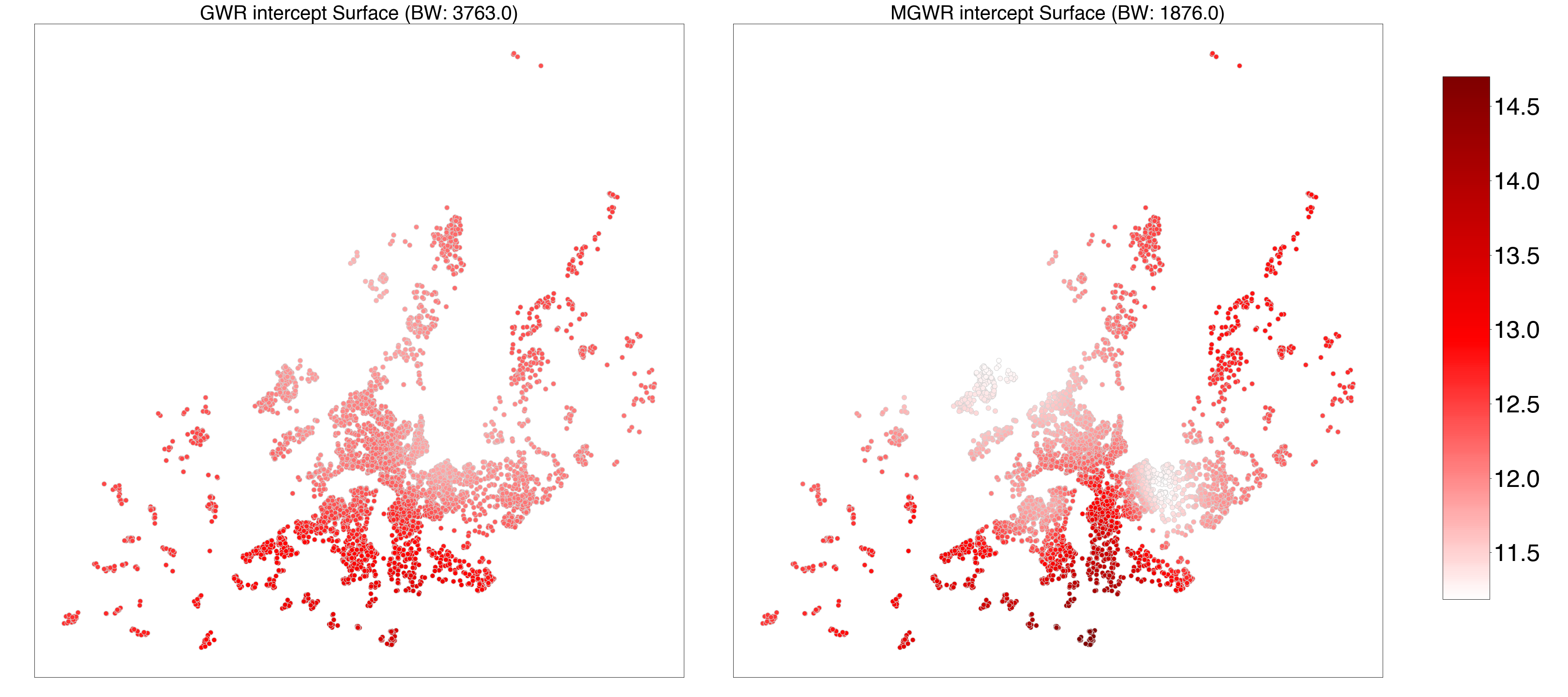

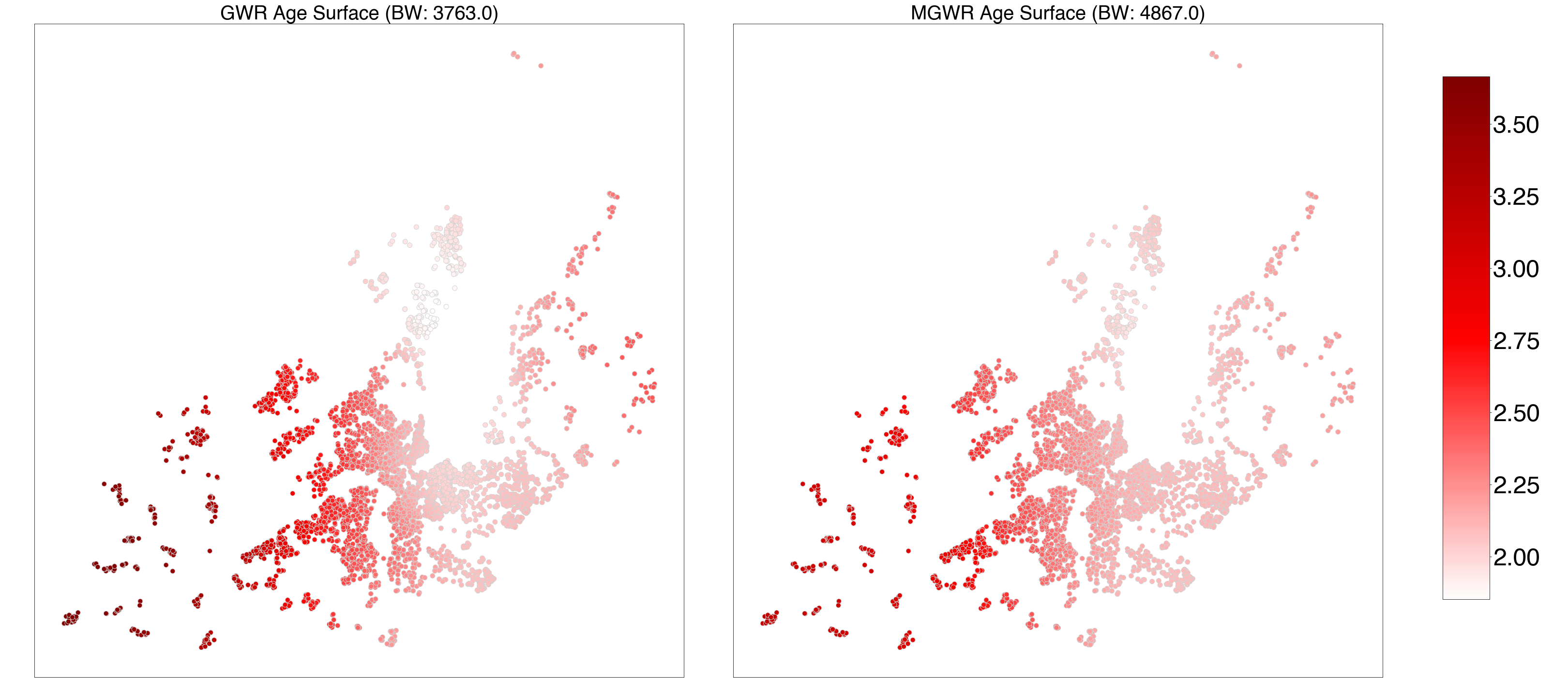

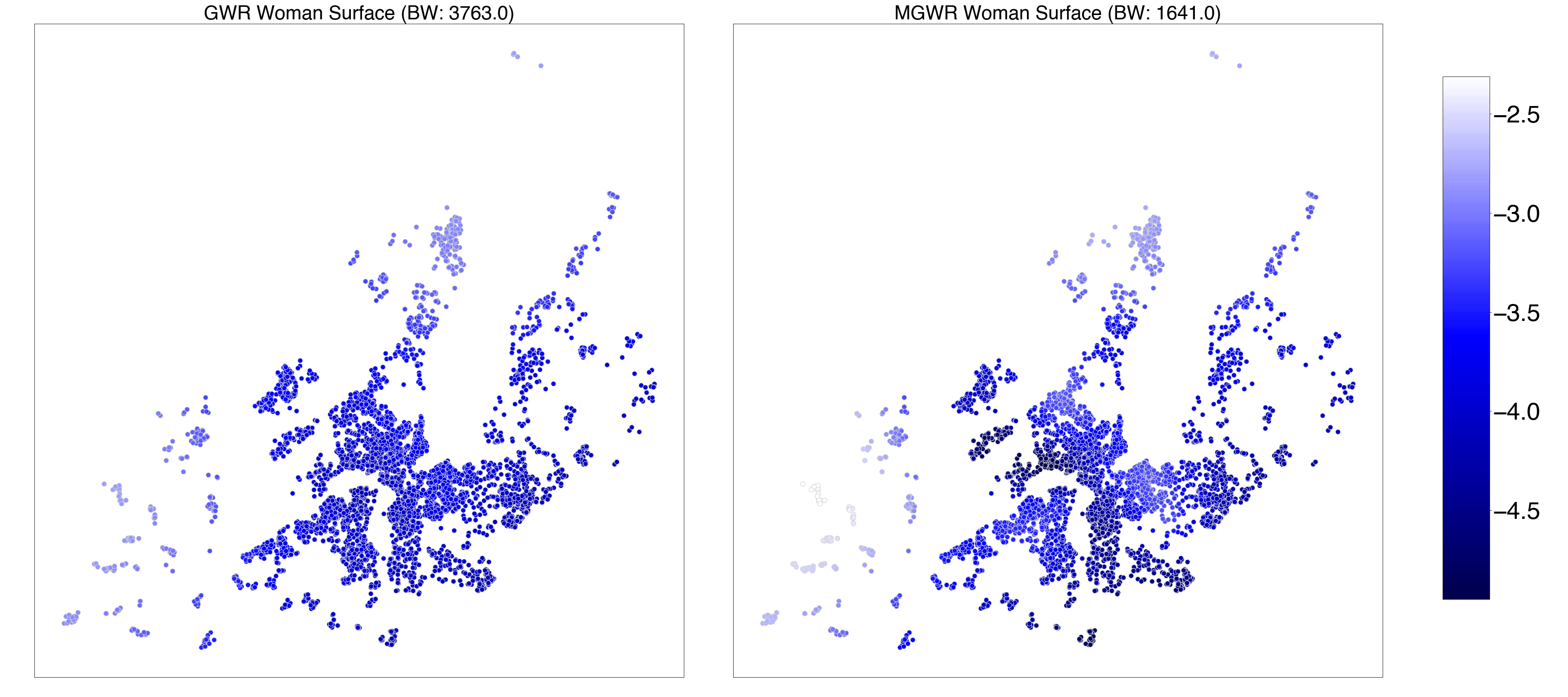

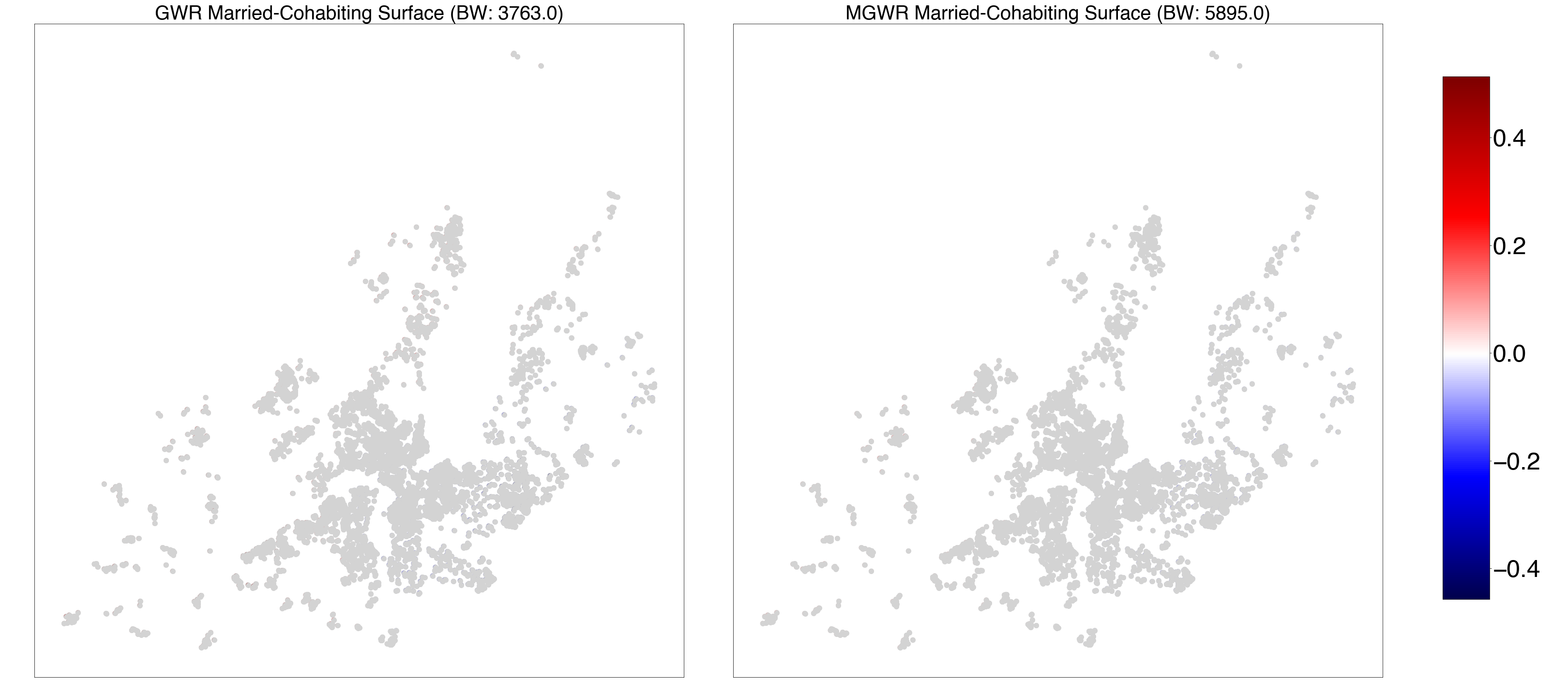

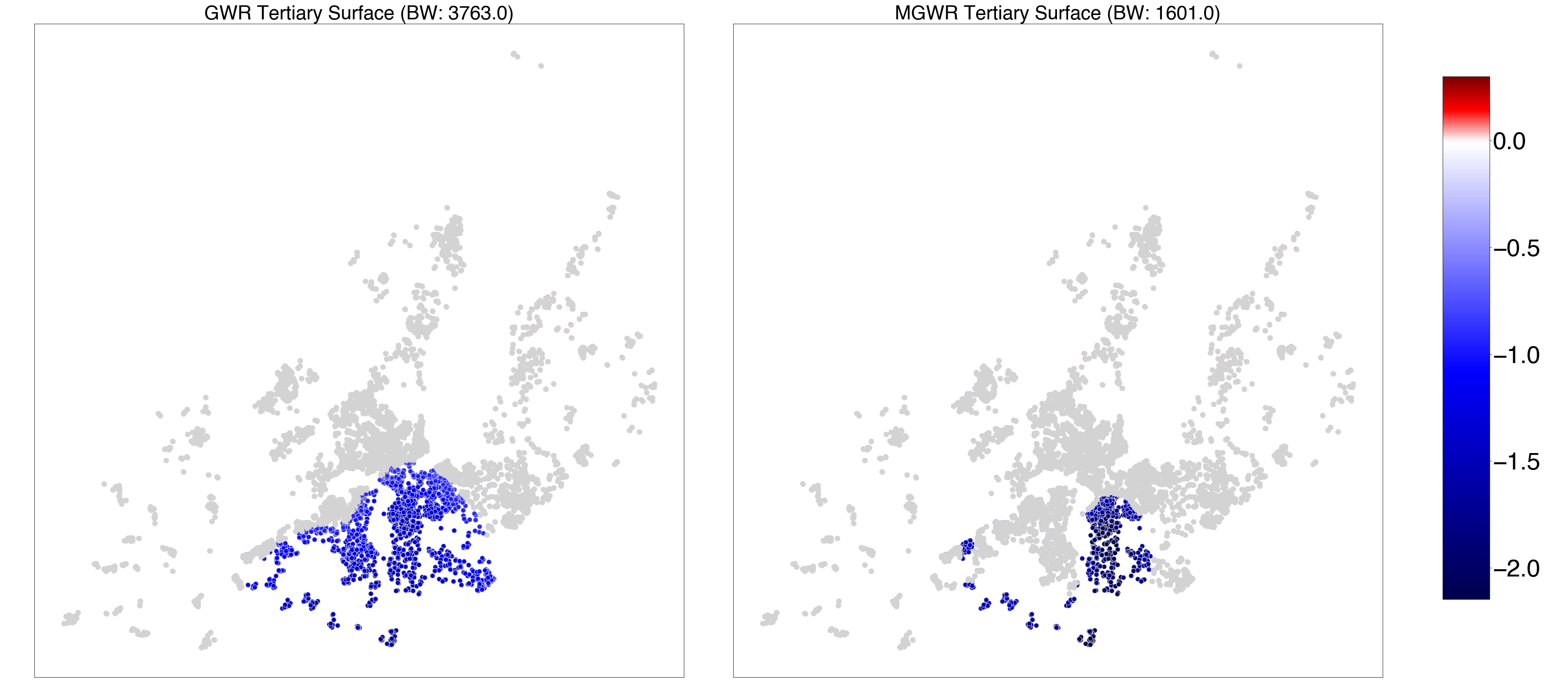

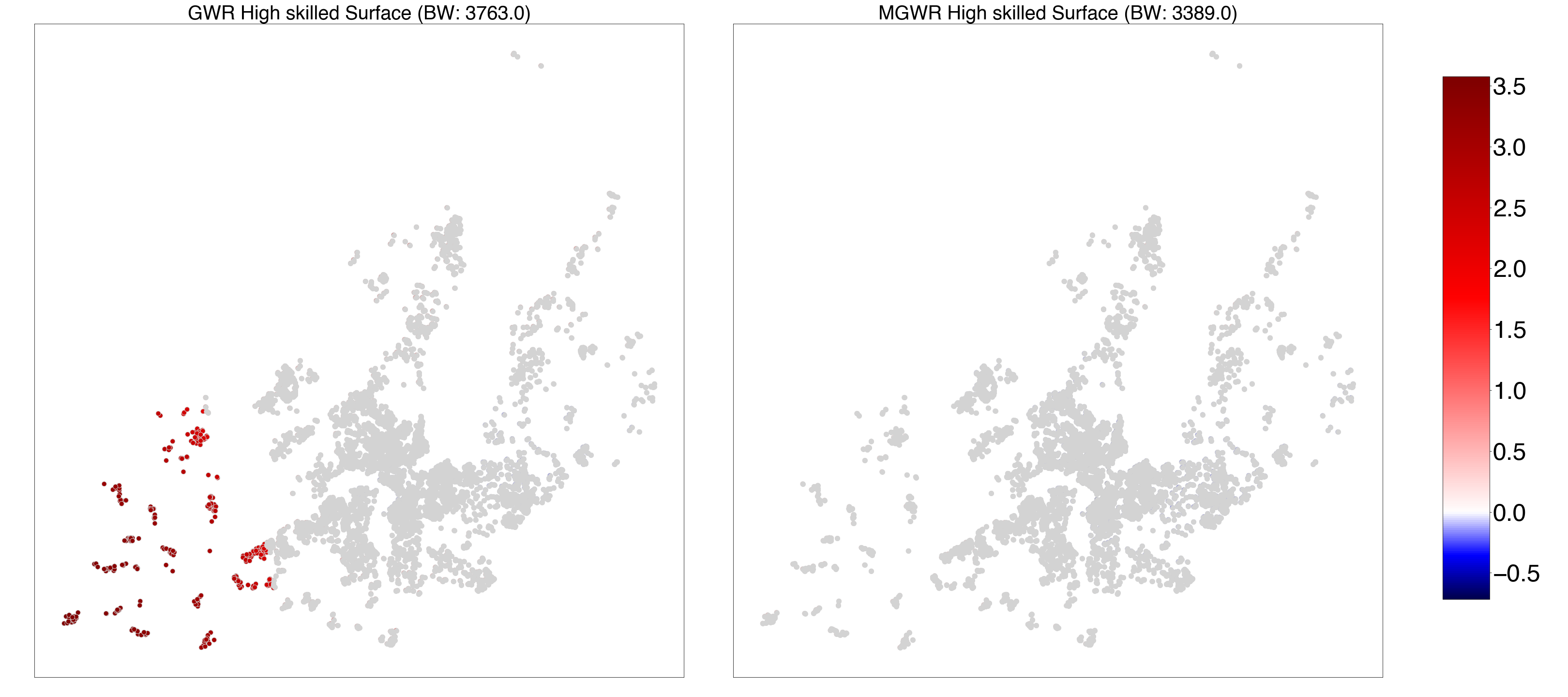

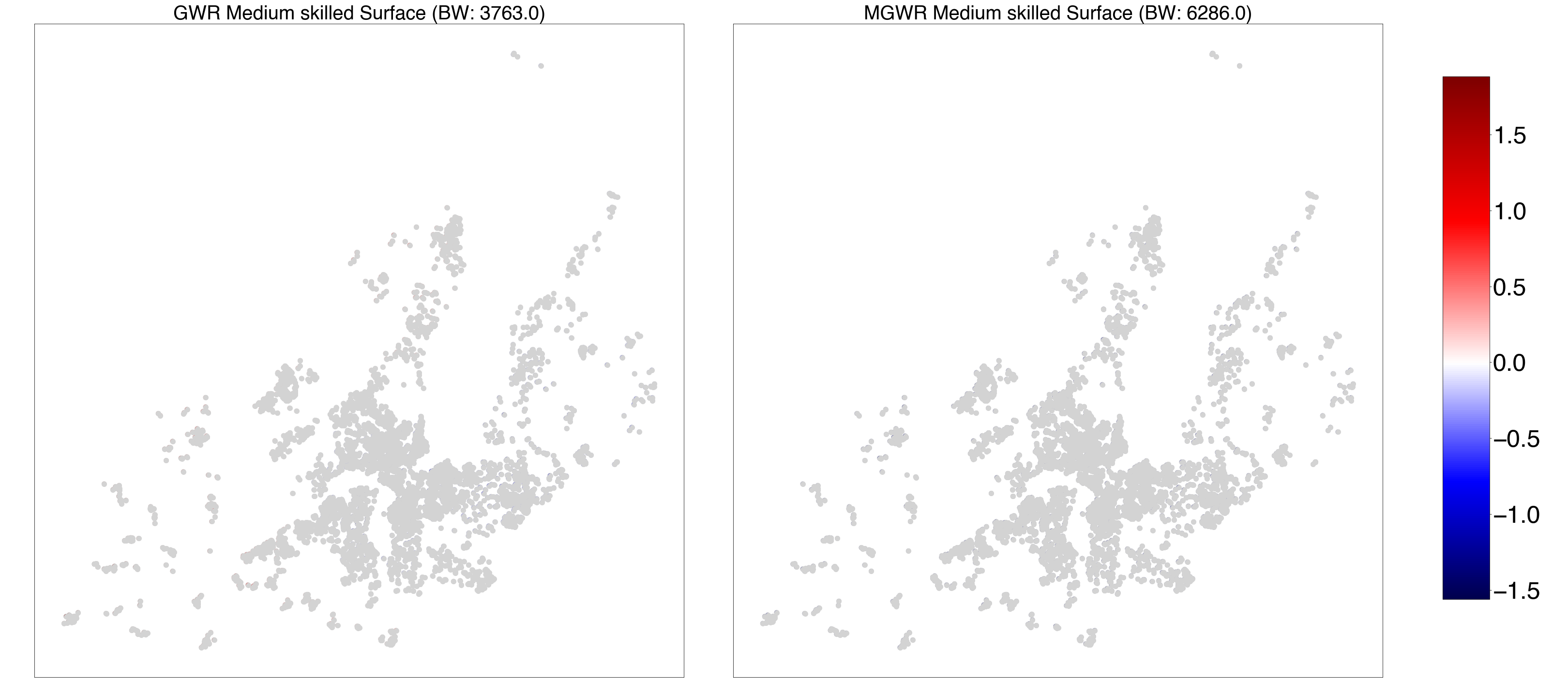

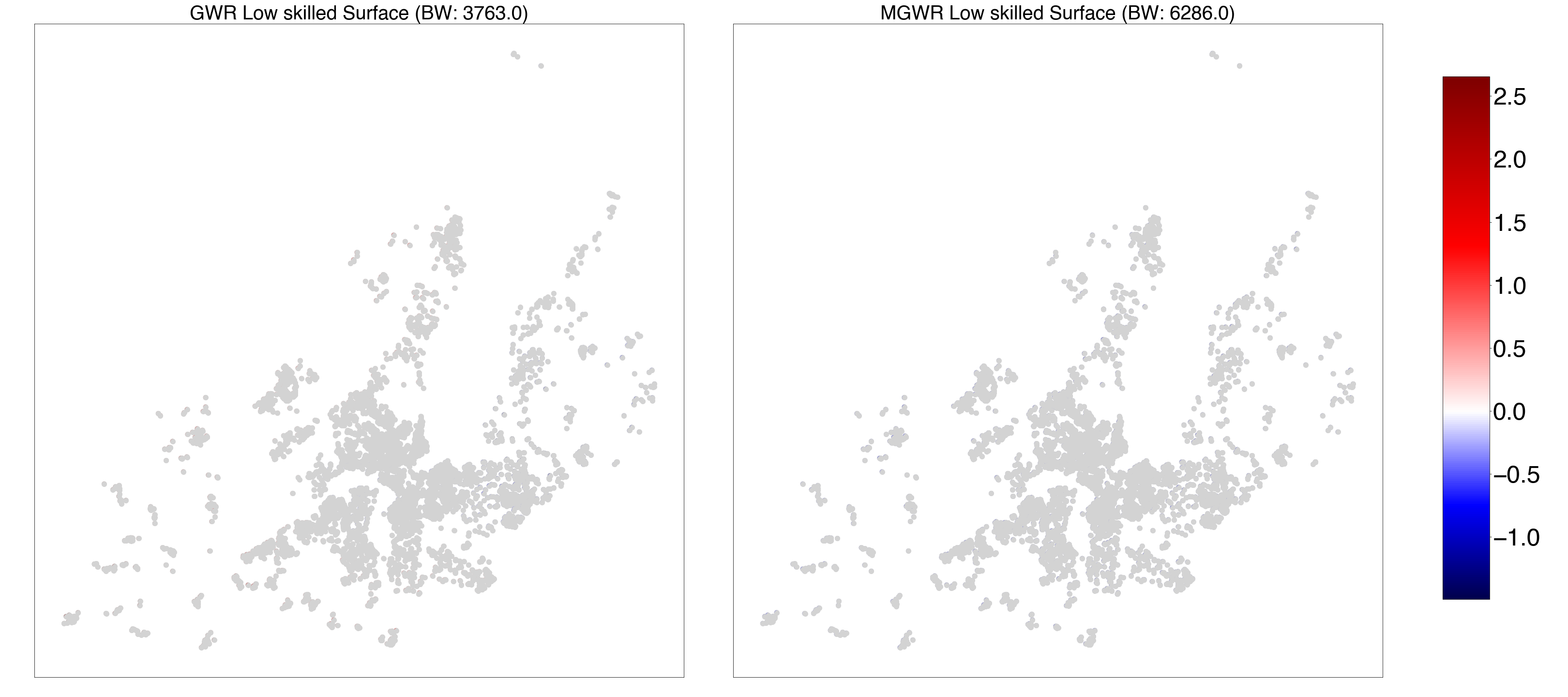

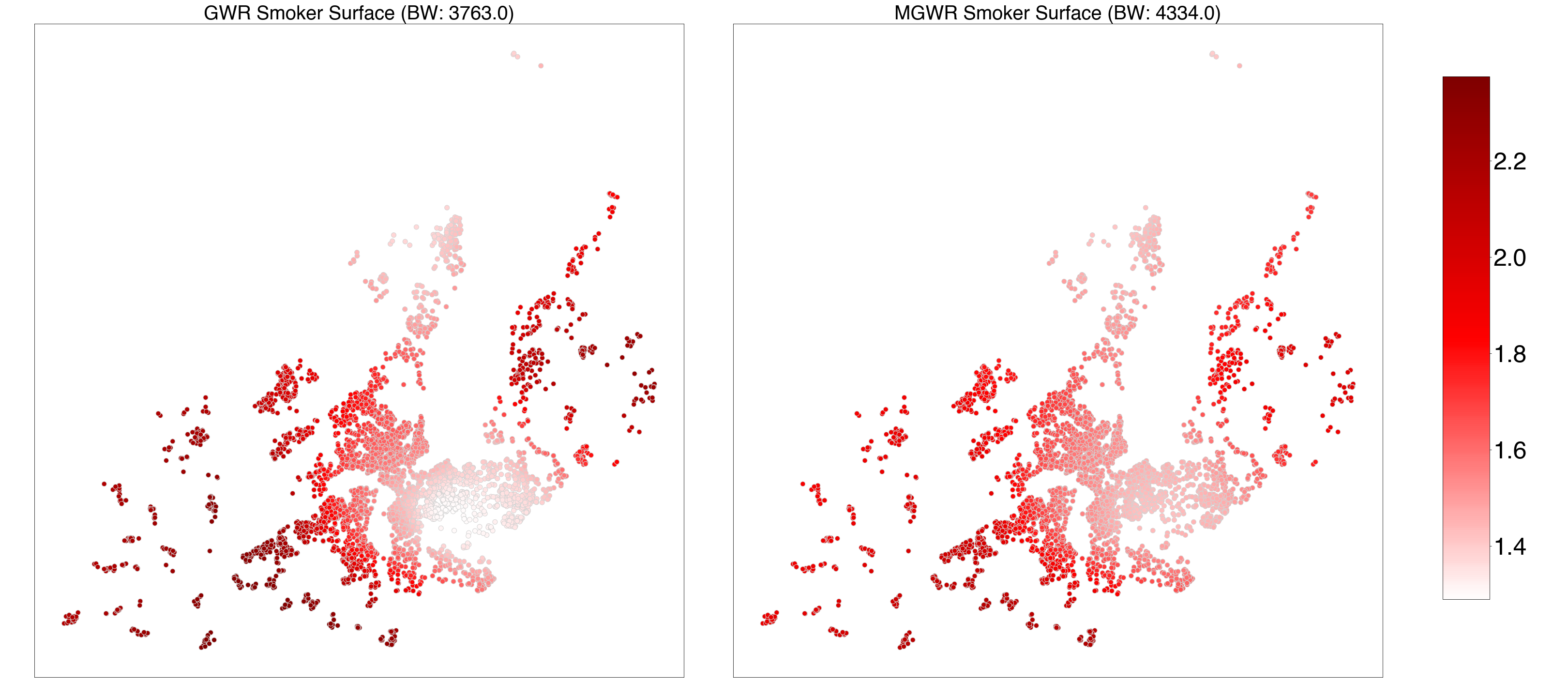

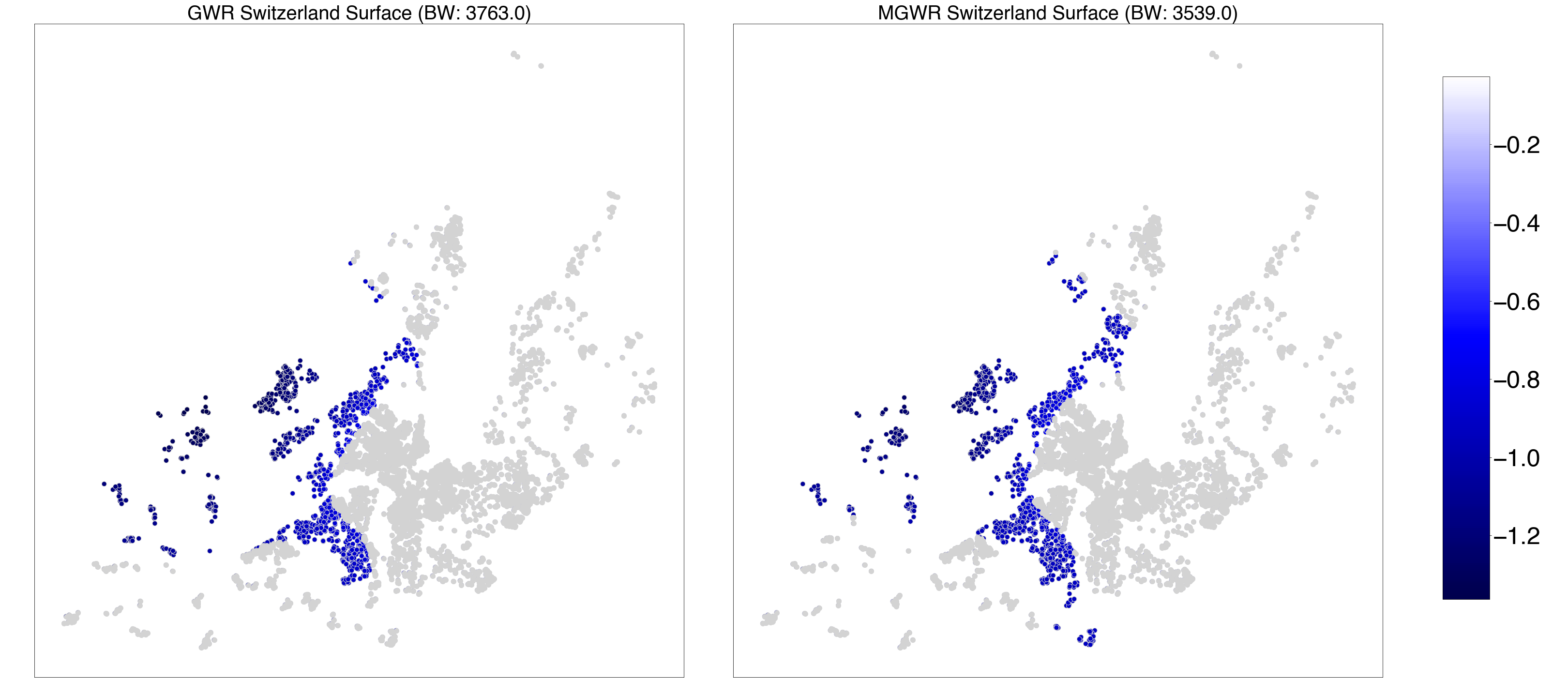

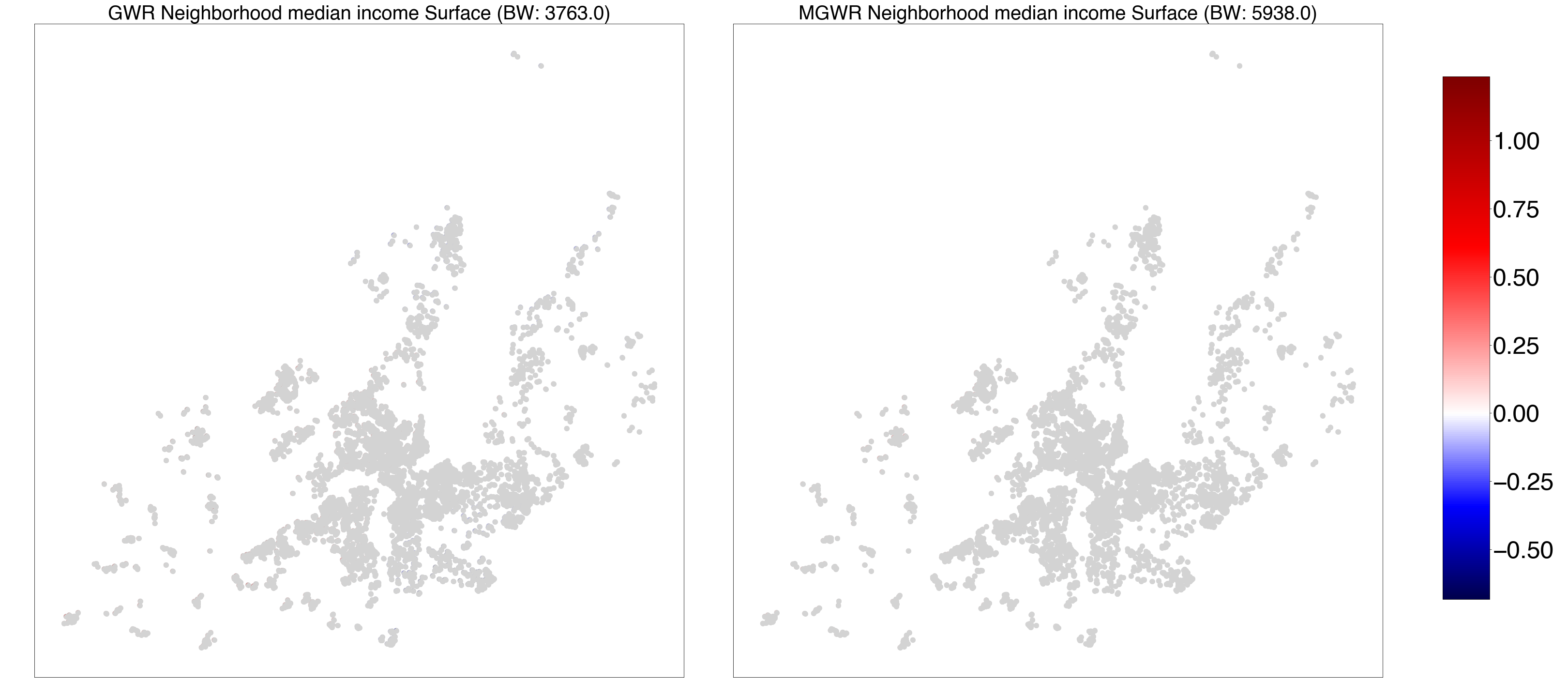

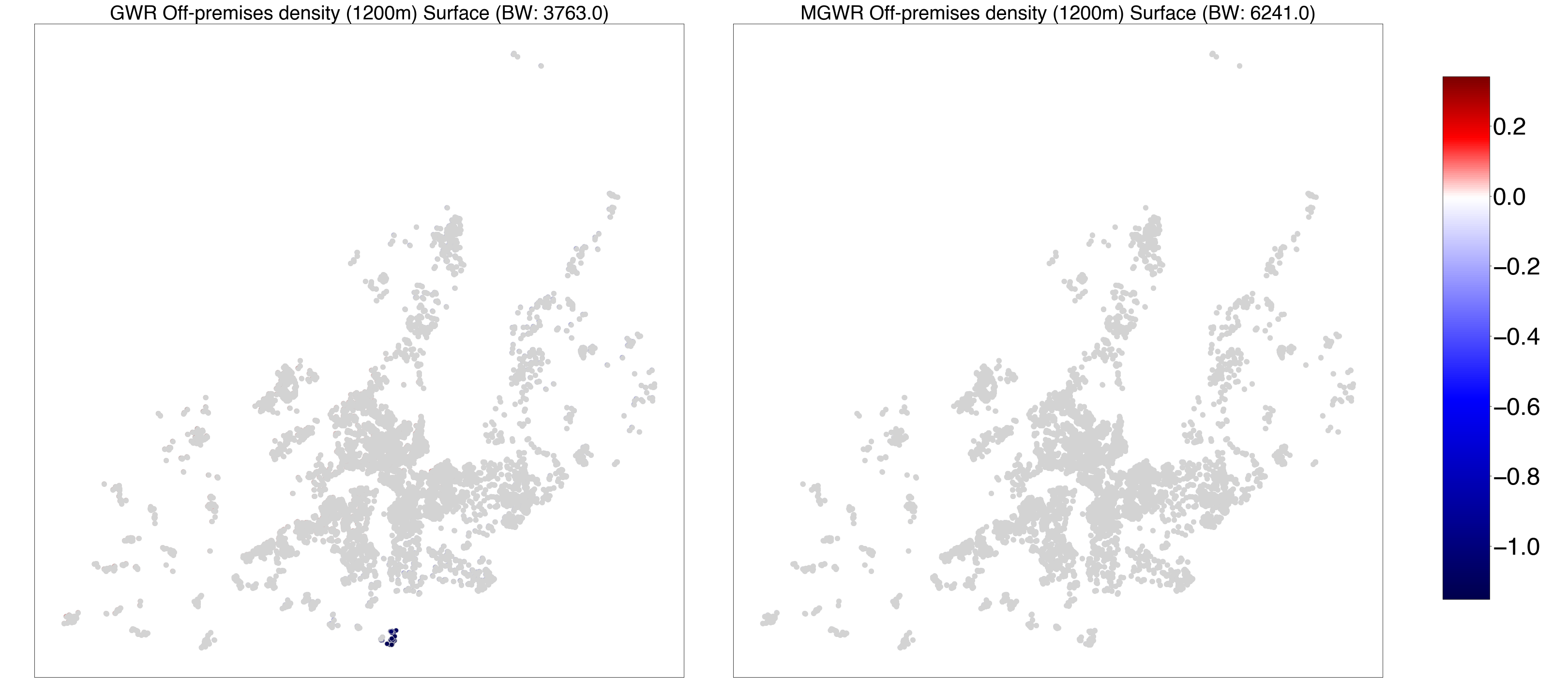

***Figure S2*** *-* ***GWR and MGWR parameter estimate surfaces for period 2 (P2)—1999.07.01-2009.10.31.*** *Composite maps for geographically weighted regression (GWR) (left) and multiscale GWR (MGWR) (right) parameter estimate surfaces for intercept, age, gender, married-cohabiting, tertiary education, high-, medium-, low-occupational level, smoking status, nationality, neighborhood median household income and off-premises outlet density.*

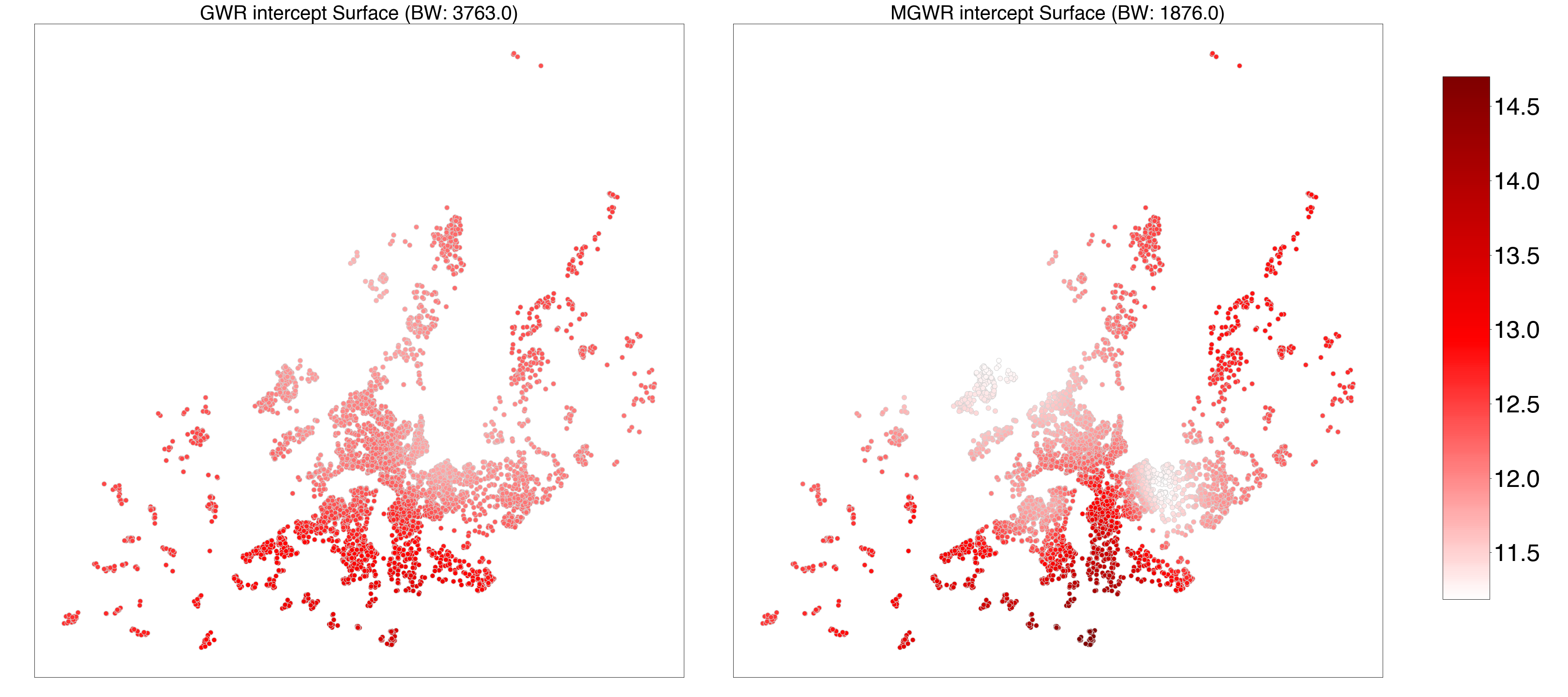

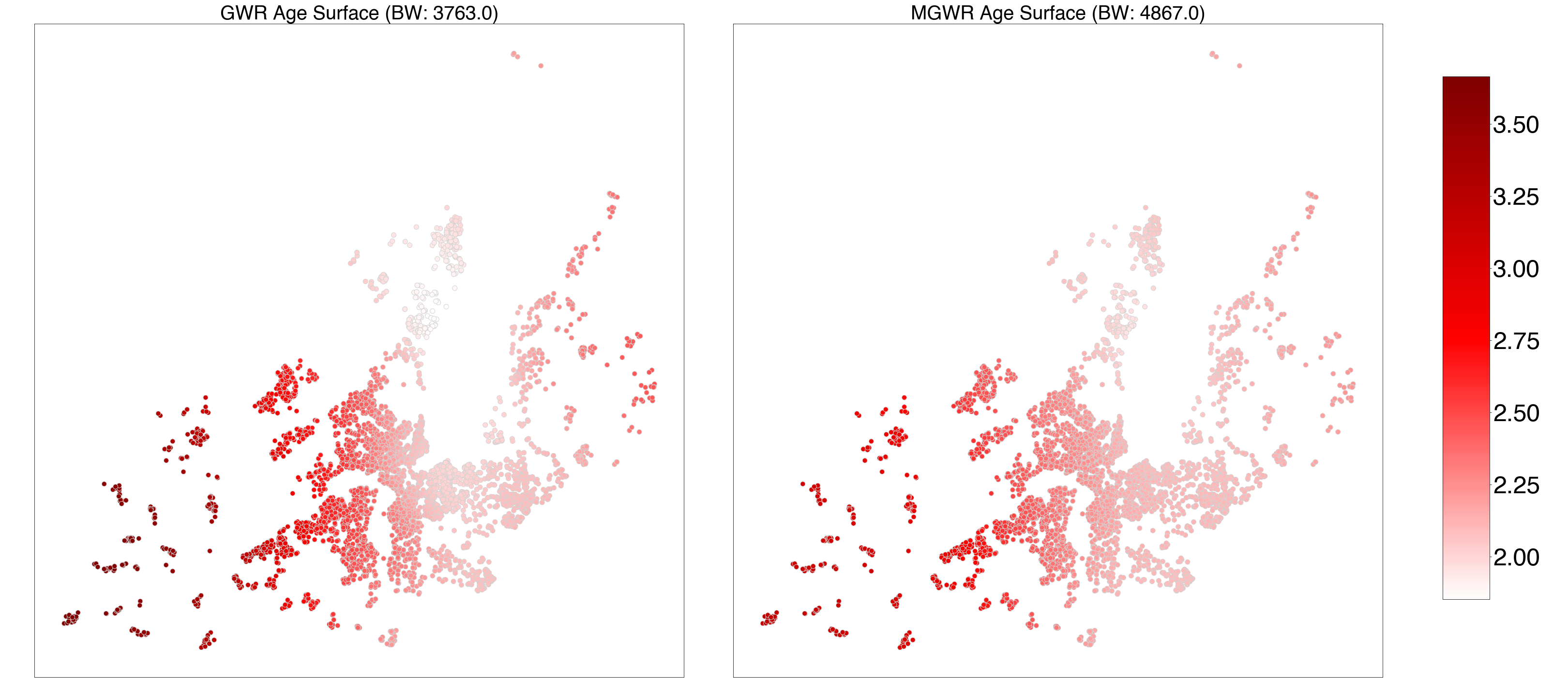

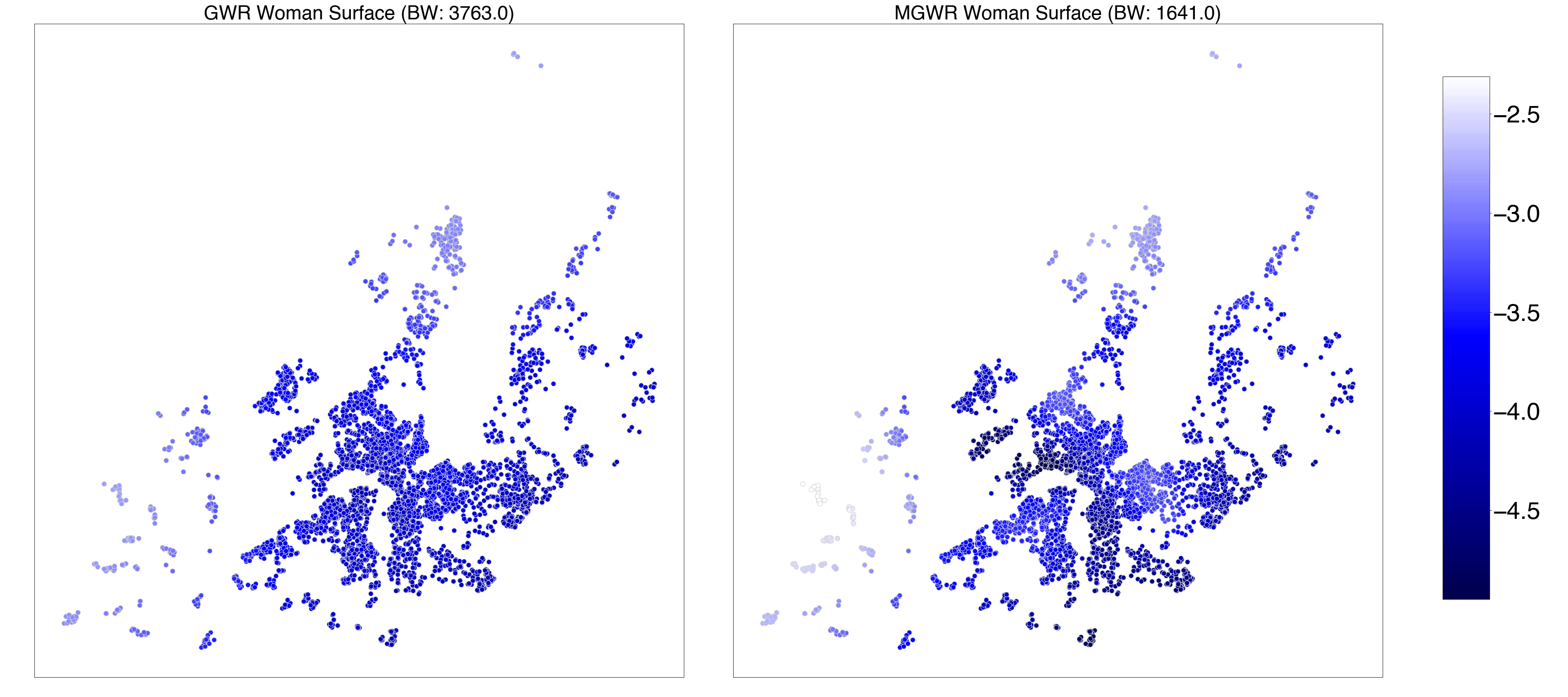

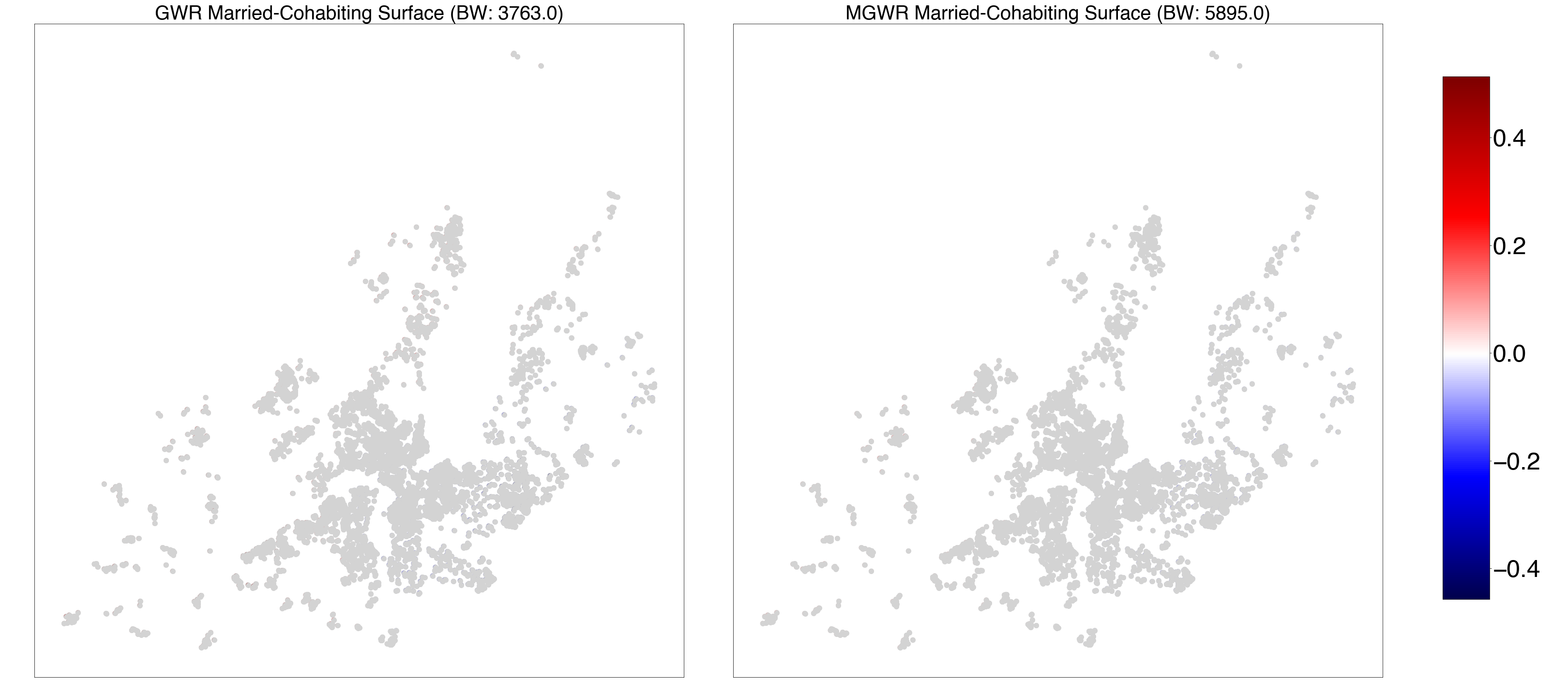

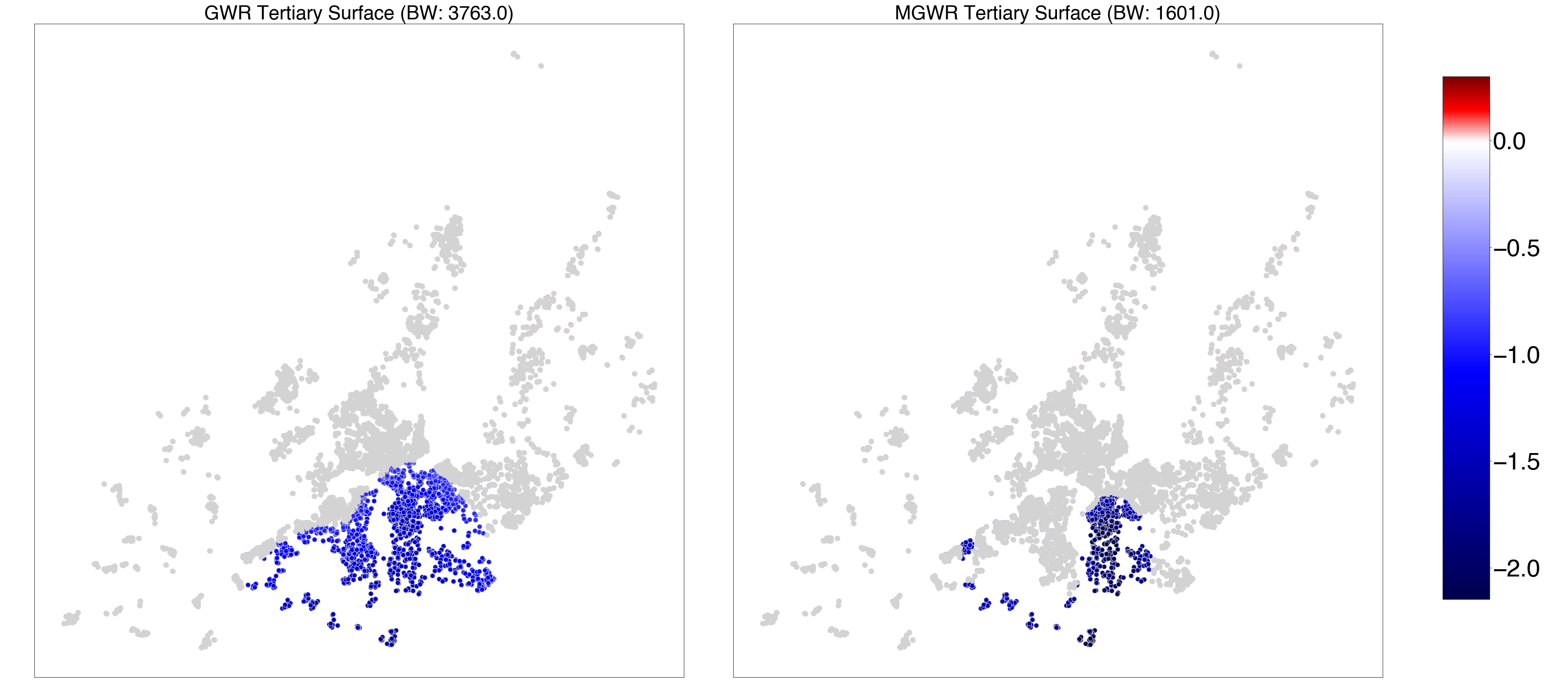

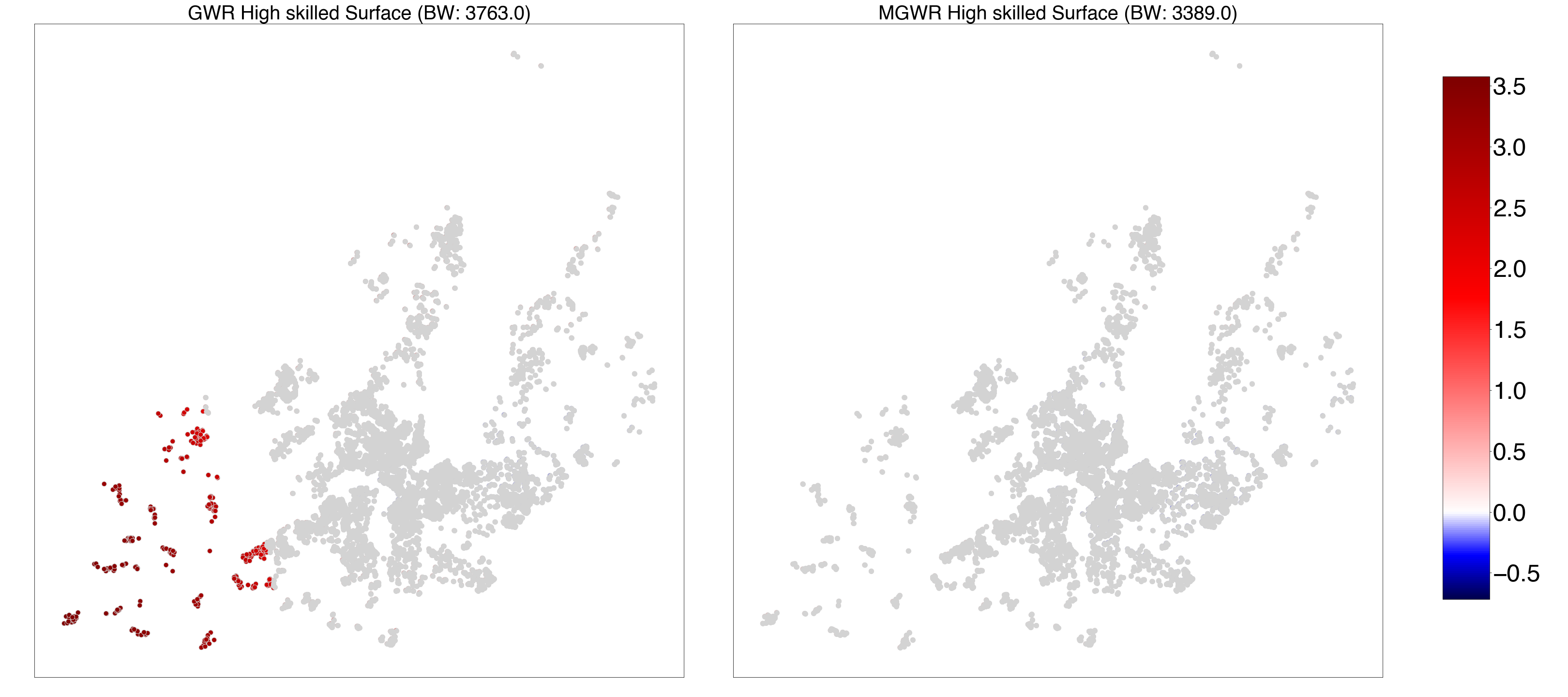

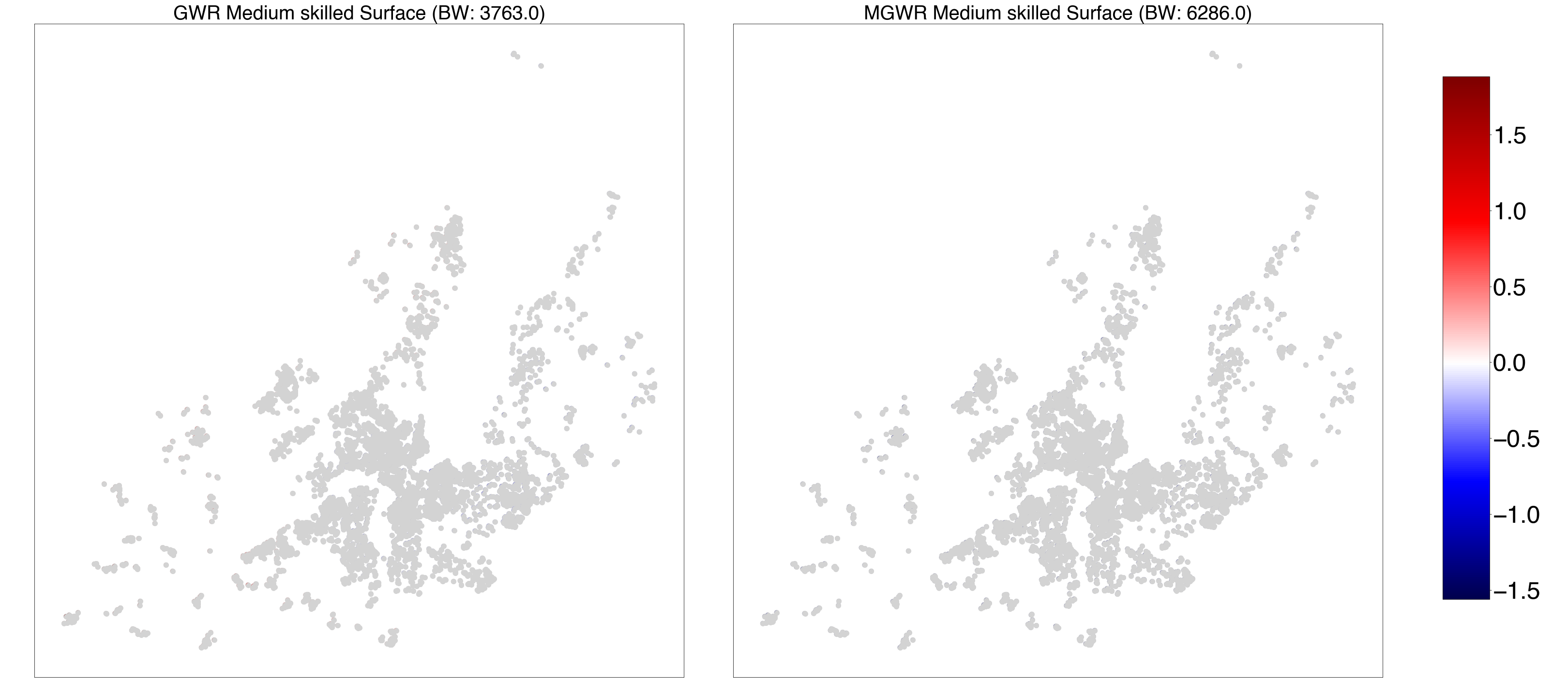

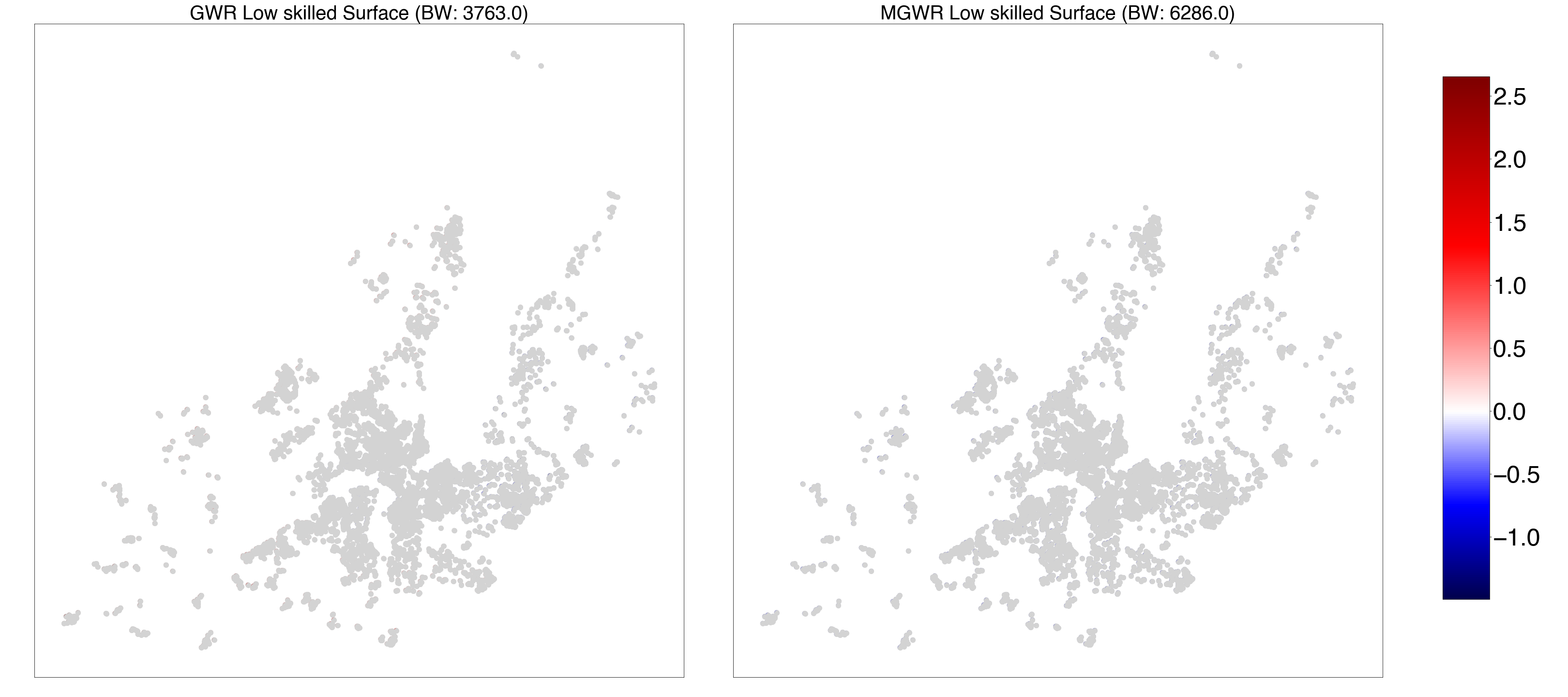

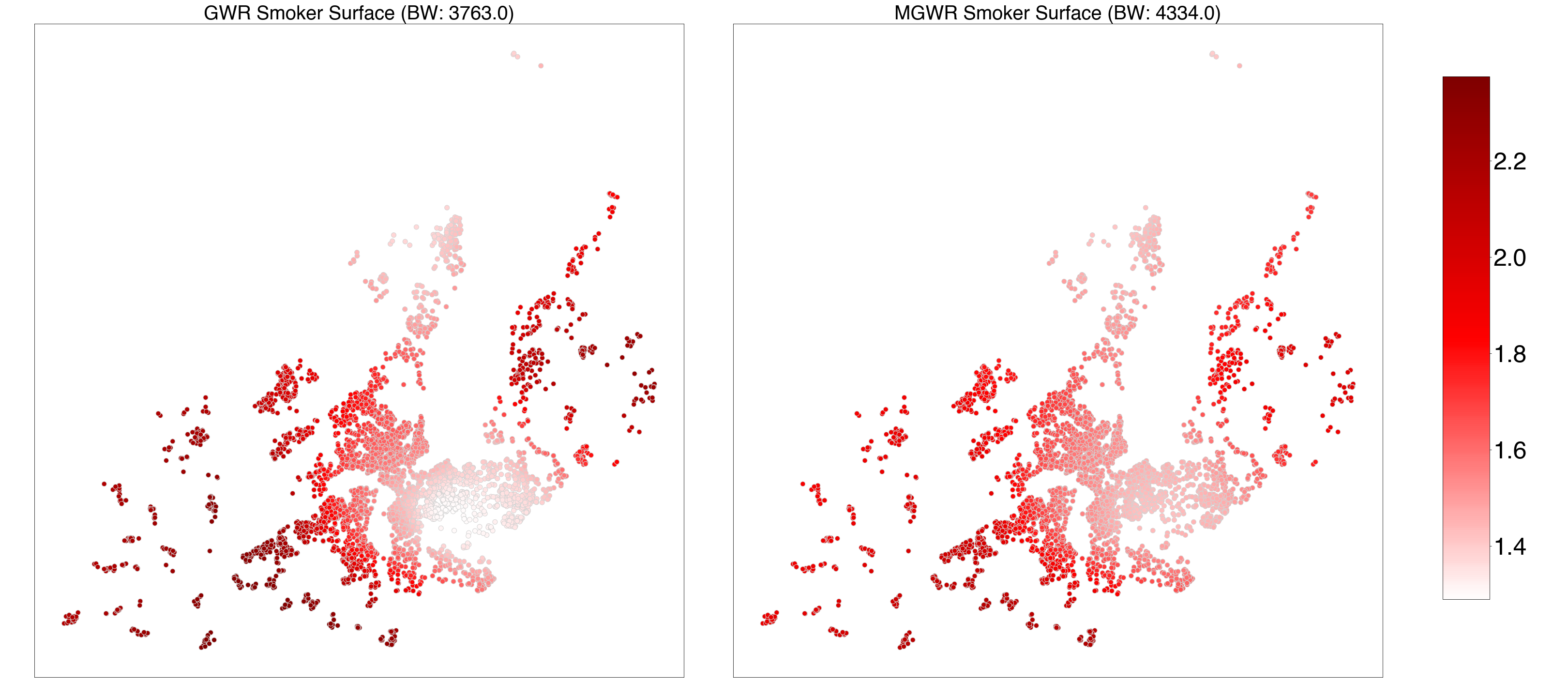

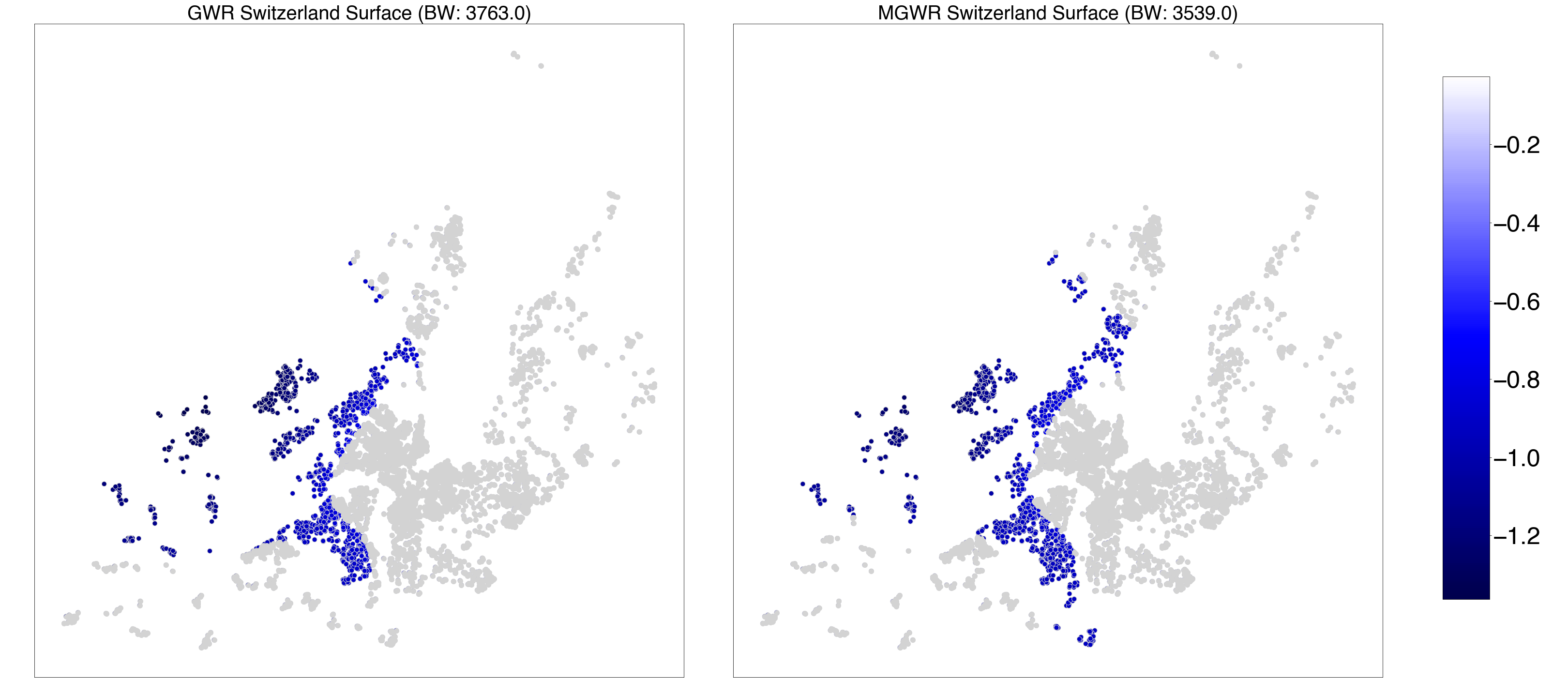

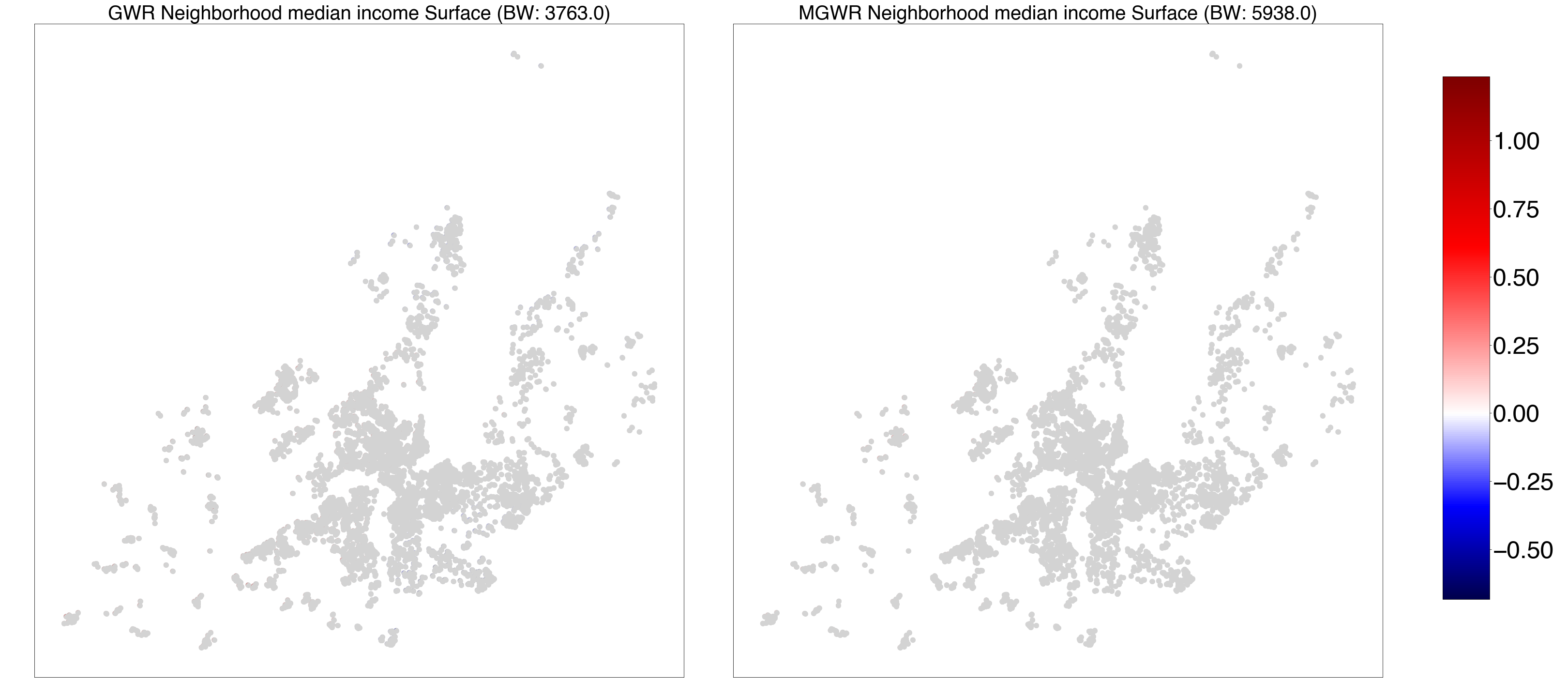

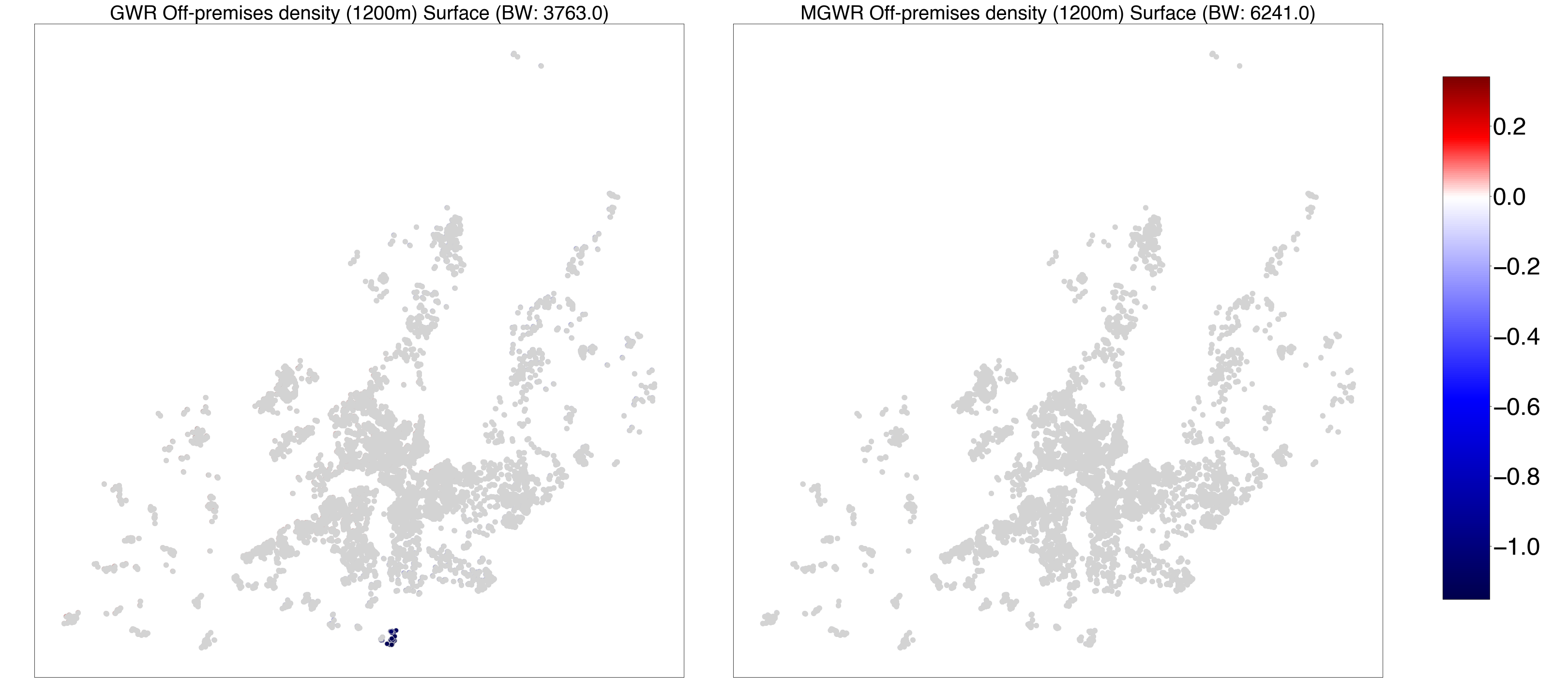

***Figure S3 - GWR and MGWR parameter estimate surfaces for period 3 (P3)—2009.11.01-2018.12.31.*** *Composite maps for geographically weighted regression (GWR) (left) and multiscale GWR (MGWR) (right) parameter estimate surfaces for intercept, age, gender, married-cohabiting, tertiary education, high-, medium-, low-occupational level, smoking status, nationality, neighborhood median household income and off-premises outlet density.*

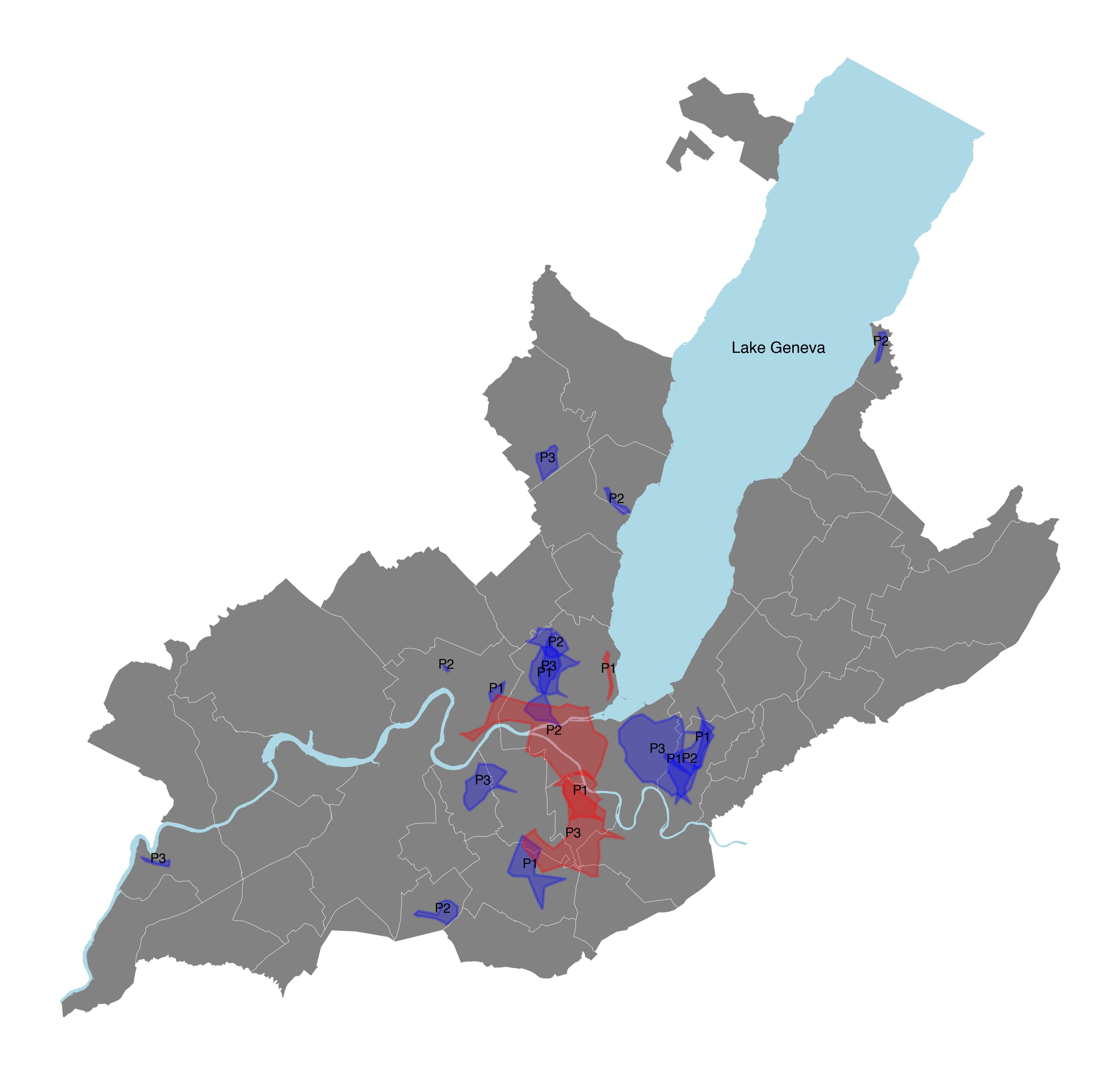

***Figure S4*** *- Hot and cold spots of alcohol consumption across the three study periods. Areas were defined using alpha shapes of the set of points in each high-high and low-low categories.*

**Table S1** - Number of alcohol outlets by year and alcohol outlet category. The periods of participation to the Bus Santé study of each participant are matched to the closest year available in the alcohol outlet dataset.

|  | Off-premise alcohol outlets |  |  |  | On-premise alcohol outlets |  |  |  |  |
| --- | --- | --- | --- | --- | --- | --- | --- | --- | --- |
| REG outlet dataset (year) | Periods of participation | Bus Santé participants (n) | Convenience stores (n) | Grocery stores (n) | Supermarkets (n) | Gas stations (n) | Bars (n) | Restaurants (n) | Clubs (n) |
| 2003 | 1993-2003 | 1250 | 280 | 172 | 43 | 85 | 134 | 1490 | 54 |
| 2005 | 2004-2006 | 1218 | 255 | 160 | 66 | 76 | 140 | 1472 | 47 |
| 2008 | 2007-2009 | 1435 | 269 | 186 | 67 | 79 | 158 | 1536 | 53 |
| 2011 | 2010-2012 | 2150 | 252 | 203 | 56 | 92 | 182 | 1642 | 56 |
| 2014 | 2013-2015 | 1913 | 552 | 259 | 130 | 176 | 374 | 3412 | 92 |
| 2017 | 2016-2018 | 2073 | 524 | 296 | 148 | 172 | 400 | 3558 | 88 |

**Table S2** - Measures of physical accessibility to alcohol outlets by period and category of alcohol outlet.

|  | Period 1 (P1) | | Period 2 (P2) | | Period 3 (P3) | |
| --- | --- | --- | --- | --- | --- | --- |
| Access to alcohol outlets | Mean (SD) | Range | Mean (SD) | Range | Mean (SD) | Range |
| On-premises density (400m) | 7.78 (13.67) | 0.0 - 104.0 | 8.06 (14.36) | 0.0 - 103.0 | 9.76 (16.91) | 0.0 - 115.0 |
| On-premises density (1200m) | 25.66 (37.03) | 0.0 - 201.4 | 26.08 (38.26) | 0.0 - 198.73 | 31.16 (43.18) | 0.0 - 213.5 |
| Bar density (400m) | 0.67 (1.94) | 0.0 - 23.0 | 0.74 (2.02) | 0.0 - 24.0 | 1.03 (2.68) | 0.0 - 28.0 |
| Bar density (1200m) | 2.05 (4.23) | 0.0 - 28.23 | 2.18 (4.37) | 0.0 - 28.84 | 3.16 (5.69) | 0.0 - 39.37 |
| Restaurant density (400m) | 6.69 (11.81) | 0.0 - 85.0 | 6.88 (12.35) | 0.0 - 86.0 | 8.17 (14.29) | 0.0 - 100.0 |
| Restaurant density (1200m) | 21.9 (32.0) | 0.0 - 170.82 | 22.06 (32.91) | 0.0 - 168.29 | 25.7 (36.47) | 0.0 - 174.62 |
| Club density (400m) | 0.41 (0.74) | 0.0 - 5.0 | 0.44 (0.77) | 0.0 - 5.0 | 0.56 (1.05) | 0.0 - 8.0 |
| Club density (1200m) | 1.71 (1.64) | 0.0 - 7.8 | 1.84 (1.8) | 0.0 - 8.43 | 2.29 (2.23) | 0.0 - 12.48 |
| Off-premises density (400m) | 2.08 (3.03) | 0.0 - 18.0 | 1.97 (3.03) | 0.0 - 20.0 | 2.34 (4.06) | 0.0 - 30.0 |
| Off-premises density (1200m) | 6.33 (7.76) | 0.0 - 38.12 | 5.97 (7.57) | 0.0 - 38.17 | 6.83 (8.96) | 0.0 - 46.0 |
| Gas station density (400m) * | 0.16 (0.62) | 0.0 - 6.0 | 0.16 (0.59) | 0.0 - 7.0 | 0.15 (0.66) | 0.0 - 8.0 |
| Gas station density (1200m) * | 0.64 (1.31) | 0.0 - 9.09 | 0.63 (1.32) | 0.0 - 9.24 | 0.62 (1.32) | 0.0 - 8.6 |
| Grocery (small) store density (400m) | 0.35 (0.76) | 0.0 - 5.0 | 0.33 (0.73) | 0.0 - 5.0 | 0.67 (1.4) | 0.0 - 10.0 |
| Grocery (small) store density (1200m) | 0.99 (1.43) | 0.0 - 6.26 | 0.96 (1.4) | 0.0 - 6.29 | 2.0 (2.85) | 0.0 - 18.08 |
| Convenience store density (400m) | 1.29 (1.98) | 0.0 - 17.0 | 1.27 (2.04) | 0.0 - 16.0 | 1.37 (2.68) | 0.0 - 22.0 |
| Convenience store density (1200m) | 3.94 (4.84) | 0.0 - 23.76 | 3.79 (4.85) | 0.0 - 23.73 | 3.92 (5.46) | 0.0 - 28.95 |
| Supermarket density (small) (400m) | 0.02 (0.12) | 0.0 - 1.0 | 0.04 (0.21) | 0.0 - 2.0 | 0.14 (0.5) | 0.0 - 4.0 |
| Supermarket density (small) (1200m) | 0.03 (0.13) | 0.0 - 0.99 | 0.11 (0.29) | 0.0 - 2.2 | 0.46 (0.78) | 0.0 - 4.51 |
| Supermarket density (large) (400m) | 0.21 (0.44) | 0.0 - 2.0 | 0.13 (0.36) | 0.0 - 2.0 | 0.02 (0.15) | 0.0 - 2.0 |
| Supermarket density (large) (1200m) | 0.6 (0.6) | 0.0 - 2.6 | 0.41 (0.55) | 0.0 - 2.65 | 0.08 (0.24) | 0.0 - 2.14 |
| Grocery (large) store density (400m) | 0.05 (0.22) | 0.0 - 1.0 | 0.07 (0.28) | 0.0 - 2.0 | 0.14 (0.55) | 0.0 - 6.0 |
| Grocery (large) store density (1200m) | 0.14 (0.26) | 0.0 - 1.13 | 0.2 (0.35) | 0.0 - 2.14 | 0.36 (0.73) | 0.0 - 5.97 |

*Note: *, excluded from off-premises outlets after 2005; SD, standard deviation*

**Table S3** - Model fit metrics for ordinary least squares (OLS) regression, geographically weighted regression (GWR), and multiscale geographically weighted regression (MGWR)

|  | Model 1 | | | Model 2 | | | Model 3 | | |
| --- | --- | --- | --- | --- | --- | --- | --- | --- | --- |
|  | OLS | GWR | MGWR | OLS | GWR | MGWR | OLS | GWR | MGWR |
| **Period 1** |  |  |  |  |  |  |  |  |  |
| $R^{2}$ | 0.135 | 0.136 | 0.146 | 0.135 | 0.135 | 0.149 | 0.135 | 0.135 | 0.149 |
| AIC | 45390.1 | 45393.3 | 45384.4 | 45392.4 | 45395.6 | 45384.4 | 45392.4 | 45395.6 | 45384.2 |
| AICc |  | 45393.4 | 45385.1 |  | 45395.7 | 45385.5 |  | 45395.7 | 45385.2 |
| GMI | 0.00335 | 0.00319 | 0.00237 | 0.00366 | 0.00347 | -0.0001 | 0.00381 | 0.00363 | -0.00011 |
| GMI p-value | 0.051 | 0.045 | 0.092 | 0.034 | 0.034 | 0.455 | 0.028 | 0.033 | 0.456 |
| **Period 2** |  |  |  |  |  |  |  |  |  |
| $R^{2}$ | 0.143 | 0.143 | 0.146 | 0.143 | 0.143 | 0.146 | 0.143 | 0.143 | 0.146 |
| AIC | 60085.5 | 60089.1 | 60086.8 | 60083.5 | 60087.2 | 60085.2 | 60082.4 | 60086.2 | 60084.2 |
| AICc |  | 60089.2 | 60087 |  | 60087.3 | 60085.4 |  | 60086.3 | 60084.4 |
| GMI | 0.00018 | 0.00002 | -0.00017 | -0.00005 | -0.0002 | -0.00033 | -0.0001 | -0.00024 | -0.00036 |
| GMI p-value | 0.737 | 0.42 | 0.483 | 0.855 | 0.484 | 0.473 | 0.879 | 0.5 | 0.463 |
| **Period 3** |  |  |  |  |  |  |  |  |  |
| $R^{2}$ | 0.113 | 0.124 | 0.129 | 0.113 | 0.128 | 0.129 | 0.113 | 0.127 | 0.129 |
| AIC | 51351.4 | 51342 | 51332.5 | 51351.7 | 51339.6 | 51332.3 | 51351.2 | 51339.8 | 51332.2 |
| AICc |  | 51342.7 | 51333.6 |  | 51340.7 | 51333.4 |  | 51340.8 | 51333.3 |
| GMI | 0.00233 | -0.00017 | -0.00209 | 0.00236 | -0.00083 | -0.0021 | 0.0023 | -0.00085 | -0.00211 |
| GMI p-value | 0.093 | 0.453 | 0.119 | 0.088 | 0.363 | 0.118 | 0.095 | 0.361 | 0.115 |

*Note: GMI, Global Moran’s I; AIC, Akaike information criterion; OLS, ordinary least squares; (M)GWR, (Multiscale) geographically weighted regression*

**Table S4** - Global modeling (OLS) of the associations between socio-demographic and alcohol availability characteristics for each period (n = 18, 515), Bus santé study, Geneva, Switzerland, 1993-2018.

|  | Period 1 (P1) | | | Period 2 (P2) | | | Period 3 (P3) | | |
| --- | --- | --- | --- | --- | --- | --- | --- | --- | --- |
| Variables | Model 1 ($\beta$, SE) | Model 2 ($\beta$, SE) | Model 3 ($\beta$, SE) | Model 1 ($\beta$, SE) | Model 2 ($\beta$, SE) | Model 3 ($\beta$, SE) | Model 1 ($\beta$, SE) | Model 2 ($\beta$, SE) | Model 3 ($\beta$, SE) |
| Intercept | 21.37 (1.82)* | 21.38 (1.82)* | 21.37 (1.82)* | 19.47 (1.54)* | 19.45 (1.54)* | 19.43 (1.54)* | 15.63 (1.27)* | 15.63 (1.27)* | 15.63 (1.27)* |
| Age | 2.28 (0.27)* | 2.27 (0.27)* | 2.26 (0.27)* | 3.18 (0.22)* | 3.18 (0.22)* | 3.18 (0.22)* | 2.44 (0.19)* | 2.44 (0.19)* | 2.44 (0.19)* |
| Gender (Woman) | -11.37 (0.56)* | -11.38 (0.56)* | -11.38 (0.56)* | -10.52 (0.45)* | -10.52 (0.45)* | -10.52 (0.45)* | -7.43 (0.38)* | -7.43 (0.38)* | -7.43 (0.38)* |
| Married-Cohabiting | -0.88 (0.65) | -0.89 (0.65) | -0.88 (0.65) | -0.2 (0.5) | -0.15 (0.5) | -0.13 (0.5) | -0.04 (0.4) | -0.04 (0.4) | -0.06 (0.4) |
| Tertiary | -0.95 (0.64) | -0.93 (0.64) | -0.93 (0.64) | -0.78 (0.5) | -0.81 (0.5) | -0.83 (0.5) | -1.33 (0.43)* | -1.34 (0.43)* | -1.32 (0.43)* |
| High skilled | 4.09 (1.73)* | 4.1 (1.73)* | 4.1 (1.73)* | 4.38 (1.47)* | 4.37 (1.47)* | 4.38 (1.47)* | 2.82 (1.23)* | 2.82 (1.23)* | 2.83 (1.23)* |
| Medium skilled | 1.37 (1.65) | 1.37 (1.65) | 1.38 (1.65) | 1.47 (1.41) | 1.47 (1.41) | 1.48 (1.41) | -0.07 (1.18) | -0.07 (1.18) | -0.07 (1.18) |
| Low skilled | 1.54 (1.67) | 1.54 (1.67) | 1.54 (1.67) | 1.23 (1.42) | 1.21 (1.42) | 1.22 (1.42) | 0.92 (1.18) | 0.92 (1.18) | 0.93 (1.18) |
| Smoker | 7.22 (0.64)* | 7.23 (0.64)* | 7.22 (0.64)* | 6.33 (0.5)* | 6.32 (0.5)* | 6.31 (0.5)* | 4.36 (0.46)* | 4.36 (0.46)* | 4.37 (0.46)* |
| Switzerland | -1.51 (0.61)* | -1.51 (0.61)* | -1.51 (0.61)* | -2.17 (0.48)* | -2.15 (0.48)* | -2.15 (0.48)* | -1.32 (0.41)* | -1.31 (0.41)* | -1.33 (0.41)* |
| Neighborhood median income | 0.87 (0.28)* | 0.83 (0.28)* | 0.84 (0.29)* | 0.34 (0.23) | 0.4 (0.23) | 0.45 (0.24) | 0.13 (0.2) | 0.14 (0.2) | 0.1 (0.21) |
| Bar density (1200m) | 0.55 (0.27)* |  |  | 0.36 (0.22) |  |  | -0.11 (0.2) |  |  |
| On-premises outlet density (1200m) |  | 0.36 (0.28) |  |  | 0.49 (0.23)* |  |  | -0.07 (0.2) |  |
| Off-premises outlet density (1200m) |  |  | 0.37 (0.28) |  |  | 0.55 (0.23)* |  |  | -0.15 (0.2) |

*Note: All continuous predictors are mean-centered and scaled by 1 standard deviation; *p-value < 0.05; SE, standard e*

**Table S5** - Bandwidths and parameter estimates (mean, SD) for multiscale geographically weighted regression (MGWR)

|  | Period 1 (P1) | | | | |  | Period 2 (P2) | | | | |  | Period 3 (P3) | | | | |  |
| --- | --- | --- | --- | --- | --- | --- | --- | --- | --- | --- | --- | --- | --- | --- | --- | --- | --- | --- |
|  | Model 1 |  | Model 2 |  | Model 3 |  | Model 1 |  | Model 2 |  | Model 3 |  | Model 1 |  | Model 2 |  | Model 3 |  |
| Variables | BW | $\beta$ mean (SD) | BW | $\beta$ mean (SD) | BW | $\beta$ mean (SD) | BW | $\beta$ mean (SD) | BW | $\beta$ mean (SD) | BW | $\beta$ mean (SD) | BW | $\beta$ mean (SD) | BW | $\beta$ mean (SD) | BW | $\beta$ mean (SD) |
| Age | 2966 | 2.15 (0.22) | 4867 | 2.16 (0.22) | 4507 | 2.16 (0.22) | 4867 | 3.07 (0.56) | 2844 | 3.08 (0.54) | 4507 | 3.07 (0.54) | 4507 | 2.27 (0.22) | 2966 | 2.27 (0.22) | 4867 | 2.27 (0.22) |
| Woman | 7012 | -5.84 (0.09) | 1641 | -5.84 (0.09) | 5079 | -5.84 (0.09) | 1641 | -5.25 (0.03) | 7012 | -5.25 (0.03) | 5079 | -5.25 (0.03) | 5079 | -3.79 (0.46) | 7012 | -3.79 (0.46) | 1641 | -3.79 (0.46) |
| Married-Cohabiting | 6976 | -0.35 (0.02) | 5895 | -0.35 (0.02) | 5213 | -0.35 (0.02) | 5895 | -0.1 (0.06) | 6976 | -0.08 (0.06) | 5213 | -0.07 (0.06) | 5213 | -0.03 (0.09) | 6976 | -0.03 (0.09) | 5895 | -0.04 (0.09) |
| Tertiary | 7010 | -0.45 (0.01) | 1601 | -0.44 (0.01) | 5213 | -0.44 (0.01) | 1601 | -0.46 (0.02) | 7010 | -0.48 (0.02) | 5213 | -0.48 (0.02) | 5213 | -0.79 (0.54) | 7010 | -0.79 (0.54) | 1601 | -0.79 (0.54) |
| High skilled | 7012 | 0.76 (1.26) | 3389 | 0.71 (1.12) | 813 | 0.72 (1.12) | 3389 | 1.63 (0.02) | 7012 | 1.62 (0.02) | 941 | 1.63 (0.02) | 941 | -0.17 (0.41) | 7012 | -0.17 (0.41) | 3389 | -0.17 (0.41) |
| Medium skilled | 7012 | -0.17 (0.02) | 6286 | -0.23 (0.02) | 5213 | -0.22 (0.02) | 6286 | 0.61 (0.03) | 7012 | 0.61 (0.029) | 5213 | 0.61 (0.03) | 5213 | -1.31 (0.02) | 7012 | -1.32 (0.02) | 6286 | -1.31 (0.02) |
| Low skilled | 7012 | -0.13 (0.03) | 6286 | -0.18 (0.03) | 5213 | -0.17 (0.03) | 6286 | 0.58 (0.03) | 7012 | 0.57 (0.03) | 5213 | 0.57 (0.03) | 5213 | -0.94 (0.03) | 7012 | -0.94 (0.03) | 6286 | -0.93 (0.03) |
| Smoker | 6261 | 2.95 (0.44) | 4334 | 2.97 (0.44) | 2003 | 2.96 (0.44) | 4334 | 2.64 (0.14) | 6261 | 2.63 (0.14) | 2003 | 2.63 (0.142) | 2003 | 1.64 (0.18) | 6261 | 1.63 (0.18) | 4334 | 1.64 (0.18) |
| Switzerland | 4563 | -0.71 (0.07) | 3539 | -0.74 (0.07) | 5057 | -0.73 (0.07) | 3539 | -1.21 (0.33) | 4563 | -1.2 (0.33) | 5057 | -1.2 (0.33) | 5057 | -0.51 (0.33) | 4563 | -0.51 (0.33) | 3539 | -0.52 (0.33) |
| Neighborhood |  |  |  |  |  |  |  |  |  |  |  |  |  |  |  |  |  |  |
| median income | 7012 | 0.74 (0.01) | 5938 | 0.58 (0.036) | 5213 | 0.62 (0.04) | 5938 | 0.28 (0.02) | 7012 | 0.34 (0.03) | 5213 | 0.39 (0.03) | 5213 | 0.1 (0.09) | 7012 | 0.1 (0.09) | 5938 | 0.08 (0.09) |
| Bar density (1200m) | 7012 | 0.51 (0.12) |  |  |  |  | 6241 | 0.29 (0.01) |  |  |  |  | 5213 | -0.02 (0.01) |  |  |  |  |
| On-premises |  |  |  |  |  |  |  |  |  |  |  |  |  |  |  |  |  |  |
| outlet density (1200m) |  |  | 6241 | 0.24 (0.01) |  |  |  |  | 7010 | 0.44 (0.01) |  |  |  |  | 7012 | -0.01 (0.02) |  |  |
| Off-premises |  |  |  |  |  |  |  |  |  |  |  |  |  |  |  |  |  |  |
| outlet density (1200m) |  |  |  |  | 4188 | 0.4 (0.01) |  |  |  |  | 5213 | 0.51 (0.01) |  |  |  |  | 6241 | -0.09 (0.02) |

*Note: SD, standard deviation; BW, Bandwidth*

**Table S6** - Global Moran’s I statistics, z-scores and p-values calculated incrementally at a fixed distance band of 200m, 400m, 600m, 800m, 1000m and 1200m.

| Period | Distance (m) | Moran’s I | p-value | z-score |
| --- | --- | --- | --- | --- |
| Period 1 (P1) |  |  |  |  |
|  | 200 | 0.002835 | 0.336 | 0.409427 |
|  | 400 | 0.01113 | 0.003 | 2.877306 |
|  | 600 | 0.00618 | 0.026 | 2.019882 |
|  | 800 | 0.003421 | 0.081 | 1.426879 |
|  | 1000 | 0.002301 | 0.127 | 1.098764 |
|  | 1200 | 0.002295 | 0.113 | 1.245588 |
| Period 2 (P2) |  |  |  |  |
|  | 200 | 0.008293 | 0.06 | 1.565672 |
|  | 400 | 0.004359 | 0.066 | 1.532462 |
|  | 600 | 0.000091 | 0.475 | 0.059402 |
|  | 800 | 0.001245 | 0.228 | 0.75365 |
|  | 1000 | 0.000711 | 0.275 | 0.487146 |
|  | 1200 | 0.000468 | 0.329 | 0.368993 |
| Period 3 (P3) |  |  |  |  |
|  | 200 | 0.002404 | 0.299 | 0.442388 |
|  | 400 | 0.003024 | 0.157 | 0.947753 |
|  | 600 | 0.00193 | 0.185 | 0.817132 |
|  | 800 | 0.000869 | 0.31 | 0.469111 |
|  | 1000 | 0.001966 | 0.138 | 1.113678 |
|  | 1200 | 0.00263 | 0.041 | 1.857616 |
